# Separating the genetics of disease, treatment, and treatment response using graphical modeling and large-scale electronic health records

**DOI:** 10.1101/2025.04.29.25326633

**Authors:** Malgorzata Borczyk, Nick Machnik, Jacek Hajto, Ilse Krätschmer, Paula Konowalska, Bartosz Baszkiewicz, Michal Korostynski, Matthew R. Robinson

## Abstract

Genetic variants affect baseline health and biomarker values, which in turn may impact both the therapy selected for an individual and the magnitude of change induced by the medication. Here, we propose an approach for complex longitudinal repeated measures biobank data, which separates genetic effects for disease from the genetic effects for medication usage and those for treatment response. For 211,845 individuals, we construct a pre-post study design from 1,420,443 repeated blood pressure (BP) measurements and 1,117,900 prescription records for common BP influencing drugs, using electronic health records. We model these jointly alongside 8,430,446 imputed single nucleotide polymorphism (SNP) markers and 17,852 whole-exome sequence loss-of-function (LoF) variants, all within a single novel graphical modeling framework. We identify pharmacogenetic candidate SNPs and LoF variants in genes *SLC35F2, PKD1* and *KCNIP4*, which are associated with angiotensin receptor blocker therapy and response after controlling for hypertensive disease status across multiple world-wide biobanks. We additionally detect and replicate established clinically relevant variants for statin treatment across multiple biobanks. We find that genetic variation for BP is predominantly shaped prior to the age of 50, but we identify 127 independent loci associated with age-specific BP changes later in life. Finally, once post-treatment measures are conditioned on pre-treatment measures and therapy, we find evidence for four independent loci influencing BP treatment response, including a variant in *ADAMTSL1* which has previously been associated with diuretic and beta-blocker response. Our graphical modeling and pre-post study design provides a robust way of detecting time-, treatment- and treatment response-specific genetic associations within large-scale biobank studies.

## Introduction

Several large-effect pharmacogenomic genes are commonly tested in the clinic to guide therapy administration and dosing, which reduces the incidence of adverse drug reactions [1, 2]. While recent studies have identified genetic variants associated with medication use and shown that baseline biomarker genetic predictors correlate with treatment [3–5], current evidence suggests that for most cardio-metabolic therapies, there are few clinically relevant gene-treatment interactions for treatment response, predominantly detected for statins [6–12].

Pharmacogenes have been identified through genome-wide association studies (GWASs), in randomized controlled trials (RCTs), and more recently in pharmacogenomic studies within large epidemiological biobank cohorts [1–13]. Typically, drug response phenotypes are often measured in pre- and post-treatment designs in the form of an absolute or relative change in a quantitative trait before and after a particular treatment. In the context of pharmacogenetic effects, the focus is on estimating genetic effects on biomarker change. However, genetic variants also affect baseline values, which in turn impact both the therapy selected for a patient and the magnitude of change induced by the medication [6–12]. This is evidenced by analyzes on cross-sectional and longitudinal data, showing that genetic predisposition to high baseline biomarker value is a reliable indicator of treatment response [3, 5, 11, 12]. Furthermore, follow-up polygenic risk score analyses and studies examining the genetic basis of being placed on particular treatments show that baseline biomarker genetic predictors correlate with the therapy given [5]. This makes separating genetic effects for the disease from the genetic effects for medication usage and those for treatment outcomes challenging. Additionally, interaction studies are susceptible to issues of scale-dependency, endogeneity bias from the relationship of therapy and baseline phenotype [12, 14], and ascertainment bias by sub-setting to individuals with complete pre-post records. These issues currently limit our ability to resolve the genetic basis of treatment and treatment response, especially when using complex large-scale biobank data linked to electronic health records for pharmacogenomic research.

Here, we propose an approach to separate genetic effects on baseline biomarker values from genetic effects for both being placed on a particular therapy and the response of the biomarker to therapy (Figure 1). We show how by learning a graphical model that is time-aware and handles repeated measures, genetic variants associated with a particular treatment are identified, conditional on the clinical measure that is the focus of the treatment. Associations for post-treatment response are also identified conditional on both the treatment made and the pre-treatment baseline measure, which separates the disease and the therapy choice from the response to the treatment. We utilize 1,420,443 repeated blood pressure (BP) measurements, annotated with 1,117,900 prescriptions for 72 drugs used in the management of BP grouped into six classes, 47,121 diagnoses of cardiovascular disease, alongside with 8,430,446 imputed SNPs and 17,852 whole exome sequence (WES) data loss of function (LoF) variants, which are all jointly analyzed in a single model (see Figures 1 and S1). After conditioning post-treatment measures on pre-treatment measures and therapy, we find evidence for four genetic variants influencing BP treatment response. We find pharmacogenetic candidate SNPs and LoF variants for angiotensin receptor blocker therapy and response, after controlling for hypertensive and cardiovascular disease status. Finally, we show that genetic variation for BP is predominantly shaped prior to the age of 50, with additional variants contributing to age-specific BP changes throughout life.

**Fig. 1:**
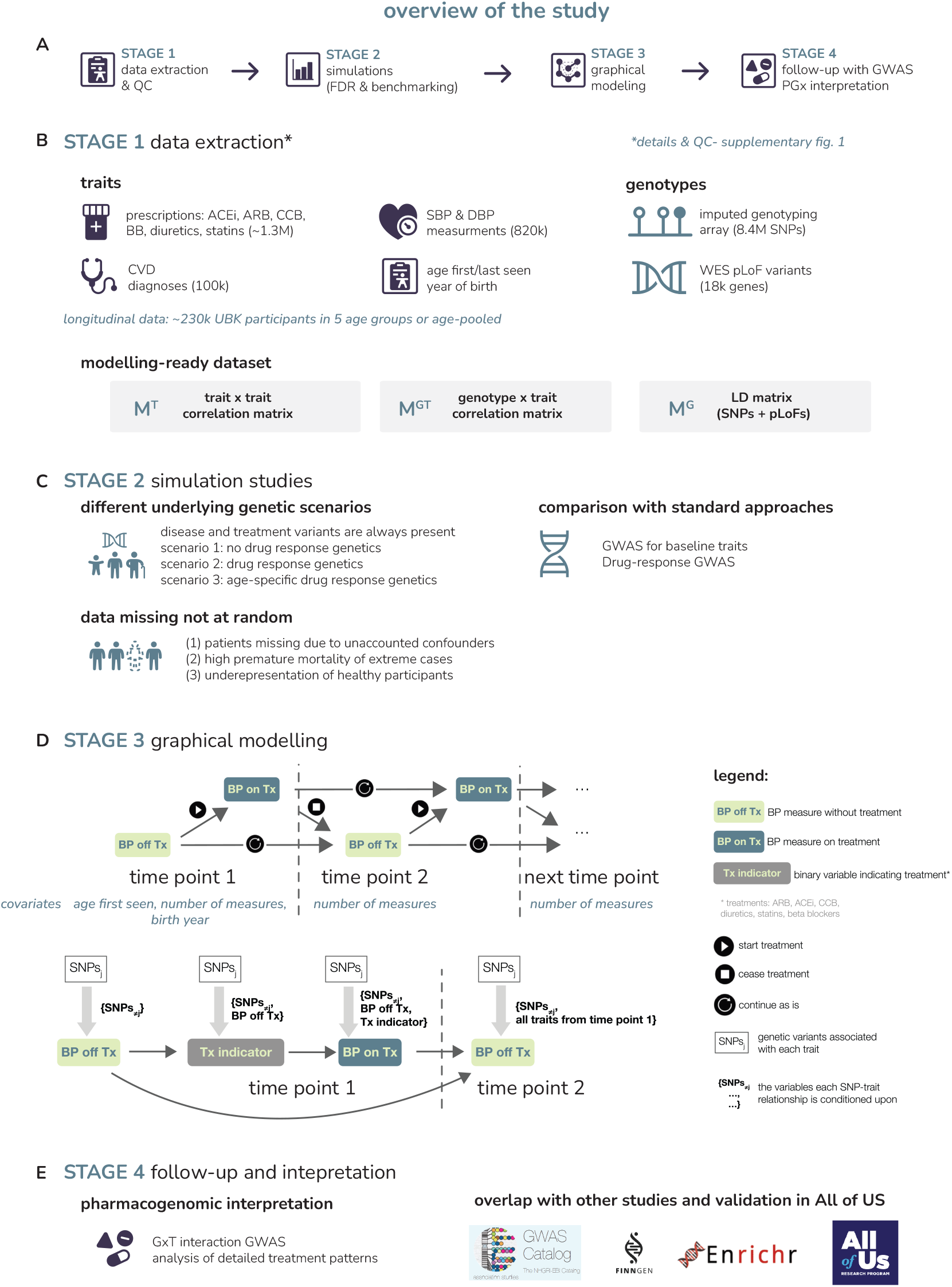
Overview of the study. (A) The structure of our study. (B) We select traits and process the data to create a pre-post design and genetic variants datasets ready for modeling. (C) We conduct simulation studies to validate our modeling approach. We study different scenarios of genetic effects on drug response traits, compare to standard modeling approaches and examine the effects of different missing data patterns. (D) We apply our graphical model to the UK Biobank data. Shown is a schematic representation of the way blood pressure (BP) measurements are aggregated and grouped into separate time points (i.e. age groups) for the age-split analysis. For each participant, we determine whether their BP measure was taken when they were not on treatment (“off Tx”) or treated with a BP influencing drug (“on Tx”). We include covariates either as baseline values (e.g. birth year) or as time point specific variables (e.g. number of measures, see Methods). Below this, we then give an overview of the conditioning scheme in the time series analysis. At the first time point, the association of off-treatment BP with each single nucleotide polymorphism (SNP) is conditioned on all other genetic variants (indicated within the brackets). For all succeeding traits, each SNP association is determined conditionally on all other SNPs as well as on temporally preceding traits, with the conditioning sets given within the brackets. Blood pressure variables are continuously distributed and the six treatment indicator variables are binary (0 for untreated, 1 for treated). (E) We then conduct a pharmacogenetic follow-up study to aid interpretation, alongside replication and enrichment analyses.

## Results

### Pre-post study design

Here we present an approach to modeling complex longitudinal biobank data, where we construct a large-scale pre-post study design by extracting data from a combination of electronic health records (EHRs), drug prescription databases, and genomic information (Figure 1B, Figure S1). We stratify clinical measurements into multiple age groups, each containing pre- and post-treatment values. We then include a number of additional variables: binary indicator variables for each treatment received within each age group, covariates specific to age groups, and several additional variables that reflect the structure of the measurements recorded for each individual (measurement source, ages first and last seen in the GP records, year of birth, etc.), for which we wish to adjust (see Methods for full details).

We then apply a novel time-aware graphical inference algorithm to these data that learns the relationships between all observed variables. The algorithm conducts conditional independence (CI) tests between different combinations of variables, to determine whether variables are marginally or conditionally independent of each other (Figure 1D). Since the data are ordered in time, the algorithm uses this prior knowledge when testing for independence between two variables, including only preceding variables or those at the same time point in the conditioning sets (see Methods, Algorithm 1). For example, for the first time point (age group), the CI tests between pre-treatment measures and genetic variants use only the genetic variants and data structure variables as conditioning variables. CI tests between treatment indicators and genetic variants use genetic variants, the data structure variables and the pretreatment measure as conditioning variables. As a result, SNPs associated with being placed on a particular treatment are those that remain significantly correlated after adjusting for the clinical measure that is the focus of the treatment. Finally, CI tests for the first time point (age group) post-treatment measure condition on the treatment indicator, the pre-treatment measure as well as all other genetic variants and data structure variables. Thus, SNP associations for post-treatment measures are conditional on the treatment made and the pre-treatment measure and can be interpreted as being correlated with the change in the clinical measure post-treatment. Moving forward in age, measures made later in life are then conditioned on measures at all previous time points. As a result, SNP associations are time-point specific for the treatment indicators and for the pre- and post-treatment clinical values.

We choose the cut-off ages between groups so that time points have similar numbers of participants to balance the power to detect associations. In biobank applications, not all individuals will have measures at all time points. Our analysis allows for this, as it is based entirely on summary statistic information, consisting only of the pairwise genetic marker-marker (LD) correlations, the pairwise genetic marker-variable correlations, and the pairwise variable-variable correlations (Figure 1). It relies on partial correlations vanishing for (conditionally) independent variables, which does not hold for binary or ordinal variables at low frequencies. To alleviate this issue, we use a heterologous correlation matrix, with Pearson correlations between Gaussian, polyserial between Gaussian and ordinal, and polychoric correlations between ordinal variables, as previously proposed [15, 16]. This effectively places binary and ordinal variables on the liability scale and has the additional advantage of allowing comparisons among variables on the same scale, accounting for differences in their prevalence. We also employ a false discovery rate correction to control for the multiple CI tests conducted within the algorithm (see Methods). This builds upon our recently proposed CI-GWAS approach [16].

### Robust identification of age- and trait-specific genetic effects

First, we conduct a simulation study in order to confirm the validity and effectiveness of our modeling approach. Our simulation is based on a structural equation model (SEM) of a time-series of blood pressure readings and medication status and, observed at two age groups, and time- and trait-specific genetic variables (see Figure 2A and Methods). For the genetic variables, we use real observed imputed SNP data on chromosome 1 from 458,747 UK Biobank individuals. In addition, we include a latent unobserved random variable in the SEM to reflect potential unobserved confounding factors common to population studies at baseline. In the time-series, we simulate three types of measures, each observed at time points, here called age groups: *(i)* pretreatment blood pressure; *(ii)* binary indicators of treatment with two different drugs; and *(iii)* post-treatment blood pressure, which is defined only for individuals who received treatment in the same age group. Whether the pre-treatment blood pressure is defined at the second age group also depends on an individual’s treatment history. Firstly, individuals who are not placed on treatment in the first age group have a pre-treatment observation in the second age group. Secondly, a proportion of individuals are simulated to have stopped treatment between the two observation periods, and these also have pre-treatment observations in the second age group. A graphical presentation of the simulation design is shown in Figure 2A.

**Fig. 2:**
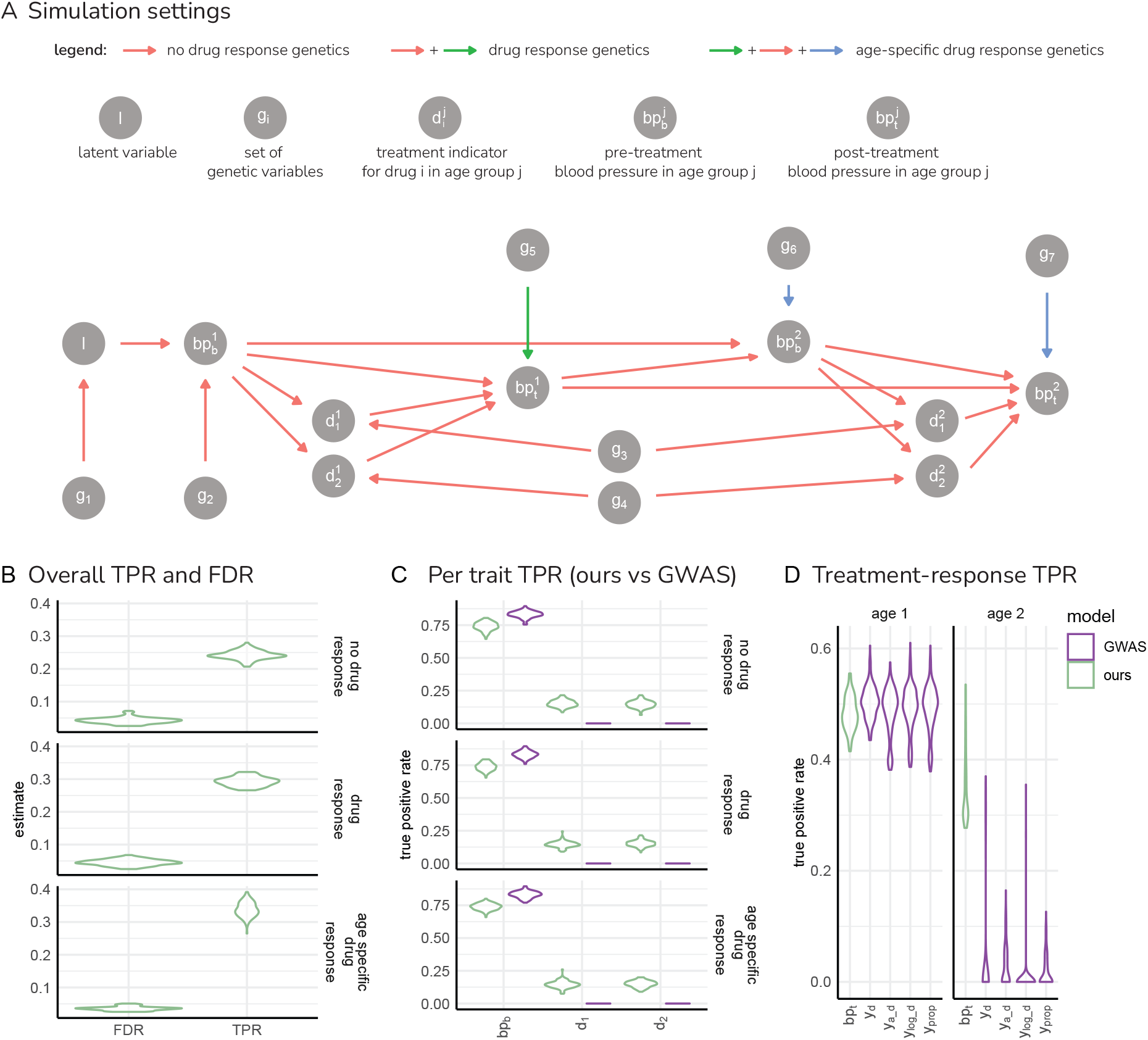
Detection of time- and trait-specific genetic effects in simulation. (A) We simulate three main settings, shown as directed acyclic graph. l: unobserved latent variable; *g*_1_ to *g*_7_: represent 100’s of genetic variables influencing each trait; 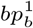 and 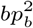: simulated baseline (pre-treatment) blood pressure measures at age group 1 and 2 respectively (indicated in superscript); 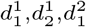 and 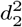: simulated indicator variables for two different drug treatment groups (indicated in subscript) at age group 1 and 2 respectively (indicated in superscript); 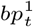 and 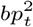: simulated post-treatment blood pressure measures at age group 1 and 2 respectively (indicated in superscript). For each setting, we use SNP data of chromosome 1 for 458,747 individuals and run 100 replicates. Red arrows indicate the “no drug response” setting; the “drug response” setting has the addition of genetic effects (*g*_5_) on 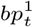 (green arrow); and the “age-specific drug response” setting has the addition of genetic effects, *g*_6_ and *g*_7_ on 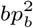 and 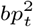 respectively (blue arrows). (B) False discovery rate (FDR) and true positive rate (TPR) for the discovery of associated genetic variables for all traits when applying our graphical algorithm to the simulated data of each setting. (C) True positive rate of our graphical algorithm (labeled “ours” in green) for the discovery of associated genetic variables for simulated baseline (pre-treatment) blood pressure and simulated indicator variables for two different drug treatment groups. We compare the TPR of our graphical approach when discovering post-treatment genetic variables to traditional genome-wide association study estimates (labeled “GWAS” in purple). (D) True positive rate of our graphical algorithm for the discovery of associated genetic variables for each trait at each age group (labeled “age 1” and “age 2”). We compare the TPR of our graphical approach when discovering post-treatment genetic variables to previously proposed genome-wide association study approaches by calculating: the absolute difference of baseline pre-treatment and post-treatment measures (*y*_*d*_ = |*y*_*post*_ −*y*_*pre*_|); the logarithmic relative difference (*y*_*log*_*_*_*d*_ = *log*(*y*_*post*_) −*log*(*y*_*pre*_)); the proportional change (*y*_*prop*_ = *y*_*post*_/*y*_*pre*_); and the difference adjusted for baseline levels (*y*_*a*_*_*_*d*_ = |*y*_*post*_− *y*_*pre*_| −*y*_*pre*_*β*, where *β* is the regression coefficient of the effect of the baseline on the difference). Conditional and joint analysis is conducted for the standard genome-wide association study approaches.

We use this framework to simulate three different scenarios, each with 100 replicates of sample size 458,747. The settings differ in whether there are simulated SNP effects for: pre-treatment blood pressure at age 1 and drug treatment only (labeled “no drug response genetics”); pre- and post-treatment blood pressure at age 1 and drug treatment only (“drug response genetics”); or all measures at all ages (“age specific drug response genetics”). We apply our graphical inference algorithm to each of these 300 datasets, and infer SNP marker associations for the eight traits (two drug indicators, a pre-treatment and post-treatment measure at each age group) jointly within a single model. We show that across the 300 sets of data the false discovery rate (FDR) for the discovery of associated genetic variables for all traits, calculated as the proportion of marker associations returned by our algorithm that are not true associations, remains well-controlled around the 5% threshold set by our FDR multiple testing control (Figure 2B). The true positive rate (TPR) for the discovery of associated genetic variables for all traits, calculated as the proportion of true simulated SNP associations that are returned by our algorithm, is also shown in Figure 2B and is then split across the eight traits in Figure 2C and D. The TPR varies according to the sample size of the trait, with 458,747 individuals having pre-treatment measures at age group 1, approximately 35,000 individuals being on each treatment at each age, 54,000 individuals being on-treatment at age group 1 with post-treatment measures and around 100,000 individuals receiving treatment by the second age group (which is comparable to the UK Biobank data presented below).

At a sample size of *n* = 450, 000, our procedure has a theoretical limit of detection for a variant at 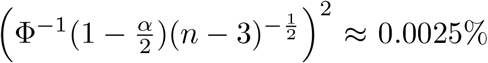 explained, where Φ is the cumulative density function of the standard normal distribution and *α* is a sensitivity parameter for the CI tests (see Methods for full details). This also corresponds to the power for this effect size in a standard genome-wide association study, which would be 90% at the genome-wide significance level of 5 · 10−8. Thus, we expect our approach to have the same power as standard association testing. We show empirically that the TPR and FDR of our approach is just slightly below that of marginal association testing for simulated baseline pre-treatment blood pressure (Figure 2C). We also show that conditioning on pre-treatment measures within our graphical model gives higher TPR for discovering SNPs associated with drug treatment (Figure 2C) as compared to genome-wide marginal association testing of each drug indicator independently (Figure 2). Power calculations for dichotomous traits are given in the Methods, with values given in Figure S2.

Additionally, we compare to age-stratified genome-wide association analyzes of pre-post treatment blood pressure for each of the 300 data sets. Previous association testing studies (e.g. Ref [11]) propose defining drug response phenotypes as either: (i) the absolute difference of baseline pre-treatment and post-treatment measures (| *y*_*post*_− *y*_*pre*_|); *(ii)* the logarithmic relative difference (*log*(*y*_*post*_) −*log*(*y*_*pre*_)); *(iii)* the proportion *y*_*post*_/*y*_*pre*_; or *(iv)* the difference adjusted for baseline levels (|*y*_*post*_ −*y*_*pre*_| −*y*_*pre*_*β*, where *β* is the regression coefficient of the effect of the baseline on the difference). For each of the 300 data sets, we conduct marginal association testing for each of these four drug response phenotypes using the UK Biobank chromosome 1 data observed for 458,747 individuals. We then conduct conditional and joint analysis using the COJO approach of GCTA to facilitate a comparison of the TPR and FDR. As expected, at the first age group, the power (TPR) of our graphical approach is the same as the pre-post response models (Figure 2D left age 1 panel which combines the “drug response genetics” and “age-specific drug response genetics” settings). In the “age-specific drug response” setting, we simulate additional SNP effects that are specific to the second age group and find that our approach has higher power to recover these as compared to an age-stratified pre-post response model (Figure 2D right age 2 panel). Power calculations for post-treatment traits are given in the Methods and values from an additional simulation study with 100 replicates are given in Figure S3 which match the expected values.

In the “age specific drug response genetics” scenario, where we simulate different SNPs associated with both pre- and post-treatment blood pressure values across age groups, we find that our proposed approach results in a higher TPR to discover age-specific treatment-response SNPs as compared to age-stratified pre- and post-treatment blood pressure GWAS (Figure 2D, labeled *bp*_*t*_). This is because within our graphical model SNPs associated with later ages or treatments are identified conditional on preceding measures. In contrast age-stratified GWAS are not conditional, and thus markers discovered in this way are associated across multiple time points. Taken together, we show that our approach accurately selects time- and trait-specific genetic variants accounting for LD down to the theoretical limit of detection given by the sample size, with comparable power to GWAS at the baseline and superior power at later time points.

As time-series data are often subject to data censoring and ascertainment bias, we also explore how structured missing data influence the detection of the simulated causal variants (see Methods). We simulated a series of five data ascertainment settings, for each of the three scenarios, using the same data from chromosome 1 for 458,747 UK Biobank individuals. This gave a further 1,500 data sets.

We find that the FDR is well-controlled in all settings, except when there is strong study ascertainment based on a heritable unobserved latent variable where there is a slight elevation (scenario “C1” in Figure S4). Calculating the FDR for each trait separately shows that this elevation stems from an elevation in false discoveries for the simulated baseline pre-treatment blood pressure (Figure S5). This is expected as selection based on the heritable unobserved latent variable will influence the trait most to which it is most closely related. Note that we give these values to aid in understanding which traits in the model are subject to elevated FDR under different ascertainment scenarios, but that as associations with all traits are determined jointly within a single model, the appropriate FDR of the model is given in Figure S4. Strong study ascertainment in this case corresponds to removal of the top 40% of individuals from the simulation, based on a heritable unobserved latent variable (scenario “C1” in Figure S4), which also serves to reduce power to detect the correct associated variants.

In summary, taken together across a total of 1,800 data sets each of sample size 458,747 individuals, our simulations indicate that our graphical modeling approach provides a robust way of detecting time- and trait-specific associations when data are not very strongly missing at random with respect to the variables studied. We acknowledge that like with any association study, there may be more complex data missing patterns that we do not consider here. Thus, when applying our approach to the genetics of hypertension as we describe below, we additionally include variables that may correlate with data missingness, with the aim of providing additional control for these factors.

### Age-specific genetic effects for blood pressure

We apply our framework to study the genetics of BP and its management through the life-course (Figure 1). From UK Biobank EHRs, we extracted: 1,420,443 repeated BP measurements for 211,845 individuals, a series of likely confounding variables (e.g. number of visits) and 1,117,900 prescription records, from which we created treatment indicators for six drug classes commonly used in the management of hypertension and cardiovascular disease (CVD). For further details, see Methods, Supplementary

Note 1, Table S1, and Figure S1. Of the ~1.4*M* BP measurements, 20.2% were collected at the assessment centers and 79.8% came from GP records. On average, each participant has 6.2 BP measures (Figure S6). The six drug classes we analysed are: Angiotensin-Converting Enzyme Inhibitors (ACEIs), Angiotensin Receptor Blockers (ARBs), Calcium Channel Blocker (CCBs), diuretics used in hypertension management, beta-adrenergic blockers and statins. Although statins only indirectly target BP, we included them as they are the most common cardiometabolic drug class. In our dataset, when analysed in a single model with other drug classes, statins have an effect on both systolic (−1.68 mmHg) and diastolic blood pressure (−1.39 mmHg), albeit smaller than other drug classes (−4.28 mmHg for systolic BP and −1.95 for diastolic BP on average, see S2 and Figure S7).We include all of the treatment indicators, alongside 8,430,446 imputed SNP markers and 17,852 high-confidence loss-of-function (LoF) variants obtained from whole exome sequencing data, within a single graphical model. We compare the results from our model set-up in two ways: (i) age-split, where pre- and post-treatment BP records and binary drug treatment indicators are aggregated into five different age groups of similar sample size; and (ii) age-pooled, where all records are averaged into a single set of pre-post measures for each participant.

Systolic and diastolic blood pressure (SBP and DBP) are strongly phenotypically correlated (*ρ*(*SBP*_*pre*_, *DBP*_*pre*_) = 0.7213 on average across the 5 age groups, with an average *SE* = 0.002). We also find a consistently high genetic correlation for SBP and DBP within ages of 0.585 (0.137 SE) for early life (*<* 50) and 0.569 (0.110 SE) for later age (*>* 65). Thus, we analysed them both jointly and separately, because we observed that if a SNP has effects on both SBP and DBP of very similar magnitude so that *ρ*(*SNP, DBP*) and *ρ*(*SNP, SBP*) are very similar and the phenotypic correlation *ρ*(*SBP, DBP*) is large, then the partial correlation of the SNP with each blood pressure component becomes insignificant when conditioning on the other (Figure S8). While we find a number of SNPs for each blood pressure component when analyzing them jointly, analyzing them separately allowed us to identify more associated variants and better describe the age-related changes in both variables.

We find hundreds of variants associated with pre-treatment SBP or DBP, and only 9 associated with any post-treatment BP at any age(Figure 3A, Table 1, see Supplementary Data Tables for full results). After conditioning on pre-treatment BP measures and potential observed confounders, we find 6 SNPs and 3 LoF variants directly associated with treatment response. As these three LoF variants had only one or two carriers in the treated group, we excluded them from further analysis. Additionally, two of the six variants were absent when statins were not considered as BP treatment (Table S4), leaving four stable BP-response candidate SNPs: one for SBP in the age-pooled approach, two for SBP in the youngest age group, and one for DBP in the 56-60 years group (Table 2). Importantly, these variants are not associated with baseline BP measures, neither in our graphical model nor in a standard GWAS for baseline BP, with rs6065646 (p-value 0.04) having the strongest association (see Supplementary Data Tables). None of these variants are associated with baseline BP neither in GWAS catalog nor in Finngen r12 [18, 19]. One of the selected variants, the missense PANO1 (P210Q) rs191701026 SNP, has been previously associated with pulse pressure (difference between SBP and DBP), but not DBP nor SBP [20]. Another SNP, associated with post-treatment SBP in the youngest age group is rs117686634 in *ADAMTSL1*. Other variants in this gene, involved in vascular remodeling, have been suggested as associated with beta-blocker and thiazide diuretics response [21–23]. As our model includes multiple drugs, we investigated BP response variants in genotype-treatment interaction models for each drug class separately to aid interpretation. Each of these models test the absolute biomarker change (e.g. *deltaSBP* = *SBPpost* **−** *SBPpre*) for an interaction with a binary indicator of prescription for each drug class, both with and without baseline adjustment (see Methods and Supplementary Data Tables). No variants reached genome-wide significance, but rs6065646 (*GTSF1L*) has a p-value of 0.00045 for *deltaSBP* × *diuretic* interaction with baseline adjustment. Overall, our graphical model provides evidence that these four variants affect the magnitude of BP response to treatment without affecting the baseline. To assess the robustness of these findings, we performed a series of sensitivity analyzes, such as adding any CVD diagnoses prior to the first BP record and moving the prescription window (see Table S4). We report that the four BP-response variants are likely around the detection threshold as they are absent from some of the sensitivity models (Table S4). This is consistent with the substantially lower power for post-treatment associations, as the treated sample is approximately a third the size of the untreated sample (Table S5).

**Table 1:** Counts of identified SNPs and whole exome sequencing loss-of-function variants per age group and systolic and diastolic blood pressure (SBP, DBP) traits before and after treatment. To create the LD-pruned input data, SNPs were pre-filtered by setting a maximal correlation threshold of LD *R*^2^ *>* 0.8 within 1MB and selecting a focal variant of highest frequency from each set of highly correlated markers. Only variants at *FDR <* 0.05 are reported.

| Trait | Age | all SNPs |  | LD pruned |  |
| --- | --- | --- | --- | --- | --- |
|  |  | #SNPs | #LoF | #SNPs | #LoF |
| SBP pre-treatment | pooled | 307 | 22 | 329 | 23 |
| SBP pre-treatment | < 50 | 717 | 30 | 818 | 30 |
| SBP pre-treatment | 50 - 55 | 54 | 2 | 64 | 2 |
| SBP pre-treatment | 61 - 64 | 2 | 0 | 6 | 0 |
| SBP post-treatment | pooled | 1 | 1 | 1 | 1 |
| SBP post-treatment | < 50 | 2 | 1 | 1 | 1 |
| SBP post-treatment | 56 - 60 | 0 | 1 | 0 | 1 |
| DBP pre-treatment | pooled | 419 | 20 | 476 | 21 |
| DBP pre-treatment | < 50 | 839 | 20 | 925 | 21 |
| DBP pre-treatment | 50 - 55 | 68 | 0 | 82 | 0 |
| DBP pre-treatment | 61 - 64 | 1 | 0 | 0 | 0 |
| DBP pre-treatment | 65+ | 3 | 0 | 5 | 0 |
| DBP post-treatment | pooled | 1 | 0 | 1 | 0 |
| DBP post-treatment | < 50 | 0 | 0 | 1 | 0 |
| DBP post-treatment | 56 - 60 | 2 | 0 | 1 | 0 |

**Table 2:**
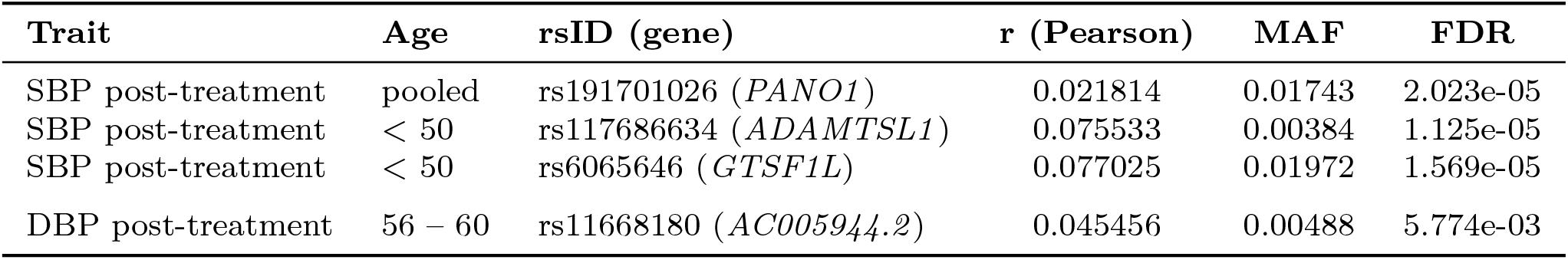
Post-treatment SBP and DBP variants detected by CI-GWAS. Listed are marginal Pearson correlation values (*r*), minor allele frequencies (MAF), and FDR-adjusted *p*-values for SNPs/LoF variants associated with post-treatment SBP/DBP across pooled and age-stratified analyzes. Variants with two or fewer carriers and SNPs not present when statins are not considered blood pressure treatment were excluded. Full results are available in Supplementary Data Tables. SNPs outside gene loci were annotated with the nearest gene. MAF was calculated within the analysed dataset.

**Fig. 3:**
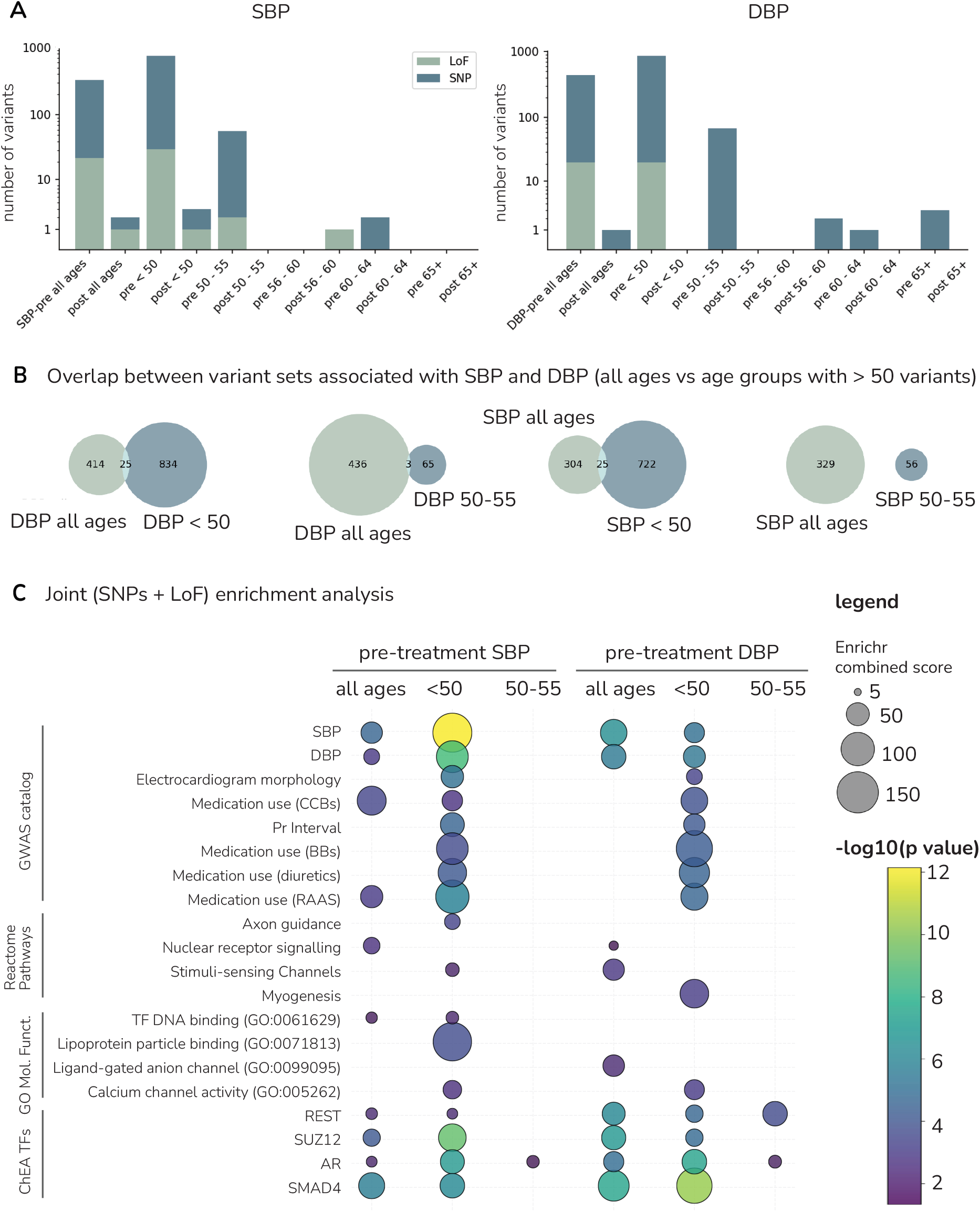
Genetic effects for systolic and diastolic blood pressure across ages. (A) Stacked barplot showing the numbers of SNPs and whole exome sequencing LoF measures discovered in the age-pooled and age-split models for systolic and diastolic blood pressure (SBP and DBP). (B) Venn diagrams displaying the overlap between variants selected in the age-pooled models (all ages) and each of the age groups with *>* 50 associated variants. SBP and DBP variants were overlapped separately. (C) Balloon plot of a joint enrichment analysis (full results available in Supplementary Data Tables), showing the top overrepresented terms in four databases. The area of the balloon is directly proportional to the product of the odds ratio and the negative natural log of the p-value. Each balloon is colored by the −*log*_10_ p-value of the enrichment test. All of the databases listed on the left have been accessed with the EnrichR tool [17]. For the GWAS catalog only cardiovascular traits have been selected.

In other EHR-based studies, statins have shown to have the highest number of proposed drug-response variants [11, 24]. Therefore, for reference, we also extracted 967,426 low-density lipoprotein cholesterol (LDL-C) measurements and analysed them within our graphical framework to investigate LDL response-associated variants. Here, across conditions, we see find associations with LDL-response (Table S3), with 15 unique variants (after removal of extremely rare variants). Many of those are consistent with well known LDL-affecting variants, including SNPs in *APOE* and LoFs of *APOB* and *LDLR*. The majority of these variants have genome-wide significant interactions with statin therapy (see Supplementary Tables) and all are also associated with pre-treatement LDL levels either in our graphical model or standard GWAS (see Supplementary Data Tables). In summary, we find more variants for LDL treatment response as compared to BP treatment response, with those for LDL being associated with both baseline and response, and those for BP being associated with treatment response only.

Apart from post-treatment BP associated variants, we find evidence for 56 and 72 genetic associations with SBP and DBP pre-treatment measures in the post 50 age groups (Figure 3, Table 1 and Methods). The majority of these are found for the measures between 50 and 55 that are conditioned on measures prior to the age of 50 and many other control variables for data structure (Figure 3, Table 1 and Methods). We find strong genetic correlations (estimated in an LD-pruned subset of the 8M SNPs) between the first and last age group of 0.929 (0.108 SE) for SBP and at 0.984 (0.116 SE) for DBP. Additionally, we find little evidence that the SNP heritability varies with age, being 0.184 (0.023 SE) and 0.180 (0.023 SE) for the first age group and 0.190 (0.019 SE) and 0.179 (0.019 SE) for the last age group, for SBP and DBP, respectively. In summary, we find evidence for genetic variation for BP that is predominantly shaped prior to the age of 50, evidence for a limited number of loci associated with age-specific phenotypic change, and indication for a few treatment response-associated variants for BP.

Associations selected by our algorithm are those that remain significantly correlated when conditioning on all other markers and traits. Conditioning on the entire genetic background of all markers is likely beneficial for accurately detecting rare to moderate frequency variant associations. However, our framework is prone to trait associations of markers that are in high LD with many other markers, simply because little variation is left after conditioning on all correlated markers. We contrasted the age-pooled and age-split analyzes for SBP, where we observe that the variant sets selected are largely non-overlapping for SBP measured prior to age 50 and age-pooled SBP measures (Figure 3B). Overall, there is a strong positive relationship between the marginal correlations of SNPs with the first age group SBP measures and of SNPs with age-pooled SBP (Figure S9). However, common variants with strong correlation with both the first age group SBP and age-pooled SBP appear to be less likely to be detected in both models, as they are in strong LD with other markers (Figure S9). This phenomenon results in a low overlap of variants selected by our algorithm between different age groups (Figure 3B, Figure S9C), despite many markers with similarly high marginal correlation values. Notably, standard approaches select mostly common variants in high LD with other markers (Figure S14).Thus, to ensure that common SNP associations are not overlooked, we also re-ran the age-pooled and age-split analyzes for SBP pre-filtering the markers by setting a maximal correlation threshold of LD *R*^2^ *>* 0.8 within 1MB and selecting a focal variant of highest frequency from each set of highly correlated markers. This serves to keep the majority of the SNP data unchanged and facilitates the selection of sets of highly correlated variables. From these data, we find a slightly larger number of common variant sets for BP across ages, but we find no change in the SNPs associated with post-treatment SBP or DBP measures (Table 1).

We assessed the replicability of our pre-treatment BP associations by estimating the regression slope of SNP correlations estimated in an independent sample of UK Biobank participants onto the SNP correlations (corrected for winner’s curse effects) used in our analyzes. A similar approach to quantify replicability has been applied in other studies [25]. As we analyse a subset of 211,845 UK Biobank individuals for whom repeated drug prescription and general practitioner measures and genotype data were available, this leaves a subset of 246,902 individuals who had only a single baseline BP measure recorded at different ages along with knowledge of whether they were on hypertension or cholesterol modifying medication at the time of measurement. We found significant regression slopes for pre-treatment SBP in age group 1 (0.82, SE 0.01, adjusted *R*^2^ = 0.73), pre-treatment DBP in age group 1 (0.80, SE 0.01, adjusted *R*^2^ = 0.78), pre-treatment age-pooled SBP (0.89, SE 0.01, adjusted *R*^2^ = 0.87), and pre-treatment age-pooled DBP (0.89, SE 0.01, adjusted *R*^2^ = 0.92), suggesting a high level of replicability of our findings.

We also analysed the p-value enrichment of the SNPs discovered in our analysis in a recent large-scale BP GWAS [20], finding an over-representation of lower than expected p-values (Figure S10). We performed gene level enrichment analyzes that included both SNPs and the genes with LoF variants with the GWAS Catalog to look for known associations with BP and other cardiovascular traits at the gene level (Figure 3C, Supplementary Note 2, Supplementary Data Tables). From all of the cardiovascular-health related traits within the GWAS catalog, the top overrepresented terms are systolic blood pressure, diastolic blood pressure, electrocardiogram features and medication use for hypertension (Figure 3C). As an example, from the 247 SBP-associated genes in the age-pooled analysis, 24 (9.7%) have been previously associated with SBP in GWAS (enrichment p-value = 1.32 ·10−5, OR = 2.90). For DBP, we find significant over-representation of genes previously associated with DBP in GWAS for the age-pooled and youngest age group DBP gene sets (age-pooled p-value = 1.67 ·10−6, OR = 3.21; age *<* 50 p-value = 1.80 ·10−6, OR = 2.60). For more details, see Supplementary Note 2. Taken together, this suggest our results are highly replicable and that the genetic variation for BP is shaped prior to mid-life, with some evidence for SNPs whose effects contribute to BP variation later in life.

Conditional on these SNP associations and all other variants genome-wide, we also discover many genes with LoF variants associated with the pre-treatment measures (Table 1). Examples of LoF variant associations (with three or more carriers within the UK Biobank) include *SMAD2*, detected in the pre-treatment SBP from all ages. This gene produces a signaling protein involved in artery remodeling [26, 27]. For DBP, in the *<* 50 age group, we replicate the previously reported association with *PDE3A*. This gene encodes a phosphodiesterase and its gain of function mutations cause brachydactyly with severe hypertension [28, 29]. Thus, we replicate known genetic syndromes associated with hypertension as well as identified novel LoF variants conditional on common variant background. In Supplementary Note 2, we provide a full discussion of our findings, with all LoF associations listed in the Supplementary Data Tables.

Finally, we note that we find hundreds of associations with potential confounders, especially with the number of observed measurement points (~ 200 SNPs and LoF variants in the age-pooled models, and ~1000 SNPs and LoF variants across the five age groups in the age-split models). These associations are described in detail in the Supplementary Data Tables and Supplementary Note 2. For CVD diagnoses made prior to the first available measurement, after conditioning on all likely confounders and pre-treatment BP values, we find four associated variants (see Supplementary Data Tables).

### Treatment-associated pharmacogenetic variants

Our framework enables the discovery of pharmacogenetic variants that have a significant association with treatment variables after controlling for the disease state. In our analyzes, we find a limited number of such variants associated with the use of statins and ARBs (Table 3 for ARBs and Table S6 for statins). As the analysis is focused on BP, the statin-associated variants likely indicate elevated LDL levels which were not modeled and are an indication for statins. Statin-related findings can be viewed as positive controls and support for the ARB variants as we indeed recover known LDL-C and statin-taking associated loci as described below.

**Table 3:**
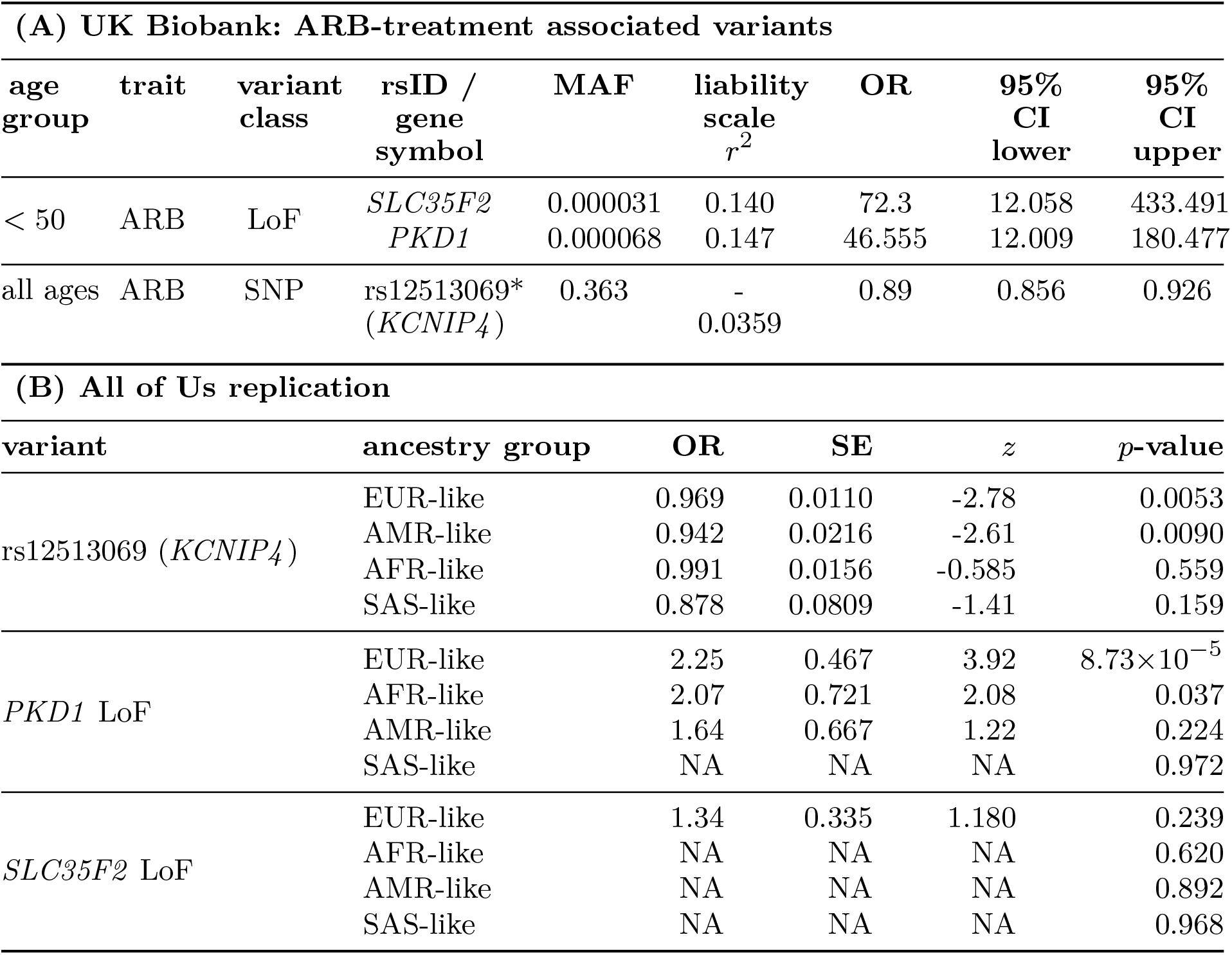
ARB-treatment associated variants and their replication in All of Us. (A) UK Biobank discovery results for ARB treatment selection (liability-scale *r*^2^, MAF, and odds ratio (OR) with 95% CI). A star indicates variants detected only in the on-drugs subset of the UKB participants (see Methods). (B) Replication in the All of Us cohort using the ever-prescribed-ARBs phenotype across 1KGP-HGDP ancestry groupings (reported as OR, SE, *z* and *p*-value). The number of individuals ever prescribed an ARB in each ancestry group: EUR-like *n* = 19,339, AFR-like *n* = 7,119, AMR-like *n* = 3,955, SAS-like *n* = 266. NAs are used if stable estimates were not obtained. MAF - minor allele frequency. ARB - angiotensin receptor blocker.

We discover two variants for ARB-taking (both LoF) and five associated with statin use (four SNPs, one LoF). When we specifically select hypertensive individuals by sub-setting our dataset to participants who received drugs at at least one time-point (Figure 4A), we uncover an additional LoF variant and eight SNPs for statins and one SNP for ARBs. In the age-split model, we discover variants only in the youngest age-group (*<* 50) and all of these are LoF variants, while in the age-pooled analysis the majority of our findings are SNPs. For a summary of these findings, see Tables 3 and S6.

**Fig. 4:**
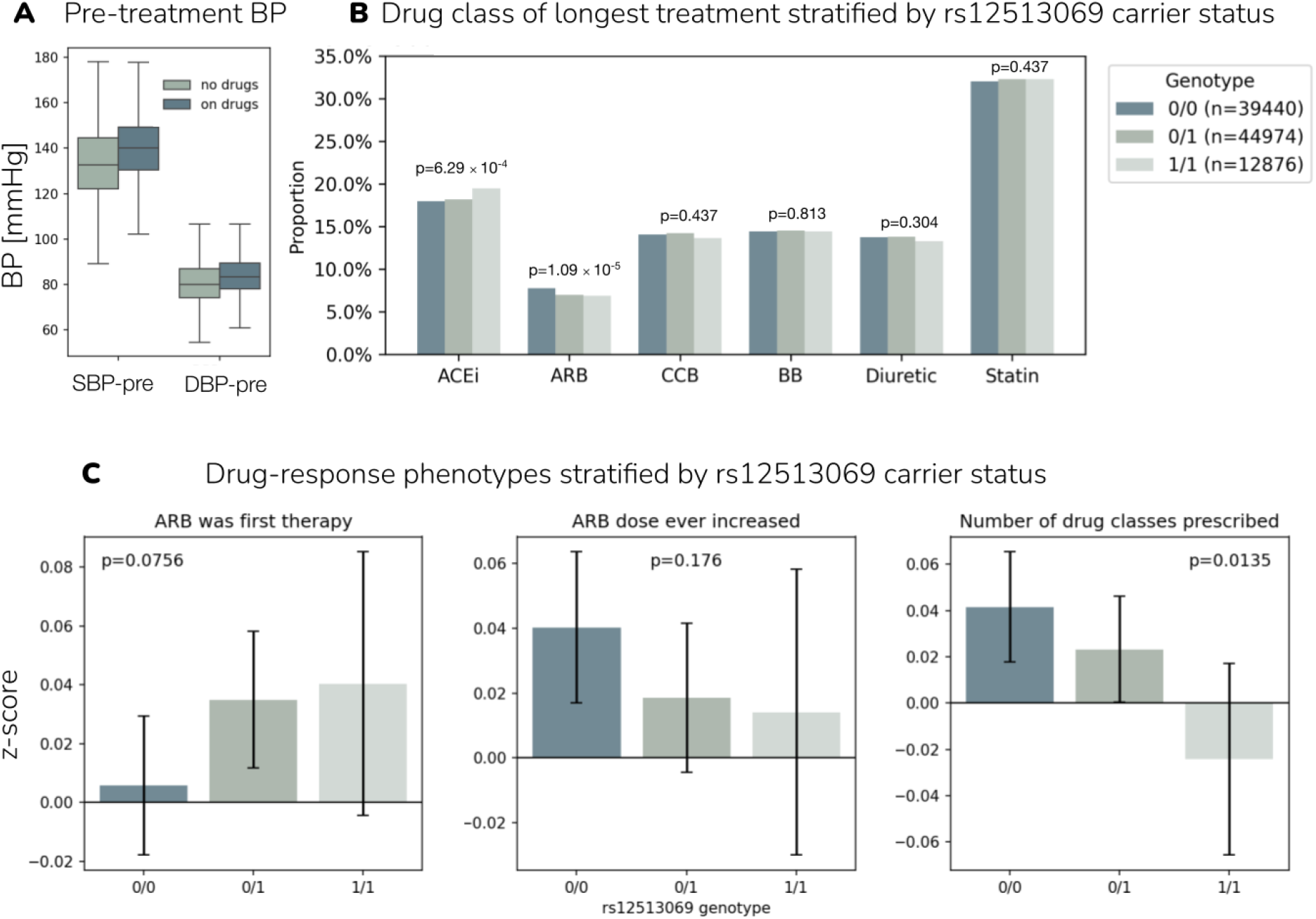
Example of ARB treatment genetics discovered in the UK Biobank. (A) Comparison of the pre-treatment systolic (SBP) and diastolic BP (DBP) measurements for UK Biobank participants with no prescriptions for any antihypertensive drugs (no drugs) versus those that were given drugs at later time points (on drugs). (B) Barplot showing which drug class was the longest treatment for each participant stratified by *KCNIP4* rs12513069 carrier status. The six drug classes we analysed are: Angiotensin-Converting Enzyme Inhibitors (ACEIs), Angiotensin Receptor Blockers (ARBs), Calcium Channel Blocker (CCBs), diuretics used in hypertension management, beta-adrenergic blockers and statins. P-values from logistic regression are reported with full results available in Supplementary Data Tables (C) Barplots (mean +/-95% CI) showing the z-score values of phenotypes associated with drug response for carriers of the SNP discovered as associated with taking ARBs. Each phenotype is adjusted for covariates and standardized to a z-score. Nominal generalized linear model p-values are noted in each of the graphs. Three phenotypes with the lowest p-values are displayed and full results are available in the Supplementary Data Tables.

Examples of associations with statin-taking include established clinically relevant variants such as SNP rs7412 in the *APOE* gene, which is associated with statin efficacy, among other phenotypes [30]. We also identify SNP rs11591147, a missense (R46L) variant in *PCSK9* with a strong reducing effect on cardiovascular risk [31]. *PCSK9* has also recently been found to be a genetic mediator of statin effects [24]. We find that carriers of the above SNPs are less likely to be treated with statins after correcting for their hypertensive status (Table S6). Among individuals in the *<* 50 age group who take statins, we identify *LDLR* LoF carriers, which serves as a positive control as *LDLR* LoF is the most common cause of familial hypercholesterolaemia [32]. All of those variants are associated with baseline LDL levels in our additional analysis where we analysed LDL directly (see Supplementary Data Tables).For each of the discovered SNPs, we searched for existing associations (at 0.8 LD value and 10−6 p-value threshold) in the Open Targets and GWAS Catalog databases. All statin-taking variants that we found have previously been reported to be associated with statin-taking or response to statins, confirming the validity of our proposed approach. All twelve statin SNPs also replicate in the publicly available summary statistics (from 500,000 individuals) of the Finnish cohort, FinnGen (see Supplementary Data Tables and Figure S11).

For ARB treatment, we find an association with Polycystin-1 (*PKD1*) LoF variants. Deleterious variants in *PKD1* cause autosomal dominant polycystic kidney disease (ADPKD). ADPKD is associated with early hypertension and loss of *PKD1* has been reported to directly impair cardiomyocyte activity and contractile function [33]. In follow-up interaction analyzes, this LoF variant has nominally significant interaction of absolute SBP change (both with and without baseline adjustment) with beta blockers, CCBs and diuretics (p-values ranging from 0.00041 to 0.025) with negative effect sizes for CCBs and diuretics. Although only suggestive, these findings indicate a possibly different response of ADPKD patients to those drugs and provide some indication why these patients are preferably treated with ARBs.

Furthermore, we find an association of LoF variants in the *SLC35F2* gene with ARB treatment. Other members of the solute carrier (SLC) gene family are well-established pharmacogenes [34]. *SLC35F2* is known to be up-regulated in tumors [35, 36]. In an animal model with a renin–angiotensin–aldosterone system (RAAS)-affecting mutation, *SLC35F2* expression was down-regulated in kidneys [37]. Notably, a closely related gene, *SLC35F3*, transports thiamine and variants in it have long been established as associated with hypertension [38]. To our knowledge, there are no reports of consequences of LoF variants in *SLC35F2* neither in humans nor in animals, and no known associations with cardiovascular traits, making it a candidate for further study. We find nine additional LoF variants for ARB-taking in the youngest age group, but exclude them from our report, as they only had a single carrier in the subset of the youngest age group that was given ARBs. Similarly for an ACEi associated LoF variant (*TSHZ1*) (see Supplementary Data Tables for full results). We additionally performed a range of sensitivity analyzes adding or removing traits and participants (see Table S4) and note that the ARB associated variants are consistently recovered, apart from a setting where age-group edges are shifted, where we recover 63.6% of those variants.

Finally, in the age-pooled analysis of hypertensive participants, we find an association between the SNP rs12513069 and ARB-taking. It is a common (minor allele frequency, MAF=0.363) intronic variant of *KCNIP4*. At a p-value threshold of 10−6, we find no existing associations in the GWAS Catalog nor in the Open Targets databases, neither for this SNP nor for any other SNP in *>* 0.8 LD with it. One GWAS study identified another SNP in *KCNIP4* (rs145489027), in LD *r*^2^ = 0.40 with rs12513069, as positively associated with ACEi-induced cough [39]. We find that carriers of rs12513069 are less likely (OR 0.89) to be treated with ARBs conditioned on their pre-treatment BP (Table 3) and their longest treatment is more likely to be with a drug from the ACEi class (OR = 1.042 for each minor allele, Figure 4B).This additionally makes *KCNIP4* a candidate for further study.

These treatment indicator associations are conditioned on pre-treatment BP, all control phenotypes, all other SNPs and LoF scores and all other drug classes. Thus, we interpret these as SNPs associated with individuals being placed onto a particular treatment as opposed to others, conditional on their BP readings. In accordance with this, in standard GWAS, none of the ARB associated variants are indicated as associated with baseline or BP change (Supplementary Data Tables).For these binary treatment indicators, we are limited in our detection ability to SNPs that explain on average more than 0.35% of the liability-scale variance of the indicator, depending on age and trait (see Table S5). As an example, in the analysis of the age stratified data, we have on average 77,120 individuals with ARB prescription indications in each age group. To have the same power for ARB as for a continuous trait with normal distribution like SBP, for which we have about 60,000 individuals with records in each age group, we would need records for 200,000 people instead, given its current low prescription rate of about 0.08 (Figure S2). However, despite this, we discover a limited number of associations with drug taking that have the potential to be clinically relevant, as they have large effect sizes or high frequency in the population.

### Replication and interpretation of the ARB pharmacogenomic candidate variants

To interpret the discovered variants in a pharmacogenomic context, we conducted two further follow-up studies. First, we sought to replicate the ARB related associations in the All of US dataset, which provides electronic health records with drug exposure information for 414,830 of its participants. We extracted the ever-prescribed-ARBs phenotype and tested for associations with either the SNP genotype or with the computed LoF burden scores. Of the three associations found in the UK Biobank (rs12513069 (*KCNIP4*), *PKD1* LoF and *SLC35F2*) we replicated two, which had smaller effect sizes but correct sign and were highly significant (Table 3B).

For rs12513069, in data on 234,353 individuals with EUR-like genetic ancestry (for a description of the All of Us cohort genetic ancestry groupings, see Methods) of which 19,339 were ever prescribed an ARB, we find a significant association (OR = 0.969, Table 3B). We also find a significant association in data on 79,105 individuals with AMR-like genetic ancestry of which 3,955 were ever prescribed an ARB (OR = 0.942, Table 3B). However, we find no evidence for an association in 84,148 individuals with AFR-like genetic ancestry of which 7,119 were ever prescribed an ARB or in 5,579 individuals with SAS-like genetic ancestry of which 266 ever prescribed an ARB. Other ancestry groups did not have sufficient ARB-prescribed individuals to conduct the analysis.

For *PKD1* LoF and the ever-prescribed-ARBs phenotype we find an association in the All of Us cohort for individuals with EUR-like genetic ancestry (OR = 2.25, Table 3B) that is weaker in individuals with AFR-like genetic ancestry (OR = 2.07, Table 3B) and individuals with AMR-like genetic ancestry (OR = 1.64, Table 3B). The association with ARB-taking and *SLC35F2* LoF had the same direction but was not significant in EUR-like participants and was not significant in all other ancestry groups.

Secondly, we performed a detailed prescription data analysis focusing on ARBs and the variants that we find to be associated with individuals being placed on these drugs as opposed to others, conditional on their pre-treatment SBP. Our dataset consists of indirect ARB-response phenotypes standardized and adjusted for covariates and includes traits such as switches to other drug classes, dose adjustments and therapy augmentations (see Figure S12, details in Supplementary Note 1). For each of the three discovered variants associated with ARB taking (rs12513069, LoF in *PKD1*, LoF in *SLC35F2*) we performed single-variant association tests (see full results in the Supplementary Data Tables), assuming an additive model. We find that for rs12513069, non-reference-allele carriers are, on average, treated longer with ACEis (Figure 4B) and prescribed drugs from less drug classes than reference-allele carriers (regression coefficient = −0.029, p-value = 0.0135, Figure 4C). To further refine this pharmacogenetic interpretation, we repeated the analysis using the WGS data from the UK Biobank. At a nominal p-value threshold of 0.05, we found further 150 variants in LD with our focal variant (rs1251306 LD *r*2 *>* 0.1) associated with the total number of drugs from different classes prescribed in the group of participants with ARB prescriptions including a short intronic indel within the *KCNIP4* locus (4-21373969-T-TGTA) with LD of 0.84 to the focal SNP and a p-value of 0.00137 (regression coefficient of −0.0374). For the LoF variant carriers, we found associations with average doses of ARBs: Irbesartan for *PKD1* LoF carriers (regression coefficient of 1.18, p-value = 0.0025) and Candesartan cilexetil for *SLC35F2* LoF carriers (regression coefficient of 2.60, p-value = 0.0021).

Finally, since the treatment indicators are conditioned on initial blood pressure, we also investigated whether running the model without BP measurements would increase the number of findings. In this analysis, only one more variant passes the detection threshold for likely causal association for the binary treatment indicators, namely rs7675300 - another SBP in *KCNIP4* with a positive liability scale squared correlation (*R*^2^ = 0.035) with ARB treatment. We also investigated if the SBP-associated variants were correlated with any of the indirect ARB-response phenotypes. We analysed the extracted ARB-response variables using multiple regression models and found that, after adjusting for the number of predictors, genetic principal components and geographical coordinates, SBP SNPs had no explanatory power regardless of whether SBP was included in the model or not (see the Supplementary Data Tables).

Thus, we conclude that there is evidence for pharmacogenetic variants that have a significant association with treatment variables even when controlling for the disease state. Despite having the power to detect clinically actionable variants, as indicated by our detection of the statin associations that are currently used in clinical practice to screen patients, we find that there may be less potential for these effects for hypertensive treatment. However, for the discovered ARB variants, we provide evidence supporting that they represent candidates for screening in terms of drug-response associations for further pharmacogenomic studies.

## Discussion

Here, we show how genetic effects on baseline biomarker values can be modeled, together with genetic effects, for both: *(i)* being placed on a particular therapy and *(ii)* the response of the biomarker to therapy. Through the analysis of a pre-post treatment study design with a time-aware repeated measures graphical model, we show that pharmacogenetic candidate SNPs and LoF variants for statin and angiotensin receptor blocker therapy and response can be identified in the UK Biobank. We replicate a large number of known statin therapy and efficacy associated variants [6, 30, 40, 41], validating our approach. We discover a novel variant in *KCNIP4* that is associated with both ARB treatment and proxy ARB treatment response traits. As this is a common variant, it could represent a clinically relevant biomarker. Another variant in *KCNIP4* is known to increase the risk of adverse drug reaction to another antihypertensive [39]. *KCNIP4* is also a potentially functionally relevant gene, associated with cardiac conduction (in Reactome Pathways) with its main function being the regulation of Kv4 potassium channels. It is hypothetically connected to ACEi-induced cough via its role in sensory nerve endings in the lungs [39]. Importantly, the associations we detect with treatment and treatment response are those that remain after controlling for hypertensive and cardiovascular disease status, alongside a host of variables correlated with data missingness, ascertainment and data structure.

We find evidence for four genetic variants that directly influence BP in response to treatment including a variant in *ADAMTSL1* previously indicated in thiazide and diuretic response [21, 23]. Recent studies suggest that the influence of common and rare genetic variants on antihypertensive drug response is relatively low but may be larger for statins [11, 12]. Previous studies have faced issues separating genetic effects for disease from genetic effects for medication usage. For example, adjusting drug response phenotypes or change scores for baseline biomarker levels has previously been widespread, but has recently been shown to introduce bias [7, 10, 11]. We note that we avoid these issues here because we do not calculate a response of change phenotype. Rather, we simply examine the variation in post-treatment values conditional on both the pre-treatment measures and the therapy allocated. We additionally show that genetic variation for BP is predominantly shaped prior to the age of 50. While we identify about a hundred independent loci associated with age-specific BP changes later in life, genetic correlations and SNP-heritability estimates were high and close to 1 across mid-to late-life, suggesting that the overall genetic architecture of BP remains largely stable across mid-to late-life, despite the presence of individual loci with age-specific effects. Taken together, our results support an emerging consistent picture of a genetic basis of baseline cardiometabolic biomarker values that is shaped in early-life, which then impacts both the therapy selected for a patient and the magnitude of change induced by the medication.

There remain a number of limitations. First, larger sample sizes are required to achieve adequate power for treatment-associated traits, which can be achieved by combining drug response GWAS from observational, EHR and RCT data as well as studies in more diverse ancestries. Our framework can easily accommodate meta-analyzed summary statistics across studies, or the model can be run for different ancestry groups. In this work, we describe a novel approach by which biobanks coupled with EHRs can contribute to pharmacogenomics research with larger sample sizes than RCTs. For systematic full meta-analyses to occur, a coordinated international effort is now required to explore and deal with the likely substantial heterogeneity between biobanks in their record linkage, prescription profiles, and ascertainment biases. Here, we have attempted to avoid the issue of heterogeneity by predominantly seeking replication in a large hold-out set of UK Biobank individuals, with additional replication in the All of Us cohort across genetic ancestry groups.

Second, this is an observational study that is both prospective and retrospective. Despite, *(i)* showing that our framework is robust to moderate data missingness, *(ii)* including a large number of variables that we hope control for missing data structure and ascertainment biases, and *(iii)* replicating our results, we cannot be certain that any of our associations are not driven by some confounding factors that we did not account for. Third, we do not observe drug responses directly and can only associate SNPs and LoFs with differences between individuals in the therapy they receive and with proxy drug response phenotypes. In our pharmacogenomics follow-up study, we see that rs12513069 carriers are generally prescribed drugs from fewer classes than non-carriers. We can suggest that patients are often prescribed drugs from multiple classes if they are non-responders, but this is only conjecture. Fourth, our approach detects genetic variants whose effects remain after conditioning on all other SNPs and may miss highly correlated common genetic variants unless these are grouped into sets prior to analysis. This is handled by simply running the model on the full and grouped data and comparing the results, which is facilitated by the computational efficiency of our algorithm.

In summary, our approach is unique for the analysis of repeated measures and can be applied to very large-scale data efficiently. We show that the power to detect variants in our algorithm is similar to a standard genome-wide association study, while controlling the FDR. Thus, we provide a robust way of detecting time-, treatment- and treatment response-specific genetic associations within large-scale biobank studies.

## Methods

### Time-aware graphical model

#### Statistical testing

We present a time-aware graphical model of genetic and trait variables developed from our recently presented CI-GWAS algorithm [16] (see Code Availability). The aim is to perform variable selection on the genetic variables, removing those which are not adjacent (partially correlated) to any trait in the learned graph. This involves testing whether each edge is absent, starting from a completely connected undirected graph. We construct hypothesis testing between two variables, *V*_*i*_ and *V*_*j*_, as follows:

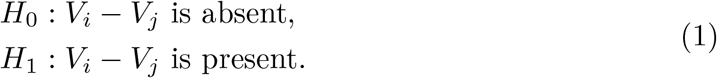

Through a series of conditional independence (CI) tests, an edge between a pair of variables is removed if the partial correlation of the two variables is insignificant given some other conditioning set of adjacent variables. In other words, there is an edge between two vertices *V*_*i*_ and *V*_*j*_ if and only if *V*_*i*_ and *V*_*j*_ are conditionally dependent given any conditioning (separation) set of *S*(*V*_*i*_) \*V*_*j*_ and *S*(*V*_*j*_)\ *V*_*i*_. Viewed as CI oracles being queried repeatedly about *V*_*i*_ and *V*_*j*_ given all possible subsets of *S*(*V*_*i*_) \*V*_*j*_ and *S*(*V*_*j*_) \*V*_*i*_, then *H*_0_ is where at least one CI oracle gives independence and *H*_1_ is where all CI oracles give dependence. Thus, the p-value of Equation 1 can be bound using

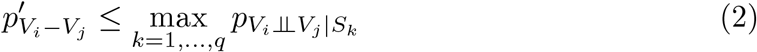

where *S*_*k*_ ⊆ {**N**(*V*_*i*_) \*V*_*j*_} or *S*_*k*_ ⊆ {**N**(*V*_*j*_) \*V*_*i*_}, consisting of other neighboring variables (**N**(·)) in the graph tested at each comparison, and *q* denotes the total number of such separation sets. If the alternative hypothesis holds for the CI tests (conditional dependence), then the null is rejected and the algorithm will not remove any of the edges between **N**(*V*_*i*_) and *V*_*i*_ as well as any of the edges between **N**(*V*_*j*_) and *V*_*j*_. Therefore, upper bounding the p-value requires simply taking the maximum of the p-values obtained for all of the CI tests for *V*_*i*_ and *V*_*j*_. As an example, suppose we measure three variables (*V*_*i*_, *V*_*j*_, *V*_*k*_) and the graphical algorithm makes two tests: *V*_*i*_ ⊥⊥ *V*_*j*_ and *V*_*i*_ ⊥⊥ *V*_*j*_ |*V*_*k*_. Suppose the p-values obtained for these tests are 5 ·10^−9^ and 5 ·10^−8^, respectively. If these p-values are less than a threshold value, then the algorithm will determine that the edge *V*_*i*_− *V*_*j*_ is present and the upper bounded p-value of this test is the maximum p-value of 5 ·10^−8^.

The algorithm conducts the tests as follows: *V*_*i*_ ⊥⊥ *V*_*j*_ |*S* is tested by first estimating the partial correlation of *V*_*i*_ and *V*_*j*_ given a separation set *S* consisting of neighbors of *V*_*i*_. We denote this by 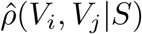. The Fisher’s z-transform of this value, *i*.*e*.

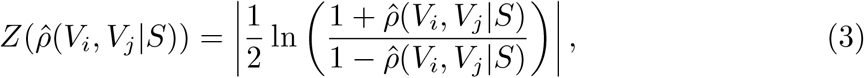

is then compared to a threshold *τ*, given by

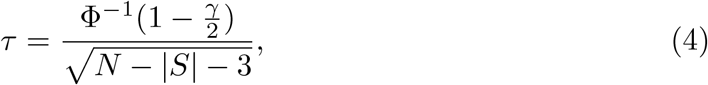

where *N* is the sample size, *γ* is a threshold applied to the test and Φ is the CDF of the standard normal distribution. 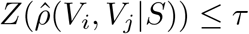 is taken to imply *V*_*i*_ ⊥⊥ *V*_*j*_ |*S*. These tests are run for consecutively increasing level *l* = |*S*|, starting with *l* = 0. The user can select an *l*_*max*_ value to limit the runtime of the algorithm. Edges between independent variables are continuously removed as the algorithm proceeds and the algorithm terminates when (*l* > *m*) ∨ (*l* > *l*_*max*_), where *m* is the number of variables.

Failure to compensate for the multiple comparisons that are made within the algorithm can result in erroneous inference. We define the false discovery rate (FDR) at threshold *α* as the expected proportion of false positives among the rejected null hypotheses

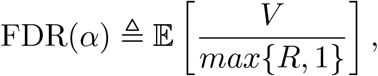

Where *V* is the number of false positives, *R* is the total number of rejected null hypotheses. This approach has been shown to provide strong control (under any form of true and false null hypotheses) of the FDR even with dependence among the *m* hypothesis tests. Benjamini and Yekutieli [42] propose

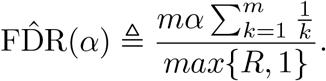

We propose applying this FDR controlling procedure (which we call “B-Y FDR”) on the edge-specific p-values after the algorithm has completed, following similar work in the statistics literature. For all kept edges, 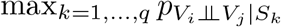 can be obtained from min 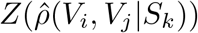. From this, a t-statistic is calculated as

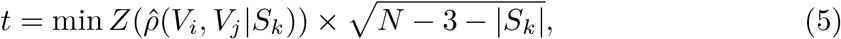

from which a p-value is obtained. The p-values from all kept edges are then corrected through the B-Y FDR correction, with the FDR set at *α* = 5%.

Our proposal builds upon previous work in the statistics literature [43]. Other work on false discovery control in similar graphical models have proposed permutation methods and adaptive FDR control [44], but these cannot be applied to high dimensional data as it requires numerous calls to the algorithm. Additionally, permutation and adaptive methods apply to parts of the graph only, rather than giving global control. As the algorithm proceeds, adaptive FDR control interferes with the variables available to form the separation sets. In our approach, the post-hoc applied multiple testing FDR correction instead ensures that the effect of errors on the final recovered graphical structure is reduced so that the FDR of the entire model is controlled [43].

### Time indexing of variables

We also introduce the use of time indices for the variables in the model which specifies the order of the variables’ measurements in time (the time index). For an edge between two variables *V*_*i*_, *V*_*j*_ that have time indices 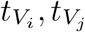, the algorithm will then not include any variable *V*_*k*_ in the conditioning set for which 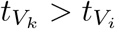 and 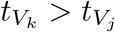. The rationale is that events in the future cannot have causal effects on events in the past. Therefore, the model will not use variables for which it is known that their measurement lies in the future to explain variation in variables in the present. Throughout, we assume that genetic variables precede all others, followed by baseline measurements, then pre-treatment measures, followed by treatment, and then post-treatment measures.

### Algorithm implementation

Our algorithm performs the conditional independence tests in a highly parallelized fashion utilizing modern GPU capabilities, enabling graphical inference for many variables in a short time. In the genomic setting, with order 10^9^ markers and order 10^2^ traits, both storage of the input correlation matrix as well as computation time required, prohibit the application of our algorithm on all variables at once. Therefore, we block the marker LD matrix in a fashion inspired by Ref. [45]. Having inferred a set of graphs with adjacency matrices *A*^*B*^ = *A*_1_, *A*_2_, …, *A*_*m*_ and separation sets *S*^*B*^ = *S*_1_, *S*_2_, …, *S*_*m*_ blockwise, we first create a merged graph with adjacency matrix *A*^∗^ following four rules: (i) a pair of traits are adjacent in *A*^∗^ if and only if they are adjacent in all *A*^*′*^ ∈ *A*^*B*^, since the presence of a single separation is sufficient to imply conditional independence; (ii) if two markers are in different blocks, they are not adjacent in *A*^∗^; (iii) if two markers are in the same block *i*, they are adjacent in *A*^∗^ if and only if they are adjacent in *A*_*i*_; and (iv) if a marker and a trait are adjacent in any block, they are adjacent in *A*^∗^. This gives a merged graph, representing a reduced set of markers that have a direct edge (partial correlation) to one or more traits in the graph. This reduces the set of variables in the graph to the ones which are most likely to have direct effects on traits. In order to prevent errors due to correlated markers being placed in separate blocks, we then repeat the algorithm on all markers selected in the first run jointly, to ensure that each marker-trait association is always tested conditional on every other potentially correlated marker.

The algorithm returns the identity of the edge kept in the graph (indicating which variables remain partially correlated conditional on others), the minimum partial correlation value for each edge, and the associated FDR corrected p-values from Eq.5.

#### Algorithm 1 Learning a time-aware graphical model

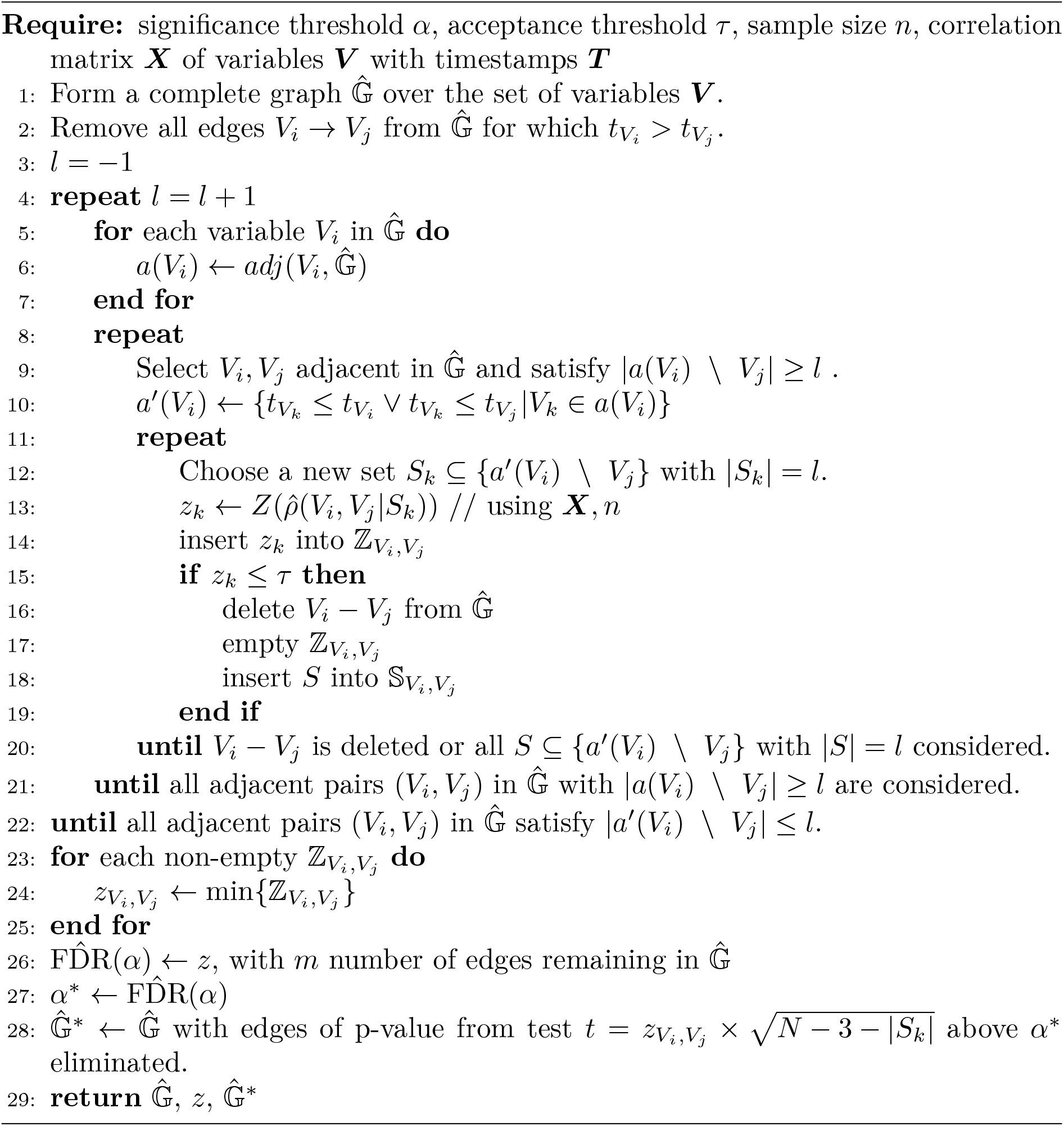

Our algorithm is described in Algorithm 1. *adj*(*V*_*i*_, 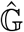) refers to the number of neighbors of variable *V*_*i*_ in 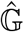. A *repeat* loop computes and stores the adjacency sets after each new conditioning set size *l*. This means that a type-II error in testing does not affect which CI tests are performed for other pairs of variables with conditioning set size *l* as all combinations are considered. It has been proved that this makes the algorithm order-independent, with tight bounding of the p-value in Eq. 2 [43]. The input required consists of ***X***, which in our case is a matrix of pair-wise correlations that the algorithm constructs from the following input correlation matrices: (i) *M*^*G*^, (ii) *M*^*GT*^ and (iii) *M*^*T*^, representing correlations between markers, between markers and traits, and between traits, respectively. These correlation matrices are prepared as follows:

- *M*^*G*^: for each input block *B*_*i*_ we obtain the LD-matrix using PLINK 1.9 with the options `--r triangle bin 4` and `--allow-no-sex`, selecting the rsIDs of the markers in the block with the `--from` and `--to` options [46].
- *M*^*GT*^ : we compute the marker-trait correlations using linear regression models. We first obtain linear regression coefficients *β*_*ij*_ for each marker *i* and trait *j* by running PLINK 2.0 on the bed file with the complete marker data and the individual-level trait-data with `--glm omit-ref --variance-standardize` options [47]. We then compute the correlation via 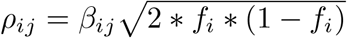, where *f*_*i*_ is the minor allele frequency of marker *i*.
- *M*^*T*^ : Here we simply compute the correlations between the individual-level data trait columns.

We calculate all correlations with the binary traits on the liability scale, as suggested by Ref. [48]. For this, elements of *M*^*GT*^ and *M*^*T*^ are populated with polyserial correlations [49] between continuous variables and binary variables, tetrachoric correlations [50] between binary variables, and Pearson correlations between continuous variables.

We provide open source code and scripts to calculate all correlations required and to run our full analysis (see Code Availability).

### Simulation studies

In our simulation study, we set up a structural equation model of a time-series of blood pressure readings and medication status, observed at two age groups. We use observed imputed SNP data from UK Biobank, described below, selecting 169,903 SNPs on chromosome 1 observed for 458,747 individuals. From these data, we construct a latent unobserved random variable *l* to reflect potential unobserved confounding factors common to population studies at baseline. We then simulate three types of trait: *(i)* pre-treatment blood pressure values; *(ii)* binary indicators of treatment with two different drugs; and *(iii)* post-treatment blood pressure values for the first age group. Individuals are then observed again at a later age group, where pre-treatment blood pressure values, binary indicators of treatment with two different drugs and post-treatment blood pressure values are also observed. Individuals who are not placed on treatment in the first age group have a pre-treatment observation in the second age group. A proportion of individuals were simulated to have stopped treatment between the two observation periods and these also have pre-treatment observations in the second age group. A proportion of individuals not receiving treatment at the second age group then go on to receive treatment and have post-treatment blood pressure observations. Individuals continuing treatment in both the first and second age groups have post-treatment measures in both age groups. A graphical presentation of the simulation design is shown in Figure 2. We use this framework to simulate three different scenarios, each with six different data ascertainment settings (no ascertainment and five ascertainment settings described below), giving 18 settings in total. For each setting, 100 replicates were run, giving a total of 1,800 data sets, each of sample size 458,747 and 169,903 SNPs.

#### Scenario 1

We first simulate a latent unobserved random variable as follows: *l* = *W*_*s*_*β*_*l*_ + *ϵ*_*l*_, with matrix *W*_*s*_ containing the observed genotypes, with one row for each of 458,747 individuals and one column for each of *m*_*c*_ = 400 randomly selected causal SNPs. Each column of *W*_*s*_ is scaled and centered to a z-score value. The effect sizes *β*_*l*_ and residuals *ϵ*_*l*_ are drawn from normal distributions: 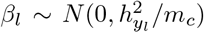, where 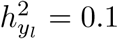, and 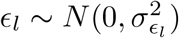, where 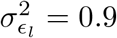.

Pre-treatment blood pressure measurements at age group 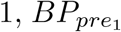, are simulated as 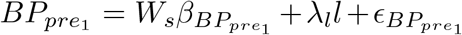, where 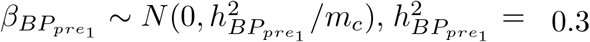, and *m*_*c*_ = 200 that are randomly selected variants that differ to those for *l. λ*_*l*_ is the effect of *l* on 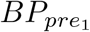, which we draw from a uniform distribution between 0.05 and 0.1. The residual variance used for simulating 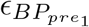 is calculated as 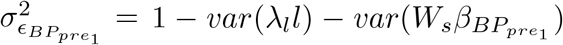. This density of causal variant allocation would give around 2,000-3,000 causal variants spread across the DNA if the simulation were to be scaled up to a genome-wide analysis. The effect sizes we simulate with *h*^2^ = 0.3 spread across 200 variants and are larger on average than would typically be observed in real data. However, this choice is important to be able to explore the TPR and FDR of our approach when simulating phenotypes. Note that below and in the main text we extensively study the theoretical and empirical detection limits (power) of our algorithm, which we show compares favorably to the power of standard GWAS association testing.

Individuals for which 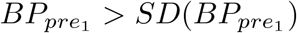, with SD the standard deviation, are selected as those with high pre-treatment blood pressure to receive treatment with either 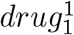 and/or 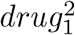. Two random vectors, 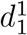 and 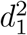 are simulated as 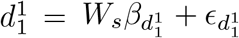 and 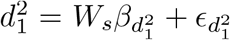, where 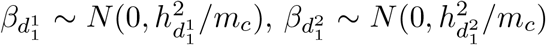 with 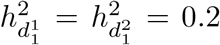, and 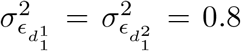, for different sets of *m*_*c*_ = 200 SNPs, randomly selected as causal variants for 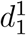 and 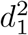,. Individuals with 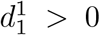 are simulated to have received 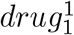 and 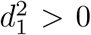 are simulated to have received 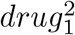. Thus, some individuals are not placed on treatment if both their 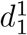 and 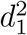 values are *<* 0, some are placed on both treatments if both their 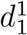 and 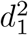 values are *>* 0, and some are placed on either 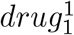 or 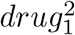 if either value of 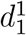 and 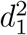 are *>* 0. This gives if either value of approximately 35,000 individuals (7-8%) on each treatment and 54,000 individuals who are simulated as being on-treatment at age group 1 who have post-treatment measures.

Post-treatment blood pressure measurements, 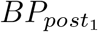, are simulated for approximately 54,000 on-treatment individuals as 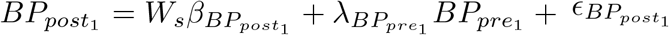, where 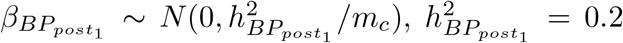; the effect of 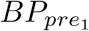 on, 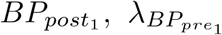 is drawn from a uniform distribution between 0.5 and 0.7; and the residual variance used to draw 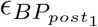 is calculated as 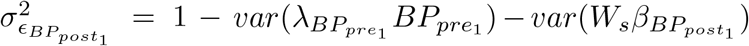. The *m*_*c*_ = 200 randomly selected causal variants differ to those for both *l* and 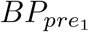. This gives a set of SNPs that influence the difference of post-treatment to pre-treatment blood pressure measures.

For the same individuals, we then simulate the same traits for a second age group. To provide a set of people that come off treatment between the age groups, we randomly select 1000 individuals whose 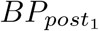 is in the lowest half of the distribution. For these individuals, and other who are not on treatment in age group 1, we sim-ulate 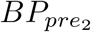as 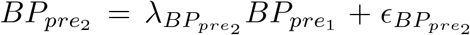, where the effect of 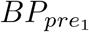 on 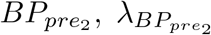, is drawn from a uniform distribution between 0.5 and 0.7. Here, there are no additional genetic effects for 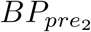. The only relationship between the SNPs and 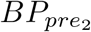 comes through 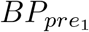. Individuals for which 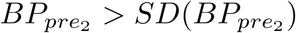 are selected as those with high pre-treatment blood pressure to receive treatment with either 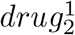 and/or 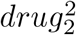at age group 2, with approximately 58,000 individuals simulated a being on treatment within each simulation replicate. We simulate the 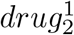 and 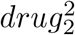 treatment in the same way as described above using the same 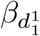 and 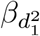 as for age 1, but different 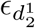 and 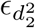. There are approximately 58,000 individuals on 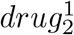 and 58,000 individuals on 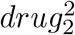, with around 100,000 individuals receiving treatment by the second age group (22% of the 458,747 individuals). Post-treatment blood pressure measurements, 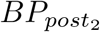, are then simulated as 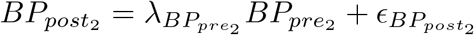, where the effect of 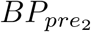 on 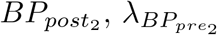, is drawn from a uniform distribution between 0.5 and 0.7. Here again, there are no additional genetic effects for 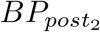, and the only relationship between the SNPs and 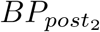 comes through 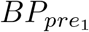.

In summary, the structural equation model is as follows:

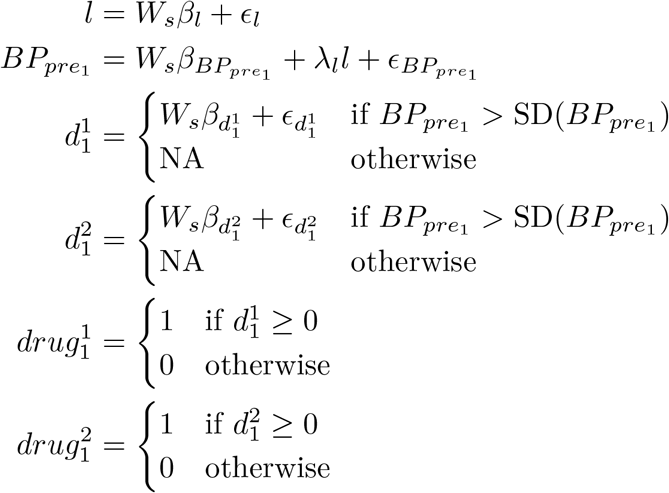

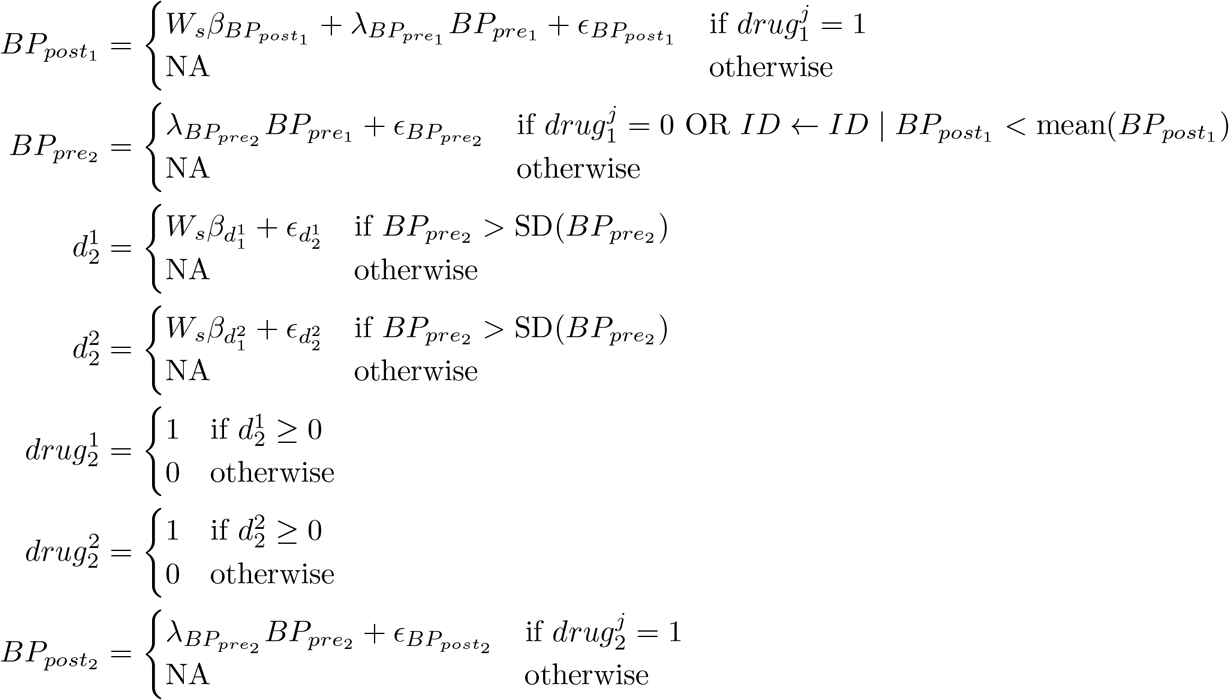

where 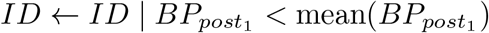 represents the sampling of 1000 individuals who come off treatment between age groups. This simulation scenario is described as the “drug response” setting as there are simulated SNP effects for 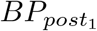.

#### Scenario 2

Our second setting repeats the first exactly, with the exception that 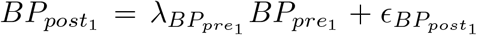 so that there is no genetic variation for blood pressure response at age 1, as there are no SNPs that influence the difference of post-treatment blood pressure to pre-treatment measures. This simulation scenario is described as the “no drug response” setting.

#### Scenario 3

Our third setting repeats the first again, but with the addition that there are genetic effects for 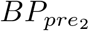 and 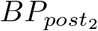 so that there are age group-specific genetic effects for both pre-treatment and post-treatment measures. We again use *m*_*c*_ = 200 randomly selected variants that differ to the other traits for both 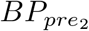 and 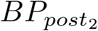. We keep the heritability of both pre-treatment and post-treatment blood pressure the same across age groups, but allow the SNP effects to differ. For pre-treatment blood pressure at age 2, we calculate the transmitted heritability as 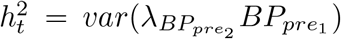 and then 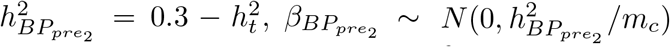 and 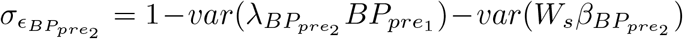. Similarly for post-treatment blood pressure at age 2, we calculate the transmitted heritability as 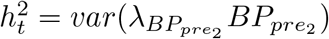 and then 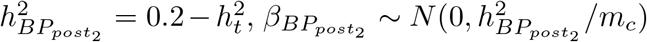 and 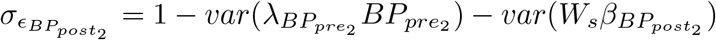. This simulation scenario is described as the “age-specific drug response” setting.

#### Censoring based on a latent selection variable

To obtain a censored dataset, we repeat all three of the simulation settings described above. We then define a threshold *frac* and remove all individuals from the dataset entirely for which *l* ≥ frac. We generate and analyzed datasets with *frac* = 0.6 (40% of individuals removed) and *frac* = 0.8 (20% of individuals removed).

#### Censoring due to decreased incentive of healthy people to go to medical check-ups

To obtain a further censored dataset, we repeat all three of the simulation settings described above. We use the values of 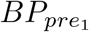 of all individuals and remove the entries of individuals for which 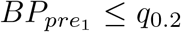, with *q*_0.2_ the lower 20% quantile with a probability 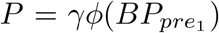, where *ϕ* is the Gaussian probability density function and *γ* is a proportionality factor (0.5 or 1.0).

#### Censoring due to death

Again, to obtain a censored dataset, we first repeat all of the simulation settings described above. At each age group *t*, we examine the values of 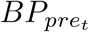 of all individuals and remove the entries of individuals for which 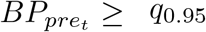, with a probability 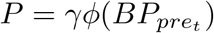, where *ϕ* is the Gaussian probability density function and *γ* is a proportionality factor which was set to 1 so that all individuals were unobserved.

### Comparison to standard treatment response models

We run Algorithm 1 over all simulation scenarios. For simulation scenarios 1-3 without censoring, we also compare the output of Algorithm 1 to previously proposed association testing approaches published in the literature to date, where drug response phenotypes are defined as either: *(i)* the absolute difference of baseline pre-treatment and post-treatment measures (|*y*_*post*_ *−y*_*pre*_ |); *(ii)* the logarithmic relative difference (*log*(*y*_*post*_) − *log*(*y*_*pre*_)); *(iii)* the proportion *y*_*post*_/*y*_*pre*_; or *(iv)* the difference adjusted for baseline levels (|*y*_*post*_ *−y*_*pre*_| *−y*_*pre*_*β*, where *β* is the regression coefficient of the effect of the baseline on the difference). For each of the 100 replicates within each of the 3 simulation scenarios, we conduct marginal association testing for each of these four drug response phenotypes using the UK Biobank chromosome 1 data observed for 458,747 individuals. We then conduct conditional and joint analysis using the COJO approach of GCTA to facilitate a comparison of the true positive rate, calculated as the proportion of simulated causal variants recovered as COJO discovered variants, and the false discovery rate, calculated as the proportion of COJO discovered variants that are not simulated causal variants.

### Detection limit calculations

We present theoretical limits of the percentage of variance in a trait explained by a marker, below which our algorithm labels associations as insignificant. We calculate these limits based on the conditional independence test, which tests the significance of the sample correlation between two variables *V*_*i*_ and *V*_*j*_, 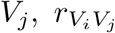, and rejects the null hypothesis 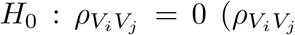 being the unobserved population correlation coefficient) if

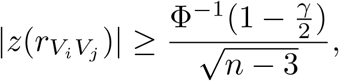

where *z* is Fisher’s z-transform of the correlation, *n* is the sample size, *γ* is the threshold applied to the test and Φ is the cumulative density function of the standard normal distribution. *n* is calculated from the standard errors of the provided correlations, *s*_*r*_, using

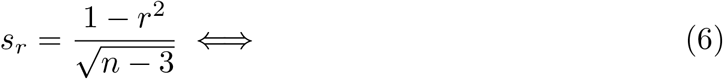

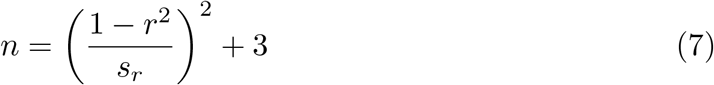

To calculate detection limits, we also use the standard errors of the correlations provided in the input, but make the approximation 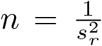, since the correlations considered are very small, such that 1 − *r*^2^ ≈ 1, and *n* − 3 ≈ *n* for large n. We then calculate the 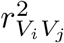 that satisfies the in the equality in the above equation, and interpret it as the variance explained in *V*_*j*_ by *V*_*i*_ at the detection limit.

To calculate the standard errors for biserial correlations, *r*_*b*_, (used for correlations between markers and binary traits), we used Hunter’s and Schmidt’s approximation [51]:

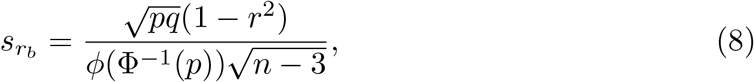

where *ϕ* is the probability density function of the standard normal distribution, *p* is the prevalence of the binary trait and *q* = 1 − *p*.

As the sample size is calculated from the standard error as stated above (using the standard error to sample size relation of the product-moment correlation), the resulting estimated sample size 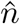 is strictly smaller than *n* in Equation 8. This reduction in the effective sample size is a consequence of the information loss resulting from the dichotomization of the liability-scale distribution of the binary trait, and has been shown to be effective in controlling the number of false positive test results caused by treating binary variable with low prevalence as Gaussian [48].

To calculate the *n* needed to get an effective sample size 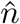 (and the corresponding detection limit) for a binary trait with prevalence *p*, we set 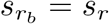 using Equations 6 and 8 and the approximations 1 − *r*^2^ ≈ 1 and *n* − 3 ≈ n, and solve for *n*:

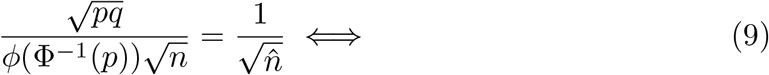

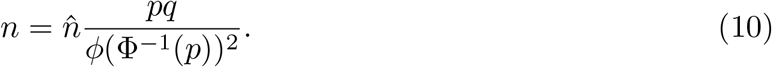

### Detection limits of genetic effects on post-treatment blood pressure

We also conducted a simulation based on the observed parameter values in the age-pooled analysis to show the relationship between the magnitude of genetic effects on the post-treatment blood pressure and their detection limit in the statistical framework used in our analysis.

We simulated data for *n* = 180, 000 individuals, which is approximately the number of individuals with non-missing records in both *ARB* and *BP*_*pre*_ in the age-pooled analysis. We simulated a Gaussian *BP*_*pre*_ and, on the basis of that, a drug indicator on the liability scale

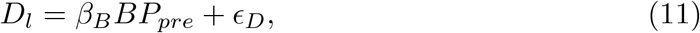

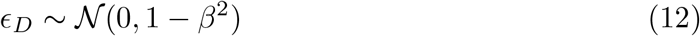

where *β*_*B*_ = 0.3, which we then dichotomized to generate a binary variable *D* with prevalence *k* = 0.05. We then calculated, for each individual *i*, the value of blood pressure after treatment:

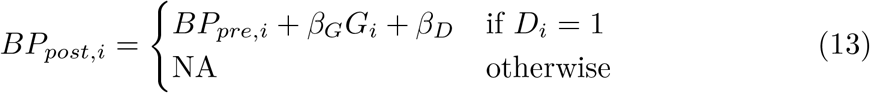

which only takes a defined value for people who have been treated (for whom *D* = 1), and differs from *BP*_*pre*_ on average by the difference *β*_*D*_ = − 0.25 plus the effect of a Gaussian genetic variable *G* of size *β*_*G*_ that we varied in the range [10^−3^, 2 · 10^−1^]. For each value of *β*_*G*_ we created 100 replicates. We then estimated the partial correlations 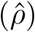 between the genetic variable *G* and post-treatment blood pressure *BP*_*post*_:

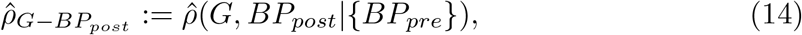

based on the Pearson correlation values calculated on pairwise complete samples. We then assessed the absolute value of the partial correlations as a function of *β*_*G*_.

### Application of the graphical model to the UK Biobank data

UK Biobank has approval from the North-West Multicenter Research Ethics Committee (MREC) to obtain and disseminate data and samples from the participants https://www.ukbiobank.ac.uk/ethics/. These ethical regulations cover the work in this study. Written informed consent was obtained from all participants.

### Phenotypic data and quality control

A detailed description of the extraction, quality control and aggregation of all blood pressure measurements (1,420,443 in the raw dataset), LDL-C measurements (967,426 in the raw dataset), associated prescriptions (1,117,900 in the raw dataset) and other related phenotypes from the UK Biobank electronic health records is available in Supplementary Note 1. All data were extracted from the UK Biobank Research Access platform with pyspark (v 3.2.3) and then processed with Hail version 0.2.132 (see Code Availability). Prescription, diagnosis and measurement codes were all recoded into unified systems and the final dataset included all collected measurements from GP and assessment centers annotated with date, source and prescription information. We collected prescriptions for most commonly used antihyptertensive and lipid-lowering drugs and grouped them into six drug categories, as given in Table S1. Unless stated otherwise, we consider the measurement to be associated with a drug if a prescription had been issued within 1 to 60 days prior to the measurement. Measurements were hard-filtered for highly improbable values and separated into pre- and post-drug treatment measures. For age-specific interrogation measures were aggregated into age groups in a way that minimized the standard deviation of the group size. For ARB response phenotypes, we also extracted dose information and prescription dates. Numerical variables were adjusted for covariates (sex, 20 genetic principal components, assessment center, genotyping batch, coordinates, measurement source) and standardized.

While it is impossible to account for all potential confounding factors in an observational study, we also collect the following data structure variables that are commonly available:

- Year of birth: Treatment patterns, health status and data collection vary over time.
- Age first and last seen in the GP records: Younger age participants have less follow-up, older participants may have missing earlier measures.
- Number of measures at each time point: Underlying health status, behavior, and clinical variation influence the patient record.
- Participant location: Treatment patterns, record collection and health status show regional variation within a study.
- Median age at measurement: This measure is used to standardize longitudinal records across time points.

We include these variables because biobank data are both prospective and retrospective: individuals enter the study at different ages and are followed over time, with information about their past being inferred from linked records. Because follow-up may be inaccurate and record linkage is imperfect, the data collected are incomplete. Both the presence/absence of an individual at specific time points and the amount of data recorded about them will reflect their underlying health status, unobserved confounding, missing data structure and time trends.

### Whole-exome sequencing data processing

The UK Biobank final release dataset of population level exome variant calls files were analyzed on the UK Biobank Research Analysis Platform (RAP) with DXJupyter-Lab Spark Cluster App (v. 2.1.1). Only biallelic sites and high quality variants were retained according to the following criteria: variant missingness *<*10%, Hardy Weinberg Equilibrium (HWE) p-value *>* 10^−15^, minimum read coverage depth of 7, at least one sample per site passing the allele balance threshold *>*0.15 (as in [52]). For genomic variants in Ensembl canonical, protein coding transcripts were annotated with the Ensembl Variant Effect Predictor (VEP) tool (see Code Availability). High-confidence loss-of-function (LoF) variants were identified with the LOFTEE plugin (v1.0.4 GRCh38). For each gene, homozygous or multiple heterozygous individuals for LoF variants received a score of 2, those with a single heterozygous LoF variant 1, and the rest 0. As genotyping data were prepared with GRCh37, position of each gene was located with the Ensembl 87 GRCh37 gtf file. Coordinates for 402 genes which were not located were lif

### Imputed genotyping data processing

We restricted our analysis to a sample of European genetic ancestry UK Biobank individuals. To infer genetic ancestry, we used both self-reported ethnic background (UK Biobank field 21000-0), selecting coding 1, and genetic ethnicity (UK Biobank field 22006-0), selecting coding 1. We then projected the 488,377 genotyped participants onto the first two genotypic principal components (PC) calculated from 2,504 individuals of the 1,000 Genomes project. Using the obtained PC loadings, we then assigned each participant to the closest 1,000 Genomes project population, selecting individuals with PC1 projection ≤ |4| and PC2 projection ≤ |3|.

Samples were also excluded based on UK Biobank quality control procedures with individuals removed that had (i) extreme heterozygosity and missing genotype outliers; (ii) a genetically inferred gender that did not match the self-reported gender; (iii) putative sex chromosome aneuploidy; (iv) exclusion from kinship inference; (v) withdrawn consent; (vi) first degree relatives. We used genotype probabilities from version 3 of the imputed autosomal genotype data provided by the UK Biobank to hard-call the single nucleotide polymorphism (SNP) genotypes for variants with an imputation quality score above 0.3. The hard-call-threshold was 0.1, setting the genotypes with probability ≤ 0.9 as missing. From the good quality markers (with missingness less than 5% and p-value for HWE test larger than 10^−6^, as determined in the set of unrelated Europeans), we selected those with minor allele frequency (MAF) ≥ 0.0002 and rs identifier, in the set of European-ancestry participants. This resulted in data set with 458,747 individuals and 8,430,446 markers (denoted as 8M dataset in the following). We also generated a dataset without highly related individuals by pruning samples with kinship *>* 0.0884 (retains third or higher-degree relatives), using the UK Biobank relatedness file (ukb rel.dat) and Hail’s maximal independent set algorithm and tie-breaker functions.

In addition, we produced an LD-pruned set of SNPs by removing markers in very high LD using the clumping approach of plink [53] (version 1.90, see Code Availability), where we ranked SNPs by minor allele frequency and then selected the highest-MAF SNPs from any set of markers with LD R^2^ ≤ 0.8 within a 1-Mb window. This results in the selection of a tagging set of variants, with only variants in very high LD with the tag SNPs removed. These filters resulted in a data set with 2,174,071 markers (denoted as 2M dataset in the following).

### UK Biobank algorithm input parameters

The input data for the algorithm were prepared as follows: First, we selected the intersection of individuals with records in the extracted phenotypic data and records in the UK Biobank. We then joined the LoF variants together with either the genetic variant data from the 8M or the 2M datasets. For these two sets of variants we computed LD-blocks, as described previously [16], using:

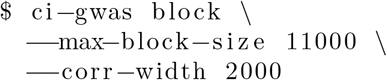

For each of the resulting blocks, we calculate the ‘mxm’ input component (correlations between genotypic variables) using plink with:

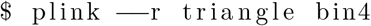

For the ‘mxp’ portion of the CI-GWAS input (correlations between genotypic variables and traits), we calculated, for every genetic variant, the correlation with each phenotype. Here, we used either the biserial correlation coefficient [49], for binary phenotypes, and the Pearson correlation coefficient for continuous phenotypes, together with the corresponding standard errors. For each correlation value, we used only the subset of individuals with records for the corresponding genetic variant and phenotype. The CI-GWAS software provides the functionality to create GWAS summary statistics of the required input format (see Code Availability).

For the ‘pxp’ portion of the CI-GWAS input (correlations between traits), we calculated the correlation between all pairs of phenotypes. Again, for each pair, we used only the subset of individuals with records in both phenotypes. We calculated the biserial correlation coefficient for pairs of continuous and binary traits, the tetrachoric correlation coefficient [50] for pairs of binary traits, and the Pearson correlation coefficient for pairs of continuous traits, with the corresponding standard errors. Unless stated explicitly otherwise, we set the p-value acceptance threshold for the individual CI-GWAS tests to *α* = 10^−3^. Note that by using biserial and tetrachoric correlations we are placing binary trait on the liability scale, thus facilitating comparisons that are independent of prevalence.

Table 4 shows the time indices used for the analysis of the simulated and real data, respectively.

**Table 4:** Time indices of variables. For (a) our simulation study, and (b) our analysis of the UK Biobank, the following time indices were provided to Algorithm 1.

| Variable | Time Index |
| --- | --- |
| (a) Simulation study |  |
| SNPs | 0 |
| $BP_{pre_1}$ | 1 |
| $drug_1^1$ | 2 |
| $drug_1^2$ | 2 |
| $BP_{post_1}$ | 3 |
| $BP_{pre_2}$ | 4 |
| $drug_2^1$ | 5 |
| $drug_2^2$ | 5 |
| $BP_{post_2}$ | 6 |
| (b) UK Biobank analysis |  |
| SNPs | 0 |
| year of birth | 1 |
| age first seen | 1 |
| age last seen | 1 |
| BP pre-treatment in age group $t$ | $3t - 2$ |
| BP post-treatment in age group $t$ | $3t$ |
| treatment in age group $t$ | $3t - 1$ |
| mean number of measurements in age group $t$ | $3t - 2$ |
| median age in age group $t$ | $3t - 2$ |

### Benchmarking within the UK Biobank data

To benchmark the results from the graphical model, we performed GWAS analyses with REGENIE (v4.2.1). A pruned set of common markers (UK Biobank v3 imputed data clumped with Plink 1.9 to *r*^2^ ≤ 0.05) was used in Step 1 to generate LOCO predictions. For binary phenotypes (drug classes and prior cvd, see Supplementary Note 1), REGENIE step 2 was run with the −−firth −−approx −−pThresh 0.000001 option. We ran three types of GWAS with REGENIE:

1. Baseline-association models: Here for each pre-treatment phenotype, we included both covariates and possible confounders (e.g. median number of measurements) as covariates in GWAS. This part was performed for both age-pooled and age-split models.
2. Analyses that aim to compare to existing PGx literature e.g. [11]: Here, we looked for variants associated with biomarker change (SBP, DBP or LDL) from pre-to post-treatment (in both age-pooled and age-split approaches) by associating the difference in the biomarker value from pre-to post-treatment in two settings: (a) including the baseline value as a covariate and (b) without adjusting for the baseline.
3. Followup interaction analysis for the age-pooled approach which tested all variants for interaction between being prescribed a drug from a given class and and treatment response defined in the same two ways as above.

*·*

A p-value of 5 · 10^−8^ was used as genome-wide significance threshold for GWAS results. For comparisons between methods for each variant, we extracted the corresponding test statistics directly for that variant.

### Replication, sensitivity and enrichment analyses

To evaluate sensitivity of the graphical model to differences in data preparation, we performed a set of additional analyses using the same subset of individuals in the UK Biobank as for our main analysis, but aggregated with different parameters: excluding statins, excluding related individuals, including prior CVD diagnosis in the models, splitting age-groups using different edges, splitting into four age groups and assuming a different time-window for associating measurements with drugs (see Supplementary Note 1 for description of their preparation).

To replicate our pre-treatment associated variant results, we used a hold-out set of data within the UK Biobank. In our main CI-GWAS analysis, we used a subset of 211,845 UK Biobank individuals where repeated drug prescription and general practitioner measures and genotype data were available. This left a subset of 246,902 individuals who had only a single baseline assessment center BP measure recorded at different ages alongside knowledge of whether they were on hypertension or cholesterol modifying medication at the time of measurement, which we used for replication. We assessed the replicability of our BP associations by estimating the regression slope of SNP correlations estimated in the independent sample of UK Biobank participants onto the SNP correlations (corrected for winner’s curse effects) from our main analysis. A similar approach to quantify replicability has been applied in other studies [25]. We controlled for winner’s curse using a conditional likelihood method for obtaining bias-reduced association estimates [54].

We additionally mapped all of our discovered SNPs to other SNPs in high LD (*R*^2^ *>* 0.8) using the available UKB LD matrices from the imputed genotype data (see Code Availability). We downloaded summary statistics from SBP/DBP GWAS [20] and for each of our SNPs assigned the lowest p-value from all its LD partners. We also parsed all of the Open Targets (including FinnGen results) and GWAS catalog records (access date: March 2025) to extract any existing associations at the p-value threshold of 10^−6^ for all of the discovered SNPs and their LD partners (see Code Availability). For the enrichment analyses, SNPs were mapped to genes based on their rsIDs with Ensembl VEP (Ensembl version 115) and joined with genes with found LoF variants into unified gene lists. All of the enrichment analyses were performed with the Enrichr tool [17] and included the following databases: GWAS catalog (gene-level), Reactome Pathways Database, Gene Ontology (GO) Molecular Function and ChEA and ENCODE consensus TFs. For the main analysis and visualization, we only included traits from the GWAS catalog that contained any of the following strings: systolic, diastolic, medication, cardio, heart, myocardial, renin, electrocardiogram, Pr Interval, diuretics, infraction. Full results are available in the Supplementary Data Tables. Traits with p-values *<* 0.05 were excluded. For visualization we present a set of terms from each database where for each of the gene lists we selected terms with the lowest p-values.

### All of Us replication analysis

For replication of the ARB-taking associated SNPs in the All of Us dataset, a cohort of participants with available drug exposure information, short-read whole genome sequencing (srWGS), EHR and age code data was selected. All of Us provide genetic ancestry groupings for all samples with srWGS data. Genetic ancestry is inferred by measuring the genetic similarity of each participant to global reference populations. The groupings of the genetic similarity categories are based on the labels used in gnomAD, the HGDP and 1000 Genomes. The following acronyms are used AFR-like (for 1KGP-HGDP-AFR-like; AFR standing for African); AMR-like (for 1KGP-HGDP-AMR-like; AMR for American); EAS-like (for 1KGP-HGDP-EAS-like; EAS for East Asian); EUR-like (for 1KGP-HGDP-EUR-like’ EUR or European); MID-like (1KGP-HGDP-MID-like; MID or Middle Eastern); SAS-like (1KGP-HGDP-SAS-likel; SAS or South Asian); and not belonging to one of the other ancestries or is an admixture (OTH or remaining individuals).

ARB ATC codes (see Table S1) have been recoded to RXUI codes via a dictionary available from SageRx, a medication ontology and medication-related data aggregator created from many different public sources of data (see Data Availability). Whole-Genome Sequences (WGS) were directly queried using Hail (v. 0.2.134-952ae203dbbe) for loci of interest in GRCh38 from the vds. For the SNP within the *KCNIP4* locus we extracted genotypes in its position chr4:21369602. The *PKD1* and *SLC35F2* genotypes were extracted for all variants within their loci (chr16:2088708-2135898 and chr11:107790991-107928293). Multiallelic variants were removed and each position and genotype was then annotated with the VEP LOFTEE tool to select pLoF variants. Only High Confidence LoF variants with call rate *>* 0.99 and less than 100 MAC were retained and aggregated into burden scores as for UKB participants. For each of the variants, a logistic regression analysis was performed within each genetic ancestry group with year of birth, sex, and all 16 genetic PCs as covariates. For the chr4:21369602, the test was conducted for the T allele effect which is the GRCh38 reference allele.

### SNP heritability and genetic correlation estimation

We used GCTA (see Code Availability) to estimate the proportion of variance attributable to a relationship matrix calculated from the 2M SNP data subset for SBP and DBP at each age group. We also used bivariate models to estimate the covariance (correlation) of the SNP effects of SBP and DBP across ages.

### Associations with drug response phenotypes

The extracted drug-response phenotypes (see Supplementary Note 1) were analyzed with linear models for their association with selected variants. Apart from variants from previously described sources we also directly analyzed Dragen WGS data for variants in LD (*R*^2^ *>* 0.1) with chr4:21369602 (both SNPs and short indels with *MAC >* 20 and call rate *>* 0.99)). Each of the drug response phenotypes was first adjusted for covariates (sex, geographical coordinates and genetic PCs) and standardized. For each of the discovered variants associated with ARB taking, we performed single-variant association tests using ordinary least squares regression. To evaluate any association of the drug-response traits with the SBP/DBP associated variants we ran multiple linear regression, where missing variant information was mean-imputed. The *R*^2^ values were adjusted for the number of predictors.

## Supplementary information

Supplementary Information accompanies this work.

## Acknowledgments

We thank members of the Robinson group at ISTA for their comments. This work was funded by an SNSF Eccellenza Grant to MRR (PCEGP3-181181), by core funding from the Institute of Science and Technology Austria, and by the SONATINA 5 National Science Center Poland grant awarded to MB (2021/40/C/NZ2/00218). We also gratefully acknowledge Poland’s high-performance infrastructure PLGrid ACK Cyfronet AGH, for providing computer facilities and support within computational grant no PLG/2021/015079. We would like to acknowledge the participants and investigators of the UK Biobank study. We gratefully acknowledge All of Us participants for their contributions, without whom this research would not have been possible. We also thank the National Institutes of Health’s All of Us Research Program for making available the participant data examined in this study. The All of Us Research Program is supported by the National Institutes of Health, Office of the Director: Regional Medical Centers: 1 OT2 OD026549; 1 OT2 OD026554; 1 OT2 OD026557; 1 OT2 OD026556; 1 OT2 OD026550; 1 OT2 OD 026552; 1 OT2 OD026553; 1 OT2 OD026548; 1 OT2 OD026551; 1 OT2 OD026555; IAA #: AOD 16037; Federally Qualified Health Centers: HHSN 263201600085U; Data and Research Center: 5 U2C OD023196; Biobank: 1 U24 OD023121; The Participant Center: U24 OD023176; Participant Technology Systems Center: 1 U24 OD023163; Communications and Engagement: 3 OT2 OD023205; 3 OT2 OD023206; and Community Partners: 1 OT2 OD025277; 3 OT2 OD025315; 1 OT2 OD025337; 1 OT2 OD025276. High-performance computing was supported by the Scientific Service Units (SSU) of IST Austria through resources provided by Scientific Computing (SciComp).

## Declarations

### Author contributions

MRR, NM and MB conceived and designed the study, conducted the analysis and wrote the paper. MRR and MK provided study oversight. JH, BB, PK, IK contributed to the analyses and the writing of the manuscript. All authors approved the final manuscript prior to submission.

### Data availability

This project uses the UK Biobank data under project number 35520. This work uses data provided by patients and collected by the NHS as part of their care and support. UK Biobank genotypic, phenotypic and linked healthcare data is available through a formal request at (http://www.ukbiobank.ac.uk). The All of Us Registered Tier Dataset is available to authorized researchers via the All of Us Researcher Workbench (https://workbench.researchallofus.org).

UK Biobank Linkage Disequilibrium Matrices are available publicly https://registry.opendata.aws/ukbb-ld.

The full set of results from graphical modeling generated by this study is available as Supplementary Data Table B.

SageRx is a medication ontology and medication-related data aggregator created from many different public sources of data https://github.com/coderxio/sagerx.

### Code availability

The CI-GWAS code is fully open source and available at https://github.com/medical-genomics-group/ci-gwas. All of the code used to prepare and analyze the data in this paper is available in a dedicated repository https://github.com/medical-genomics-group/treatment-graphical-modeling. The prescription and diagnoses recoding dictionary is available as json file within the projects’ repository https://raw.githubusercontent.com/medical-genomics-group/treatment-graphical-modeling/refs/heads/main/bp-data-extraction/recoding_dict.json.

Scripts that take a list of SNPs with their GRCh37 coordinates, extract their LD partners, and look for existing associations in GWAS Catalog and Open Targets databases) at the user-defined p-value threshold are published as a containerised stand-alone software https://github.com/ippas/ld-assoc-explorer.

R version 4.2.1 is available at https://www.r-project.org/.

Plink version 1.9 is available at https://www.cog-genomics.org/plink/1.9/.

Hail 0.2.104. is available at https://github.com/hail-is/hail.

Ensembl Variant Effect Predictor (VEP) tool is available at https://www.ensembl.org/Tools/VEP.

GCTA is available at https://yanglab.westlake.edu.cn/software/gcta/index.html.

## Supplementary Information

**Fig. S1:**
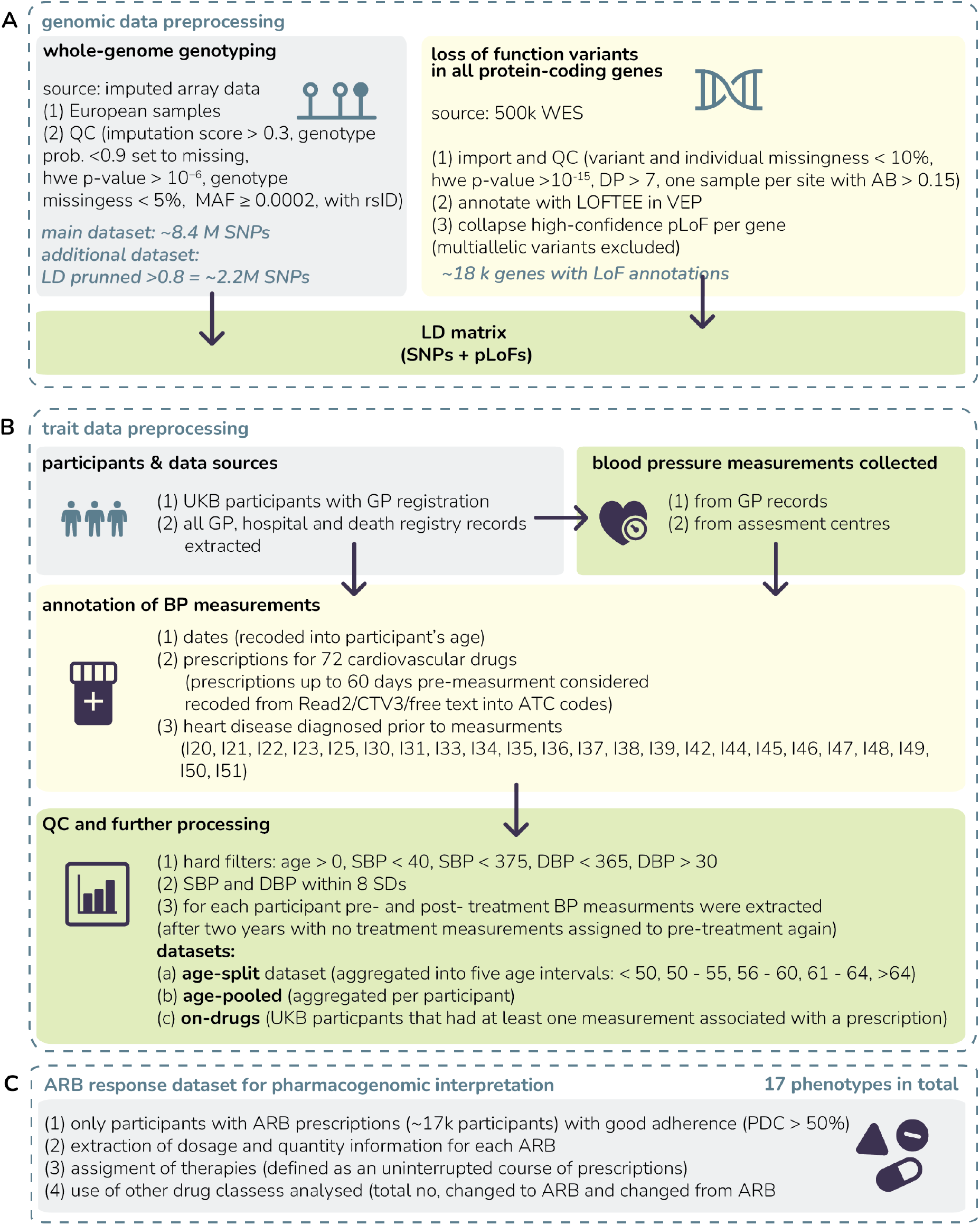
Data extraction and quality control process. (A) Imputed SNPs were included alongside whole-exome sequence loss of function variants. (B) Phenotypic values were obtained from general practitioner electronic health records and drug prescription databases, annotated and filtered. (C) A follow-up pharmacogenetic study was also conducted.

**Fig. S2:**
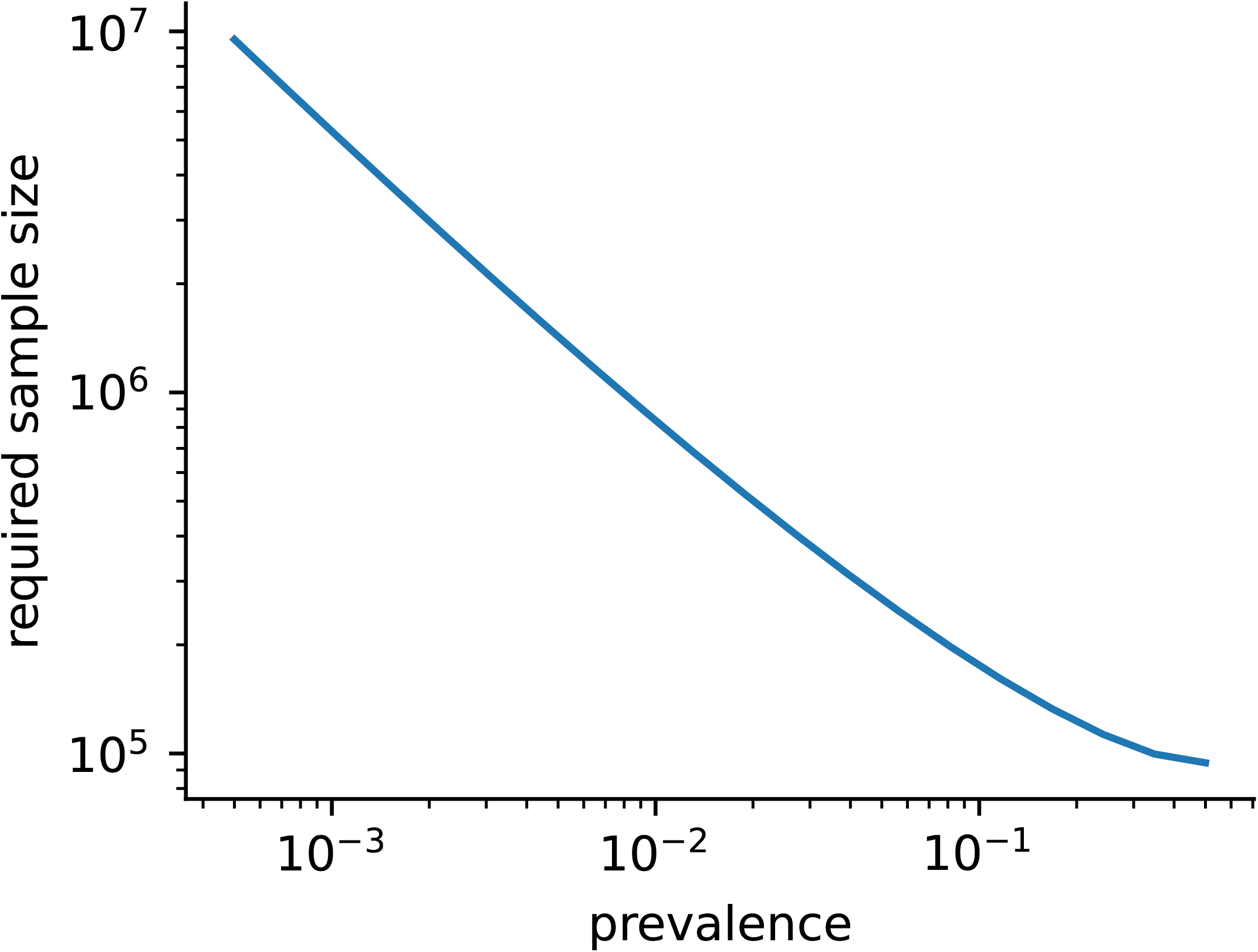
Sample size required for dichotomous trait to get same power as for continuous trait with 60k observations. Shows the the sample size required as a function of the prevalence of the dichotomous trait. See Methods for details about calculation.

**Fig. S3:**
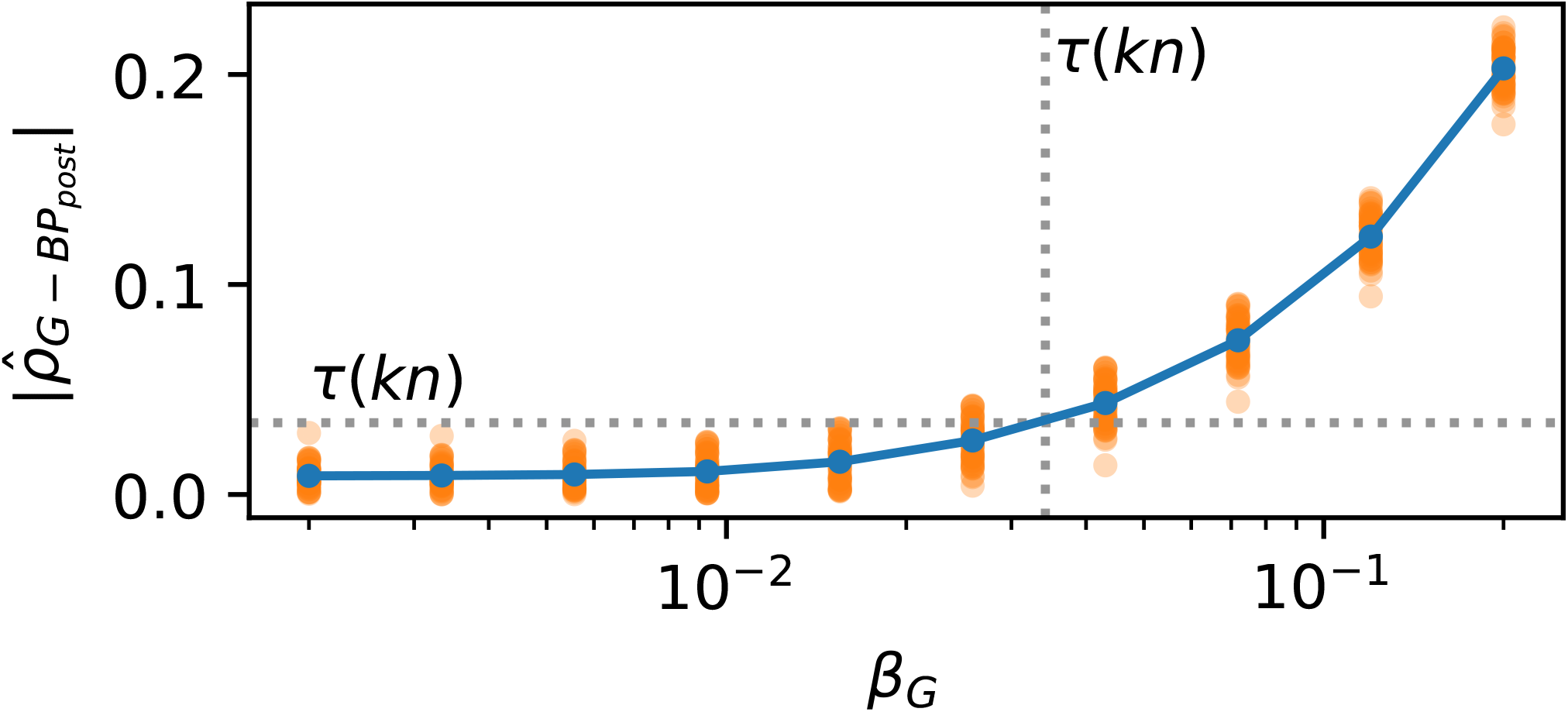
Detection thresholds of variants associated with. *BP*_*post*_ **measures**. We present the results of a simulation designed to match the correlations estimated in the analysis of real data with aged-pooled phenotypes, with *n* = 180, 000 individuals. We simulated a blood pressure trait before treatment *BP*_*pre*_, a dichotomous treatment indicator *D* with prevalence *k*, blood pressure after treatment *BP*_*post*_ (which takes a value only for treated individuals), and a genetic variable *G* with an effect *β*_*G*_ on blood pressure after treatment, using a structural equation model with parameters estimated from our real observed data (see Methods). We generated data 100 replicates for each value of *β*_*G*_. We then computed the partial correlation 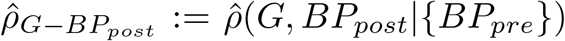, where the correlations were calculated on pair-wise complete observations. We find that, when testing 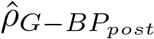 for significance at a level = 1 −03, correlations become significant (exceed *τ* (*kn*)) very closely to the marginal correlation threshold *β*_*G*_, where 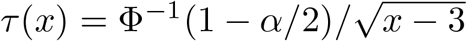. Orange dots show individual values of 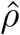, blue dots show the arithmetic mean across the 100 replicates.

**Fig. S4:**
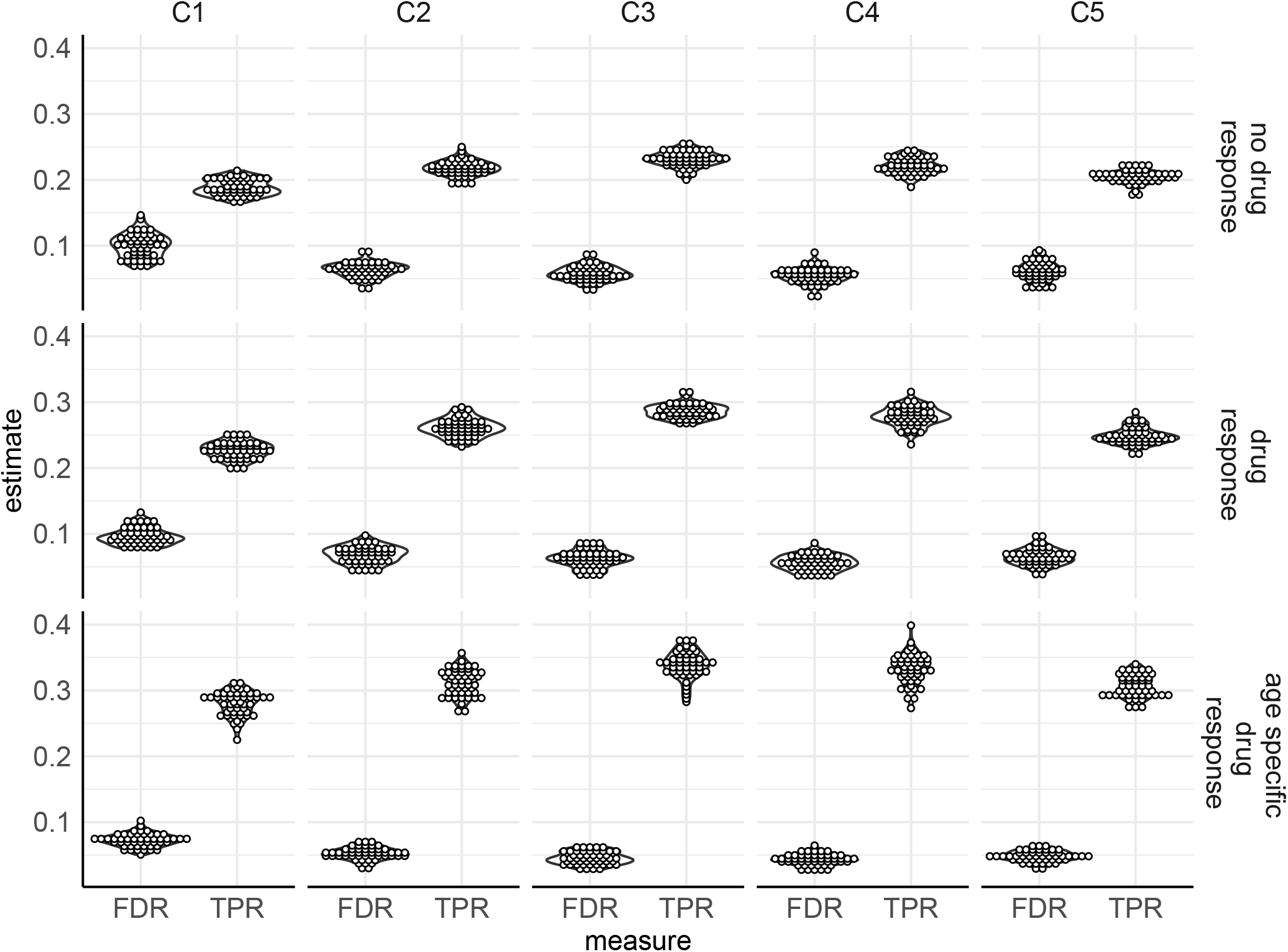
Detection of time- and trait-specific genetic effects in simulation. We simulate three main settings, shown as directed acyclic graphs in Figure 2. For each setting, we used single nucleotide polymorphism data of chromosome 1 for 458,747 individuals and 100 replicates were run. False discovery rate (FDR) and true positive rate (TPR) for the discovery of associated genetic variables for all traits when applying our graphical algorithm to the simulated data of each setting are shown. C1 and C2: censoring based on a latent selection variable, *l* ≥ frac, with *frac* = 0.6 (labeled C1) and *frac* = 0.8 (labeled C2); C3 and C4: censoring due to decreased incentive of healthy people to go to medical check-ups, where pre-treatment BP values at age 1 for which 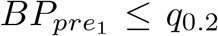, with *q*_0.2_ the lower 20% quantile with a probability 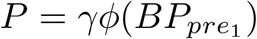, where *ϕ* is the Gaussian probability density function and *γ* is a proportionality factor (set to 1 labeled “C3” or 0.5 labeled “C4”); C5: censoring due to death at each age group *t*, we examine the pre-treatment values of 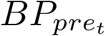 of all individuals and remove the entries of individuals for which 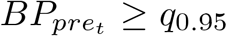. For more details, see Methods.

**Fig. S5:**
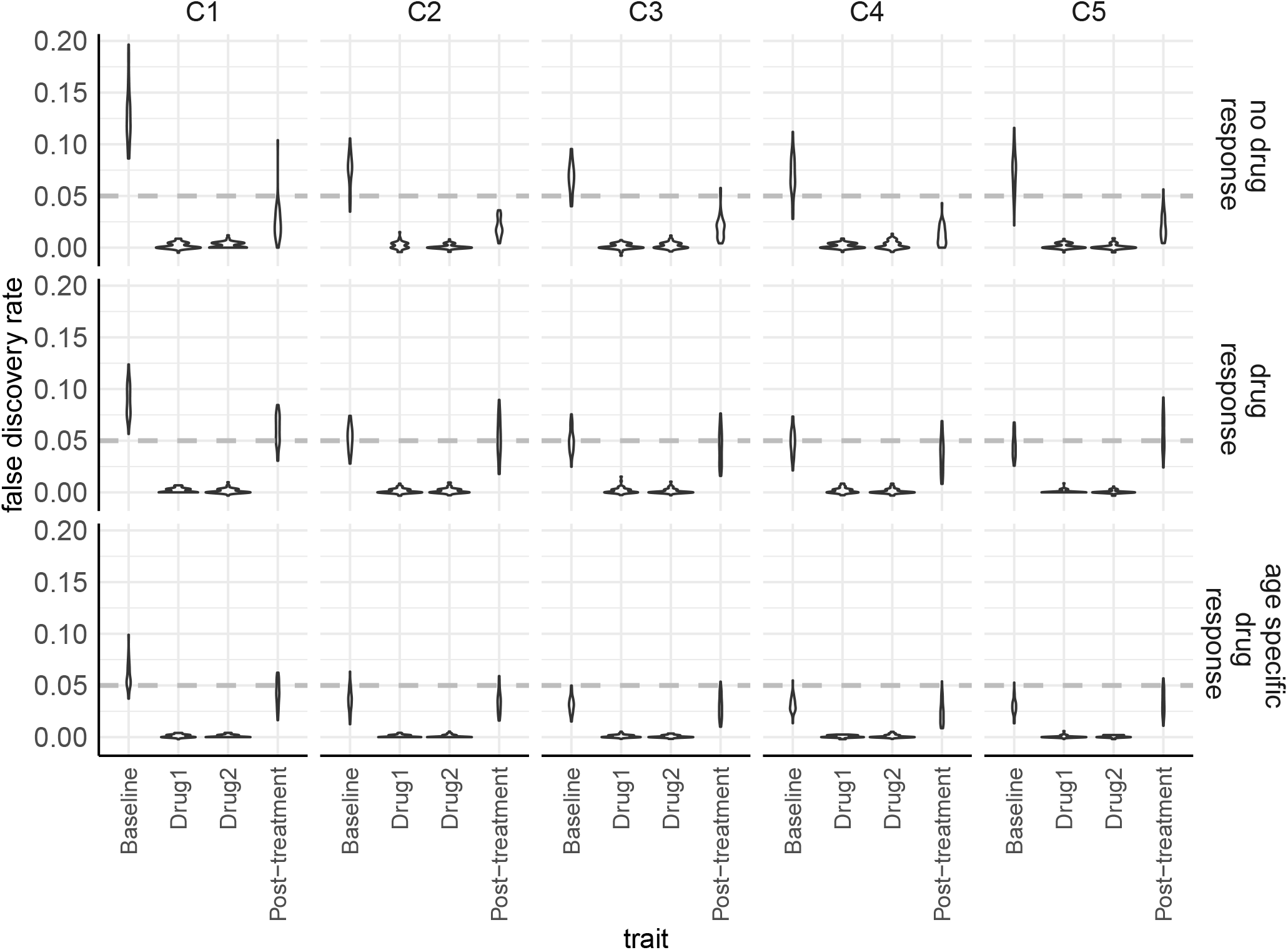
Detection of time- and trait-specific genetic effects in simulation under data ascertainment bias. We simulate three main settings, shown as directed acyclic graphs in Figure 2. For each setting, we used single nucleotide polymorphism data of chromosome 1 for 458,747 individuals and 100 replicates were run. False discovery rate (FDR) for the discovery of associated genetic variables for each trait when applying our graphical algorithm to the simulated data of each setting are shown. FDR is calculated as the proportion of associations for trait that are not simulated causal variants for that trait. Note that associations with all traits are determined jointly within a single model and that this is not the FDR of the model, which is given in Figure S4. We give these values only to aid understanding of which traits in the model are subject to elevated false discoveries under different ascertainment scenarios. C1 and C2: censoring based on a latent selection variable, *l* ≥frac, with *frac* = 0.6 (labeled C1) and *frac* = 0.8 (labeled C2); C3 and C4: censoring due to decreased incentive of healthy people to go to medical check-ups, where pre-treatment BP values at age 1 for which 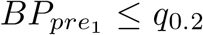, with *q*_0.2_ the lower 20% quantile with a probability 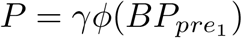, where *ϕ* is the Gaussian probability density function and *γ* is a proportionality factor (set to 1 labeled “C3” or 0.5 labeled “C4”); C5: censoring due to death at each age group *t*, we examine the pre-treatment values of 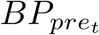 of all individuals and remove the entries of individuals for which 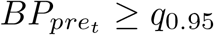. For more details, see Methods.

**Fig. S6:**
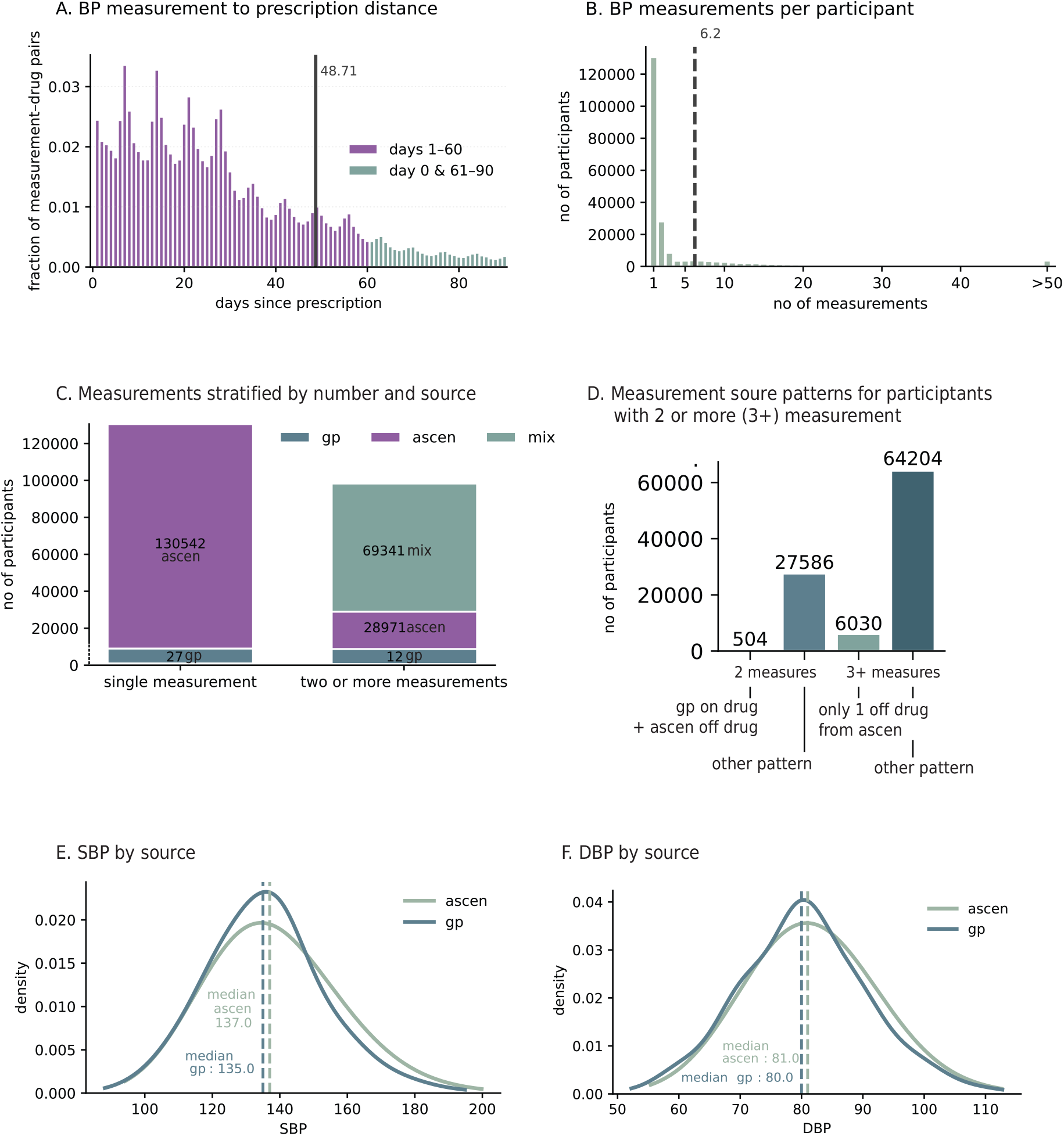
Distribution of the numbers and sources and values of collected BP measurements. **A**. Measurements associated with drugs were annotated with the days from last prescription. In violet we label measures that we consider associated with prescription. The line at 48.71 marks the average number of tablets in one prescription which led us to chose the 60-day window. All analyses have been repeated with a 30-day window for comparison (See Table S4). **B**. Number of measurements per participant, line indicates mean of 6.2. **C**. All measurements stratified by source for single (left bar) and repeated measures (right bar): ascen - assesment center, gp - general practitioners records, mix - both ascen and gp. **D**. Measurement source patterns for participants with two or more measures. Two left bars - two measures only, two right bars - three or more measures; first and third bar indicate number of participants who only have a single measurement without drugs and it is from the assessment center. **E**. and **F**. show density plots for SBP and DBP respectively stratified by source with median values labelled. ascen - assesment center, gp - general practitioners records.

**Fig. S7:**
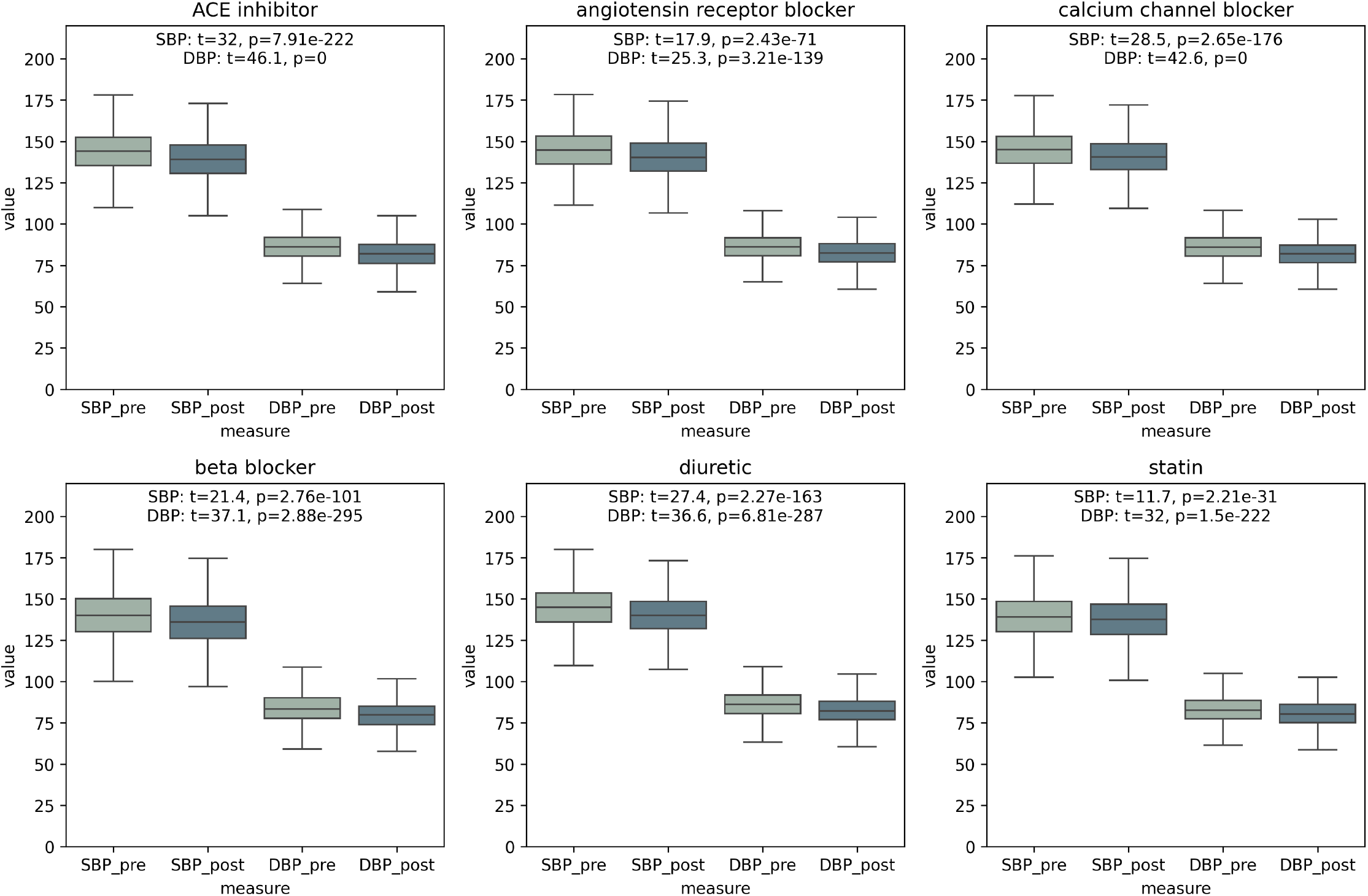
Pre- and post-treatment BP values for each of the analyzed drug classes. Boxplots are plotted for all UK Biobank participants with general practitioner records that received drugs from each class. Each of the pre- and post-treatment pairs were compared with t-tests t statistic and p-values are given on each subplot for SBP and DBP.

**Fig. S8:**
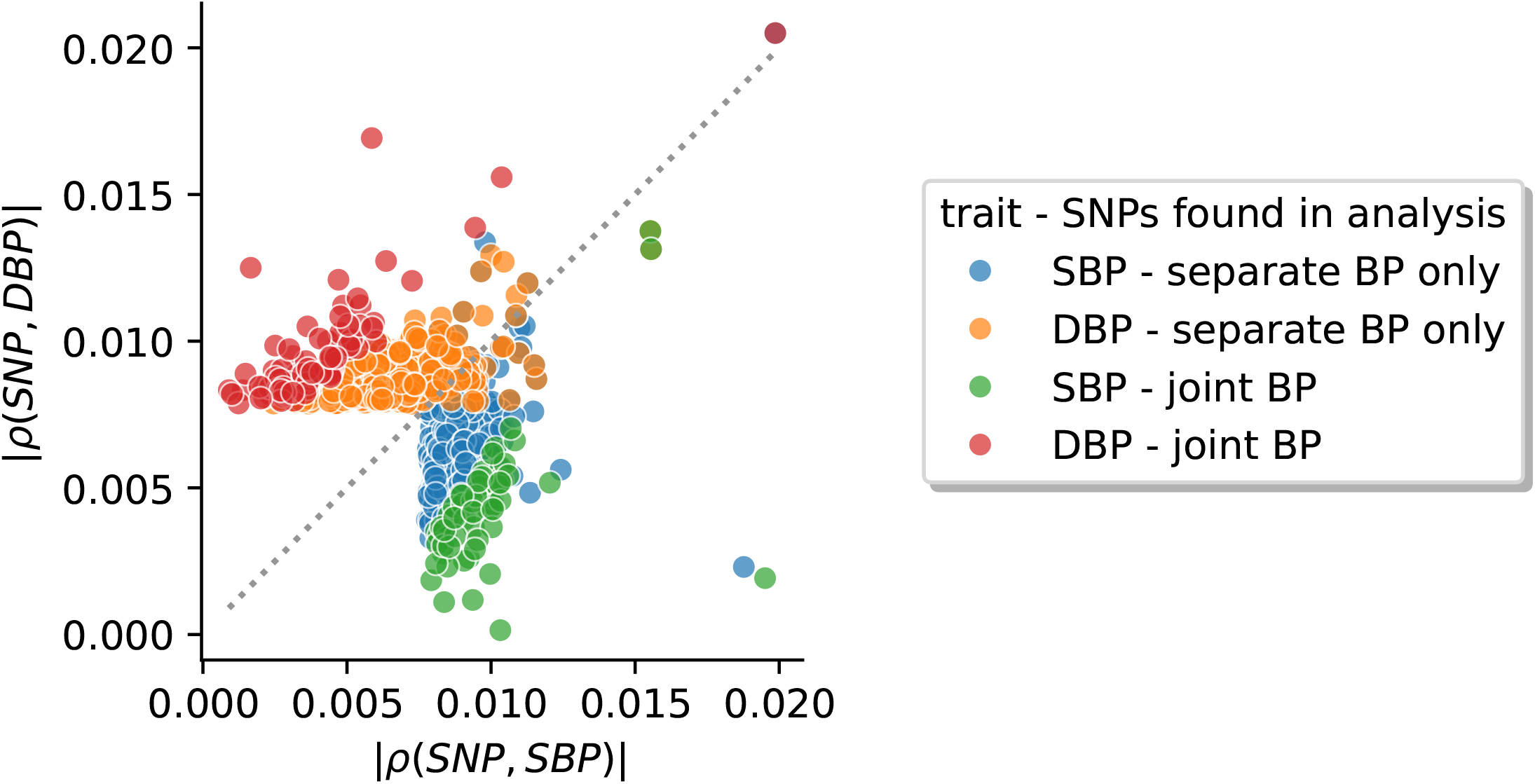
Correlations of diastolic and systolic blood pressure with markers recovered in joint and separate analyses. We inferred causal markers for systolic (SBP) and diastolic blood pressure (DBP) in three analyses: (i) including only SBP, drugs and controls, (ii) including DBP, drugs and controls and (iii) including DBP, SBP, drugs and controls. Here we show that markers found to be associated with SBP (blue, green) or DBP (orange, red) are found in the joint analysis only when their correlation with SBP is not too similar to their correlation with DBP, or else they are found only in the separate analyses.

**Fig. S9:**
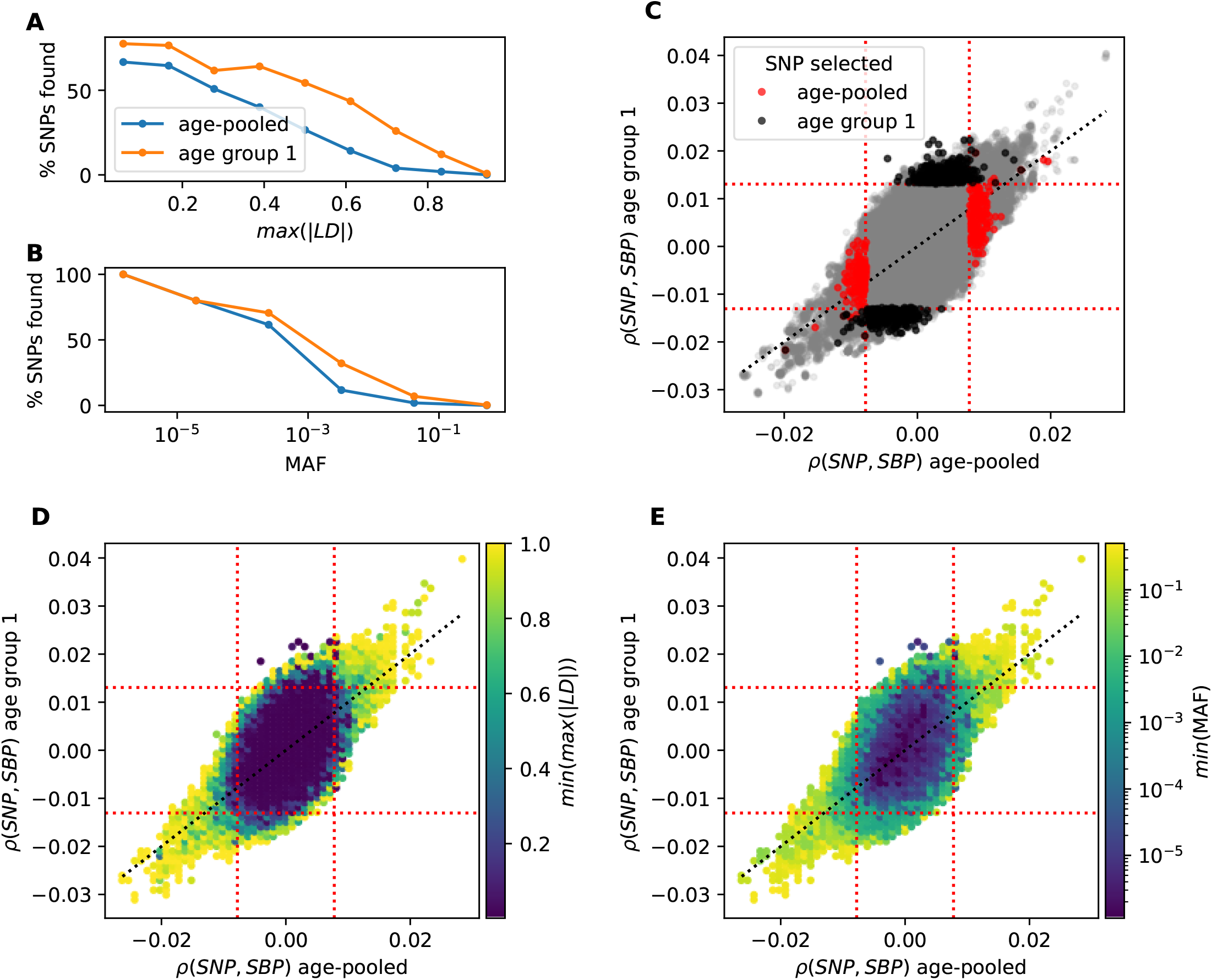
Effects of linkage disequilibrium (LD) and minor allele frequency (MAF) on the discovery of marker-trait associations with CI-GWAS and the overlap of discovered markers between the age-pooled and age-split analyses. (A) Percentage of markers with marginal correlations above the detection limits of 0.008 (age-pooled) and 0.013 (age group 1) that were recovered by CI-GWAS for *SBP*_*pre*_, calculated in ten bins in the interval [0, 1] of *max*(|*LD* |), the maximum absolute value of LD that a marker is in with any other marker. (B) Percentage as in A, but calculated in bins of MAF. (C) Marginal correlations of all 8M markers with *SBP*_*pre*_ in age group 1 and *SBP*_*pre*_ in the age-pooled dataset, with SNPs selected in each analysis highlighted in black (age group 1) and red (age-pooled), non-selected markers in grey. (D) We binned the 8M markers by correlation into 80 bins in the interval [−0.04, 0.04], then in each bin we calculated the *min*(*max*(|*LD*|)) over all markers. (E) Same binning as in D, but with *min*(*MAF*) (minor allele frequency) calculated over all markers in the bin. C, D, E: Red dotted lines indicate the detection limits in each analysis. Black dotted line shows the diagonal.

**Fig. S10:**
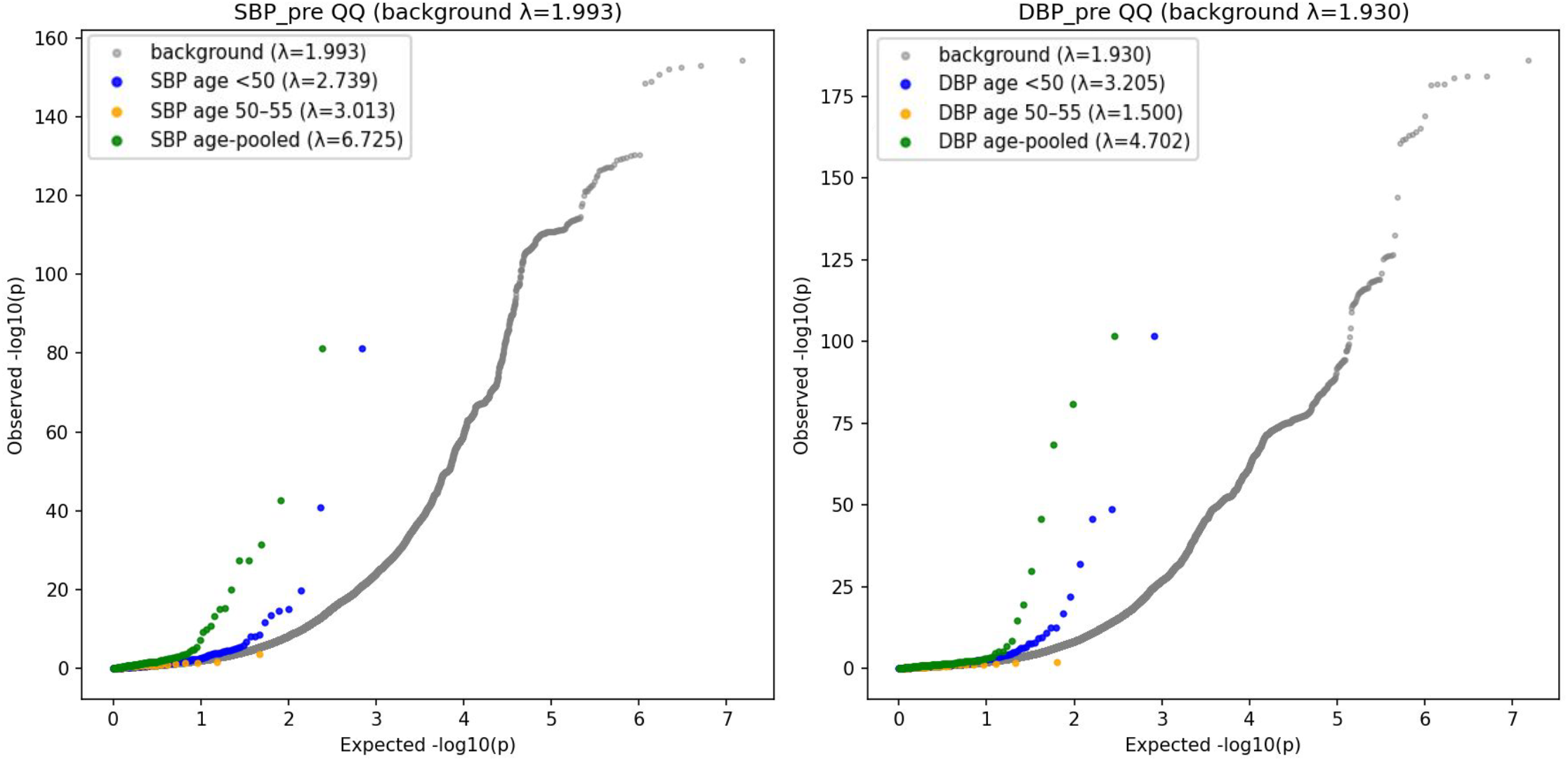
Replication of SNPs discovered in our study in a recent large-scale GWAS of blood pressure. For SNPs discovered in our analysis as well as their LD-partners (LD *r*^2^ *>* 0.8) we extracted p-values from a large-scale BP GWAS [20] and selected one representing SNP from each LD block (with the lowest p-value). The expected versus the observed −*log*_10_(*p*) values are plotted for the SNPs reported in [20] and discovered in our analysis, stratified by groups. Remaining SNPs (gray dots) include all GWAS variants after excluding our lead SNPs and their LD partners. For each list we provide the inflation factor *λ* which gives the ratio of the median chisquare statistic observed to the expected values with one degree of freedom. Higher values of *λ* indicate p-values lower than expected by chance p-values.

**Fig. S11:**
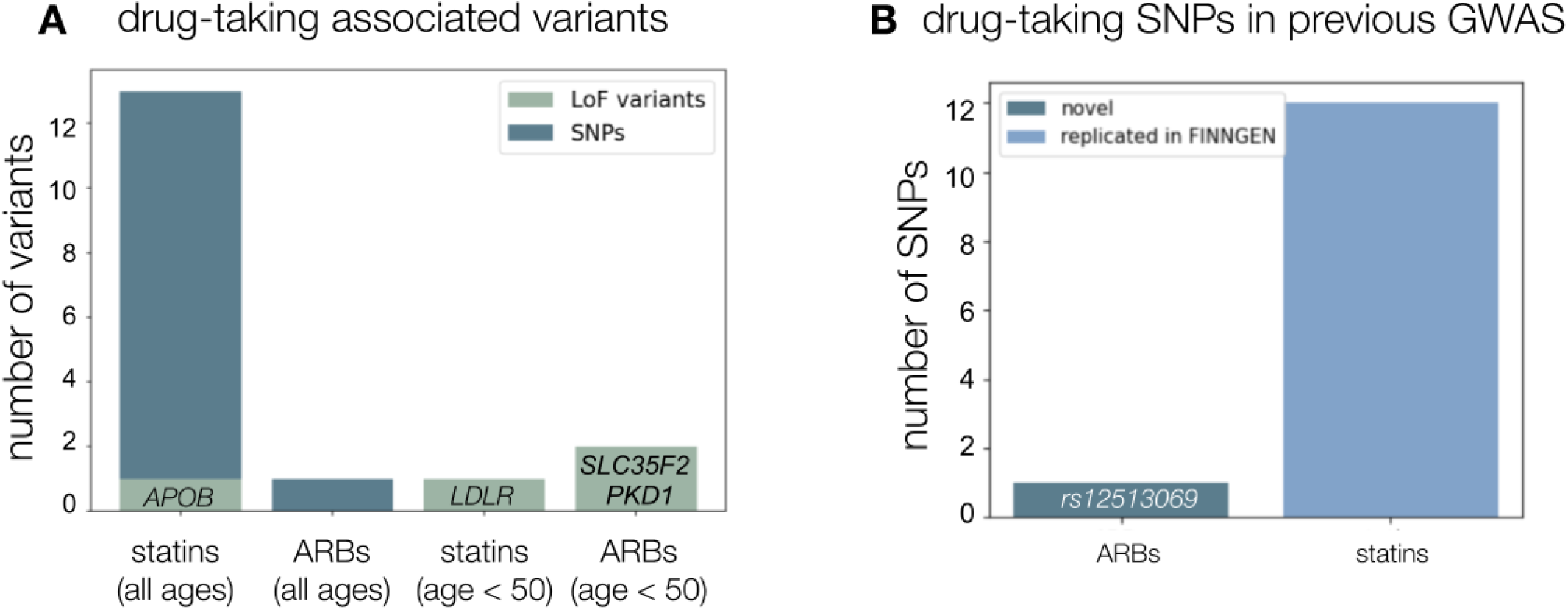
Details of treatment-selection associated genetic variants discovered in the UK Biobank. (A) Stacked barplot showing the total number of single nucleotide polymorphism (SNP) or whole exome sequence loss-of-function (LoF) variants discovered for each drug class and age groups (those with no discoveries are omitted, findings from general and on-drugs population are joined). ARB - angiotensin receptor blocker. (B) Stacked barplot indicating novel and previously found associations for SNPs. Variants that additionally replicate an external cohort (FinnGen) are marked separately.

**Fig. S12:**
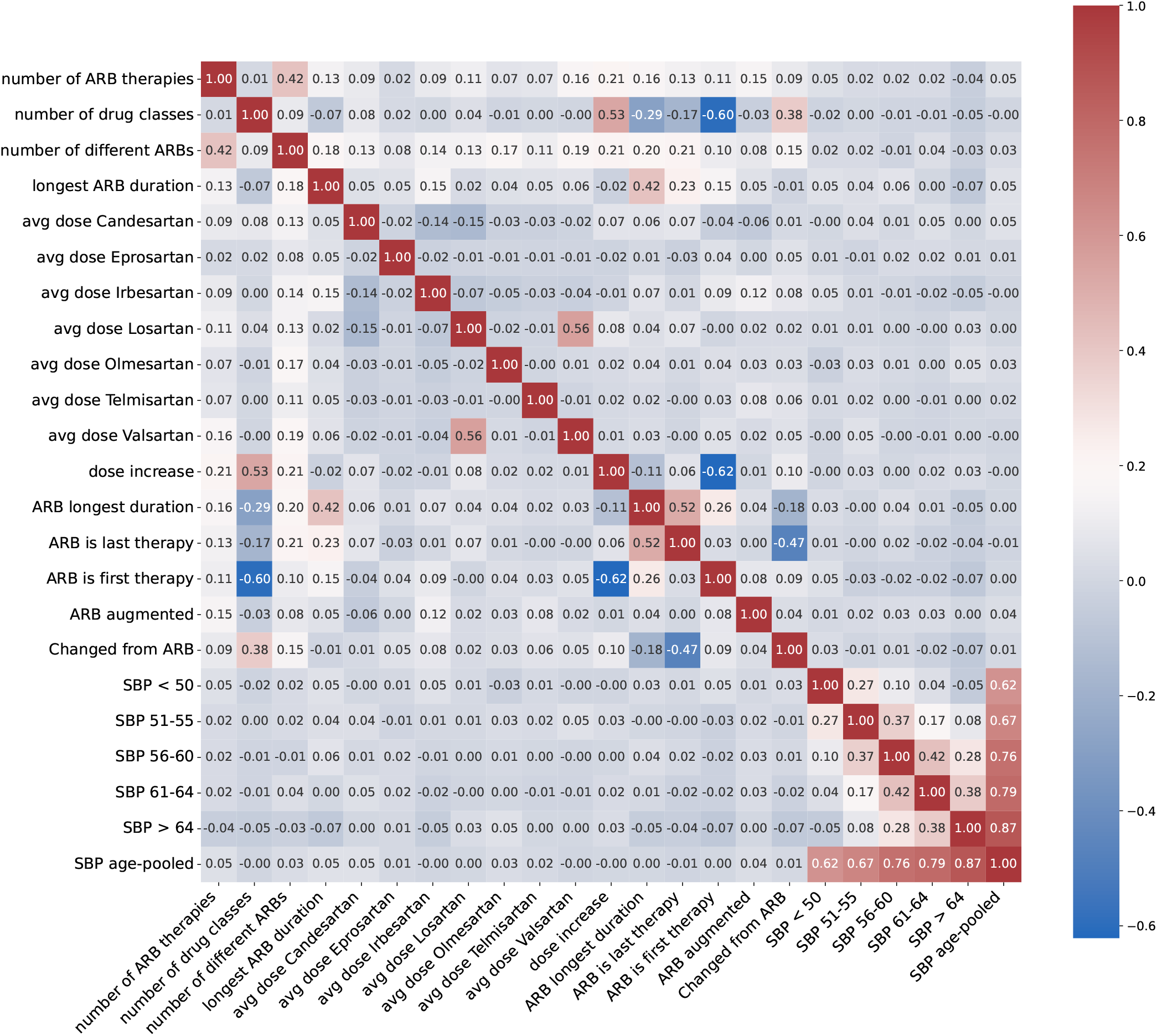
Correlation heatmap of extracted ARB-related phenotypes. See Supplementary Note 1 for details on the phenotypes. Pearson correlation values are given in each of the heatmap rectangles and colored accordingly. Pretreatment SBP values from the age-pooled and age-split datasets are also included for comparison. All phenotypes were adjusted for covariates and standardized.

**Fig. S13:**
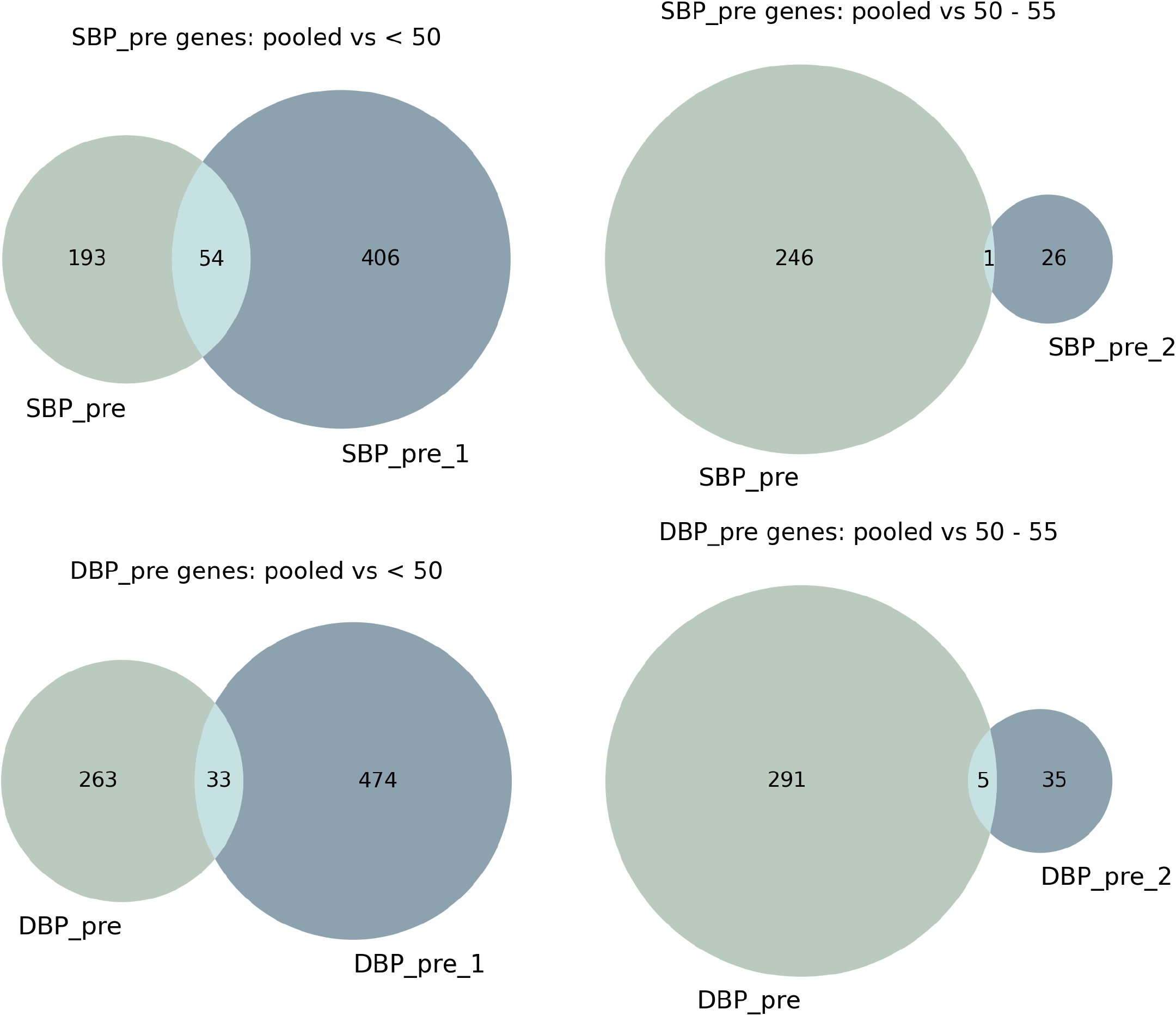
Gene-level overlap between discovered variants. SNPs were mapped to genes with Ensembl VEP and joined with genes with LoF variants. The Venn diagrams show the overlap between the age-pooled (S/DBP pre) and age-split gene lists (S/DBP pre 1/2) for DBP and SBP separately. Only lists with over 50 variants have been included.

**Fig. S14:**
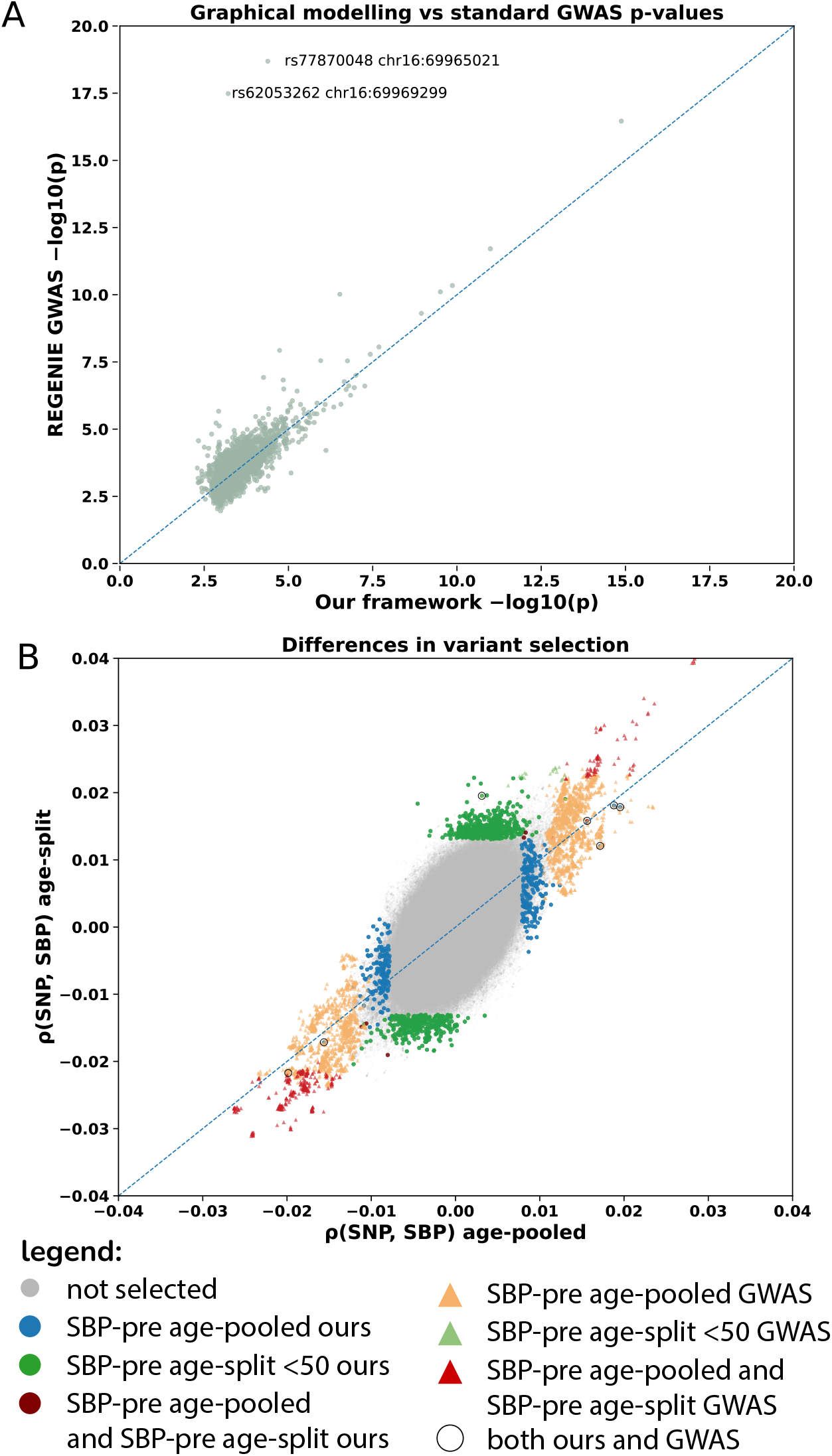
Comparison with standard methods for pre-treatment associated variants. **A**. For all variants associated with pre-treatment (baseline) SBP/DBP in both the age-pooled and age-split models (5516 variants), we plotted − log_10_(*p*) using the nominal p-values from our graphical modeling (x-axis) against those reported by REGENIE two-step regression (y-axis). **B**. Scatterplot of variant–trait correlation values. Correlations with pre-treatment SBP from the age-pooled setup are shown on the x-axis, while correlations with pre-treatment SBP in the youngest age group from the age-split model are shown on the y-axis. Grey points represent variants from the ~8M-variant dataset that were not identified as associated in any model. Blue, green, and dark red points indicate variants selected by our graphical modeling approach, while orange, red, and light green triangles indicate standard GWAS variants with *p <* 5 *×*10−8. The sets of selected variants are largely non-overlapping; variants selected by both approaches are highlighted with black circles. Standard GWAS-selected variants are mostly common variants and often show high LD with other markers (see Figure S9 for details).

**Table S1:**
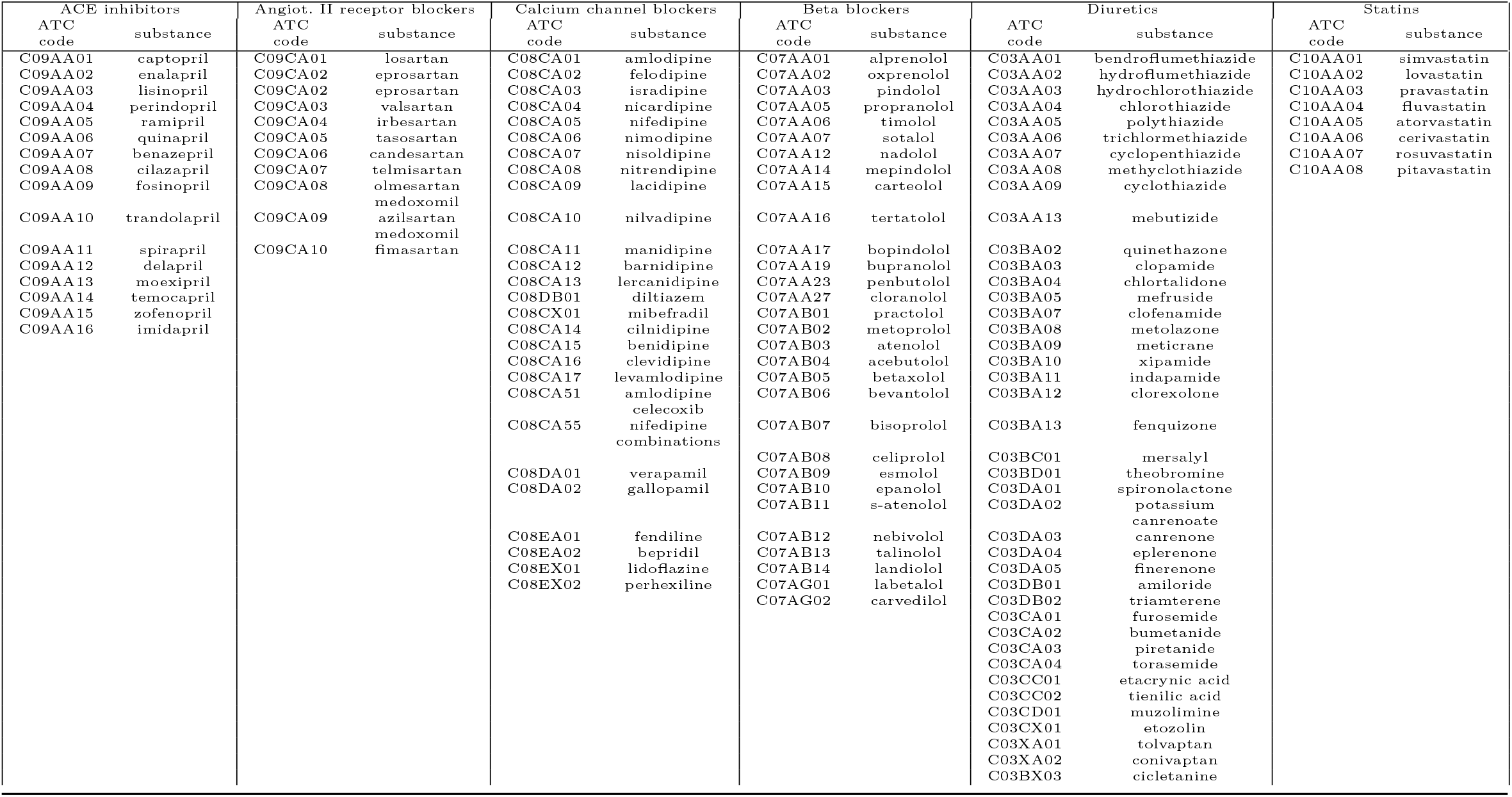
Drug substances and their classifications.

**Table S2:** Linear mixed model results for drug effects on systolic (SBP) and diastolic blood pressure (DBP). Table contains regression coefficients (*β*), standard errors (SE), *z*-statistics, two-sided *p*-values, and 95% confidence intervals from linear models fitted separately for SBP and DBP. Each model included all drug classes and confounders simultaneously as well as accounted for multiple measures and was specified as: SBP or DBP ~ ACEi + ARB + CCB + beta blocker + diuretic + statin + GP + ASCEN + age + (1|eid)

| Trait | Drug | $\beta$ | SE | $z$ | $p$ | 95% CI |
| --- | --- | --- | --- | --- | --- | --- |
| SBP | ACE inhibitor | -4.11 | 0.046 | -88.52 | $< 10^{-300}$ | [-4.20, -4.02] |
| SBP | Angiotensin receptor blocker | -4.91 | 0.066 | -74.17 | $< 10^{-300}$ | [-5.04, -4.78] |
| SBP | Calcium channel blocker | -5.24 | 0.048 | -108.90 | $< 10^{-300}$ | [-5.34, -5.15] |
| SBP | Beta blocker | -2.85 | 0.054 | -53.21 | $< 10^{-300}$ | [-2.95, -2.74] |
| SBP | Diuretic | -4.29 | 0.049 | -87.79 | $< 10^{-300}$ | [-4.39, -4.19] |
| SBP | Statin | -1.68 | 0.044 | -38.27 | $< 10^{-300}$ | [-1.77, -1.60] |
| DBP | ACE inhibitor | -1.71 | 0.031 | -54.30 | $< 10^{-300}$ | [-1.77, -1.65] |
| DBP | Angiotensin receptor blocker | -2.30 | 0.045 | -51.65 | $< 10^{-300}$ | [-2.39, -2.21] |
| DBP | Calcium channel blocker | -2.80 | 0.033 | -85.32 | $< 10^{-300}$ | [-2.87, -2.74] |
| DBP | Beta blocker | -1.70 | 0.036 | -47.03 | $< 10^{-300}$ | [-1.77, -1.63] |
| DBP | Diuretic | -1.25 | 0.034 | -37.20 | $< 10^{-300}$ | [-1.31, -1.18] |
| DBP | Statin | -1.39 | 0.029 | -47.66 | $< 10^{-300}$ | [-1.45, -1.34] |

**Table S3:** Counts of identified SNPs and whole exome sequencing loss-of-function variants per age group and LDL traits before and after treatment. Only variants at *FDR <* 0.05 are reported.

| Trait | Age | #SNPs | #LoF |
| --- | --- | --- | --- |
| LDL pre-treatment | pooled | 77 | 6 |
| LDL pre-treatment | < 50 | 943 | 32 |
| LDL pre-treatment | 50 - 55 | 265 | 9 |
| LDL pre-treatment | 56 - 60 | 55 | 1 |
| LDL pre-treatment | 60 - 64 | 1 | 1 |
| LDL pre-treatment | 65+ | 1 | 2 |
| LDL post-treatment | pooled | 8 | 2 |
| LDL post-treatment | < 50 | 7 | 1 |
| LDL post-treatment | 50 - 55 | 1 | 0 |
| LDL post-treatment | 56 - 60 | 1 | 0 |
| LDL post-treatment | 60 - 64 | 1 | 1 |
| LDL post-treatment | 65+ | 1 | 0 |

**Table S4:** Sensitivity analyses summary for SBP and DBP age-pooled and age-split variants. Panels report numbers of detected variants for the basic analysis and each sensitivity setup. Percentages in parentheses indicate the fraction retained relative to the corresponding basic analysis for that column. A 50 kb window was used as a threshold for variant overlap. Panel A shows a per-setup overview while the following tables show per-phenotype overlap.

| **(A) Overall counts (age-pooled and age-split)** |  |  |  |  |
| --- | --- | --- | --- | --- |
| Setup | SBP age-pooled | DBP age-pooled | SBP age-split | DBP age-split |
| Basic | 559 (100.0%) | 659 (100.0%) | 2499 (100.0%) | 2482 (100.0%) |
| No statins | 537 (80.3%) | 611 (79.2%) | 2439 (94.7%) | 2461 (93.9%) |
| Related up to 3rd | 581 (77.8%) | 677 (76.2%) | – | – |
| + prior CVD | 557 (99.1%) | 227 (34.6%) | 2495 (99.8%) | 2477 (99.8%) |
| Shifted age groups | – | – | 2019 (60.4%) | 2437 (70.5%) |
| 4 age groups | – | – | 2002 (84.4%) | 2019 (84.6%) |
| 30 day prescription window | 554 (69.8%) | 532 (54.5%) | – | – |

| **(B) SBP age-pooled per phenotype** |  |  |  |  |  |
| --- | --- | --- | --- | --- | --- |
| Setup | DBP post | DBP pre | ALS | Mesure No. | Statin |
| Basic | 1 (100.0%) | 439 (100.0%) | 1 (100.0%) | 215 (100.0%) | 5 (100.0%) |
| No statins | 2 (0.0%) | 396 (74.7%) | 1 (100.0%) | 214 (90.7%) | 0 (0.0%) |
| Related up to 3rd | 1 (0.0%) | 446 (73.1%) | 2 (0.0%) | 226 (82.3%) | 4 (100.0%) |
| + prior CVD | 0 (0.0%) | 8 (2.7%) | 0 (0.0%) | 214 (99.5%) | 2 (40.0%) |
| 30 day prescription window | 1 (0.0%) | 213 (40.5%) | 1 (0.0%) | 313 (82.8%) | 4 (100.0%) |

| **(C) DBP age-pooled per phenotype** |  |  |  |  |  |
| --- | --- | --- | --- | --- | --- |
| Setup | SBP post | SBP pre | ALS | Mesure No. | Statin |
| Basic | 2 (100.0%) | 330 (100.0%) | 2 (100.0%) | 220 (100.0%) | 5 (100.0%) |
| No statins | 1 (50.0%) | 309 (73.0%) | 2 (100.0%) | 224 (93.2%) | 0 (0.0%) |
| Related up to 3rd | 0 (0.0%) | 341 (74.5%) | 4 (100.0%) | 231 (82.7%) | 4 (100.0%) |
| + prior CVD | 2 (100.0%) | 330 (100.0%) | 0 (0.0%) | 220 (100.0%) | 2 (40.0%) |
| 30 day prescription window | 0 (0.0%) | 296 (62.7%) | 1 (50.0%) | 252 (80.5%) | 4 (100.0%) |

| **(D) SBP age-split per phenotype** |  |  |  |  |  |  |
| --- | --- | --- | --- | --- | --- | --- |
| Setup | SBP post<br>< 50 | SBP post<br>56–60 | SBP pre<br>< 50 | SBP pre<br>50–55 | SBP pre<br>61–64 | Median age<br>< 50 |
| Basic | 3 (100.0%) | 1 (100.0%) | 756 (100.0%) | 56 (100.0%) | 2 (100.0%) | 651 (100.0%) |
| No statins | 12 (100.0%) | 1 (100.0%) | 700 (90.7%) | 52 (64.3%) | 1 (0.0%) | 646 (97.1%) |
| + prior CVD | 3 (100.0%) | 1 (100.0%) | 756 (100.0%) | 56 (100.0%) | 0 (0.0%) | 651 (100.0%) |
| Shifted age groups | 12 (66.7%) | 1 (0.0%) | 778 (71.6%) | 55 (17.9%) | 6 (0.0%) | 100 (11.7%) |

| Setup | ARB<br>< 50 | Mesure No.<br>< 50 | Mesure No.<br>50–55 | Mesure No.<br>56–60 | Mesure No.<br>61–64 | Mesure No.<br>65+ | Statin<br>< 50 |
| --- | --- | --- | --- | --- | --- | --- | --- |
| Basic | 11 (100.0%) | 1002 (100.0%) | 12 (100.0%) | 2 (100.0%) | 5 (100.0%) | 5 (100.0%) | 1 (100.0%) |
| No statins | 11 (100.0%) | 997 (97.9%) | 11 (91.7%) | 2 (100.0%) | 7 (100.0%) | 5 (100.0%) | 0 (0.0%) |
| + prior CVD | 11 (100.0%) | 1002 (100.0%) | 10 (83.3%) | 2 (100.0%) | 5 (100.0%) | 5 (100.0%) | 1 (100.0%) |
| Shifted age groups | 7 (63.6%) | 1034 (85.9%) | 6 (25.0%) | 3 (50.0%) | 10 (80.0%) | 9 (100.0%) | 0 (0.0%) |

| **(E) DBP age-split per phenotype** |  |  |  |  |  |  |
| --- | --- | --- | --- | --- | --- | --- |
| Setup | ACEi<br>< 50 | DBP post<br>56–60 | DBP pre<br>< 50 | DBP pre<br>50–55 | DBP pre<br>61–64 | DBP pre<br>65+ |
| Basic | 1 (100.0%) | 2 (100.0%) | 871 (100.0%) | 68 (100.0%) | 1 (100.0%) | 3 (100.0%) |
| No statins | 1 (100.0%) | 5 (100.0%) | 825 (89.6%) | 65 (64.7%) | 1 (0.0%) | 2 (66.7%) |
| + prior CVD | 1 (100.0%) | 2 (100.0%) | 871 (100.0%) | 68 (100.0%) | 0 (0.0%) | 1 (33.3%) |
| Shifted age groups | 1 (100.0%) | 0 (0.0%) | 741 (69.6%) | 51 (22.1%) | 2 (0.0%) | 5 (33.3%) |

| Setup | Median age<br>< 50 | ARB<br>< 50 | Mesure No.<br>< 50 | Mesure No.<br>50–55 | Mesure No.<br>56–60 | Mesure No.<br>61–64 | Mesure No.<br>65+ | Statin<br>< 50 |
| --- | --- | --- | --- | --- | --- | --- | --- | --- |
| Basic | 598 (100.0%) | 9 (100.0%) | 906 (100.0%) | 11 (100.0%) | 4 (100.0%) | 7 (100.0%) | 6 (100.0%) | 1 (100.0%) |
| No statins | 599 (96.8%) | 9 (100.0%) | 920 (98.3%) | 11 (100.0%) | 4 (100.0%) | 8 (100.0%) | 6 (100.0%) | 0 (0.0%) |
| + prior CVD | 598 (100.0%) | 9 (100.0%) | 906 (100.0%) | 9 (81.8%) | 4 (100.0%) | 7 (100.0%) | 6 (100.0%) | 1 (100.0%) |
| Shifted age groups | 643 (57.0%) | 5 (55.6%) | 954 (84.5%) | 5 (18.2%) | 3 (75.0%) | 11 (85.7%) | 9 (100.0%) | 0 (0.0%) |

**Table S5:** Detection limit. Minimum percentage of variance explained by SNPs that can be detected in our analysis. See Methods for details about the calculations.

| trait | age-group | detection-limit [% var. expl.] |
| --- | --- | --- |
| ACE inhibitor | 1 | 0.533 |
| ACE inhibitor | 2 | 0.24 |
| ACE inhibitor | 3 | 0.166 |
| ACE inhibitor | 4 | 0.139 |
| ACE inhibitor | 5 | 0.096 |
| SBP post | 1 | 0.21 |
| SBP post | 2 | 0.097 |
| SBP post | 3 | 0.061 |
| SBP post | 4 | 0.046 |
| SBP post | 5 | 0.031 |
| SBP pre | 1 | 0.017 |
| SBP pre | 2 | 0.017 |
| SBP pre | 3 | 0.018 |
| SBP pre | 4 | 0.021 |
| SBP pre | 5 | 0.022 |
| ARB | 1 | 1.787 |
| ARB | 2 | 0.734 |
| ARB | 3 | 0.424 |
| ARB | 4 | 0.308 |
| ARB | 5 | 0.189 |
| beta blocker | 1 | 0.623 |
| beta blocker | 2 | 0.324 |
| beta blocker | 3 | 0.219 |
| beta blocker | 4 | 0.18 |
| beta blocker | 5 | 0.115 |
| CCB | 1 | 0.894 |
| CCB | 2 | 0.383 |
| CCB | 3 | 0.222 |
| CCB | 4 | 0.171 |
| CCB | 5 | 0.101 |
| diuretic | 1 | 0.803 |
| diuretic | 2 | 0.347 |
| diuretic | 3 | 0.211 |
| diuretic | 4 | 0.165 |
| diuretic | 5 | 0.112 |
| statin | 1 | 0.622 |
| statin | 2 | 0.233 |
| statin | 3 | 0.131 |
| statin | 4 | 0.095 |
| statin | 5 | 0.067 |
| DBP post | 1 | 0.233 |
| DBP post | 2 | 0.11 |
| DBP post | 3 | 0.069 |
| DBP post | 4 | 0.052 |
| DBP post | 5 | 0.035 |
| DBP pre | 1 | 0.018 |
| DBP pre | 2 | 0.018 |
| DBP pre | 3 | 0.019 |
| DBP pre | 4 | 0.022 |
| DBP pre | 5 | 0.023 |
| ACE inhibitor | all | 0.047 |
| SBP post | all | 0.017 |
| SBP pre | all | 0.006 |
| ARB | all | 0.105 |
| beta blocker | all | 0.059 |
| CCB | all | 0.055 |
| diuretic | all | 0.06 |
| statin | all | 0.034 |
| DBP post | all | 0.018 |
| DBP pre | all | 0.006 |

**Table S6:**
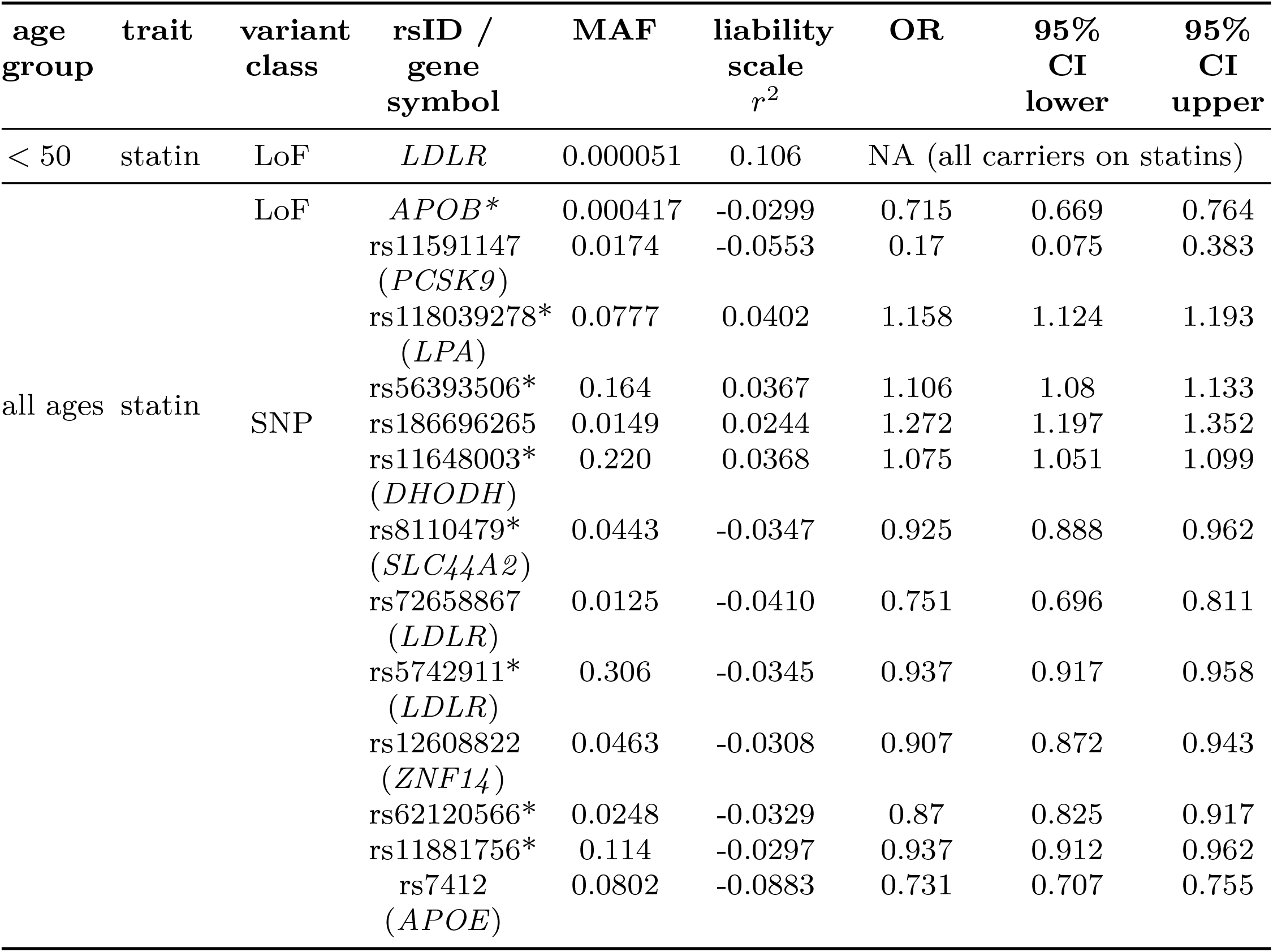
Genetic variants associated with statin-taking in the UK Biobank blood pressure management dataset. For each detected whole exome sequence LoF variant, the gene symbol is given. For each SNP, the rsID/gene symbol column gives the SNP rsID and the gene symbols in brackets if the SNP falls inside a protein-coding gene. The liability scale squared correlation (*r*^2^) between the variant and the phenotype is given as well as the odds ratio (OR) for variant carriers (both homo- and heterozygotes) being on a drug as well as the lower and upper bounds of the 95% confidence interval (CI). A star indicates variants detected only in the on-drugs subset of the UKB participants (see Methods). MAF - minor allele frequency.

## Supplementary Note 1: Supplementary Methods

### Extraction of blood pressure and LDL cholesterol measurements, diagnoses and prescription data from the UK Biobank

Blood pressure measurements and prescription data were extracted from the UK Biobank Research Access platform with pyspark (v 3.2.3) and then processed with Hail (see Code Availability). All of the UK Biobank participants with GP (General Practice) registration were selected and then all GP, hospital and death registry records were extracted for these participants with accompanying dates. For prescription extraction, a dictionary of medications used in blood pressure and cardiovascular disease management was prepared and codes in each of the prescription coding systems used in the UK Biobank GP data were translated into Anatomical Therapeutic Chemical (ATC) codes. The free text fields were also parsed for medication names.

For blood pressure (BP) measurements relevant Read2/CTV3 codes were extracted together with the measurement value. Measurements performed in the UK Biobank Assessment Center were also added to the dataset. For BP measurements in the GP data with two numerical values, the higher value was assigned to systolic blood pressure (SBP) and the lower to diastolic blood pressure (DBP). For single-value measurements, the value has been assigned to DBP if ‘diastolic’ was present in the procedure code description, otherwise to SBP. The following hard filters were applied to remove impossible measurements: age *>* 0, SBP *>* 40, SBP *<* 375, DBP *<* 365 and DBP *>* 30. A full json dictionary of all of the above items can be found in the Code Availability section. LDL cholesterol (LDL-C) measurements were extracted following a procedure published elsewhere [11] using numerical values from a combination of Read v2 and CTV3 clinical codes: 44P6. (serum LDL), 44PI. (calculated LDL), 44PD. (serum fasting LDL), and XaIp4 (calculated LDL, only in CTV3). To remove for biologically implausible outliers, we applied a strict filtering process. Values outside the range of 0.26 mmol/L to 10.3 mmol/L were excluded from the analysis.Final extracted raw datasets that included all blood pressure or LDL measurements annotated with date, source (GP or Assessment Center), prescription information, and participants age at the time of measurement. Prescriptions for major cardiometabolic drug classes have been extracted and grouped into six drug categories according to their ATC codes (Table S1). To control for CVD that may have been diagnosed in a hospital setting (UK Biobank hospital records are considered more complete) prior to the first measurement available for each participant we collected CVD diagnoses from both hospital and GP records. These included codes from the ICD10 chapters I20, I21, I22, I23, I25, I30, I31, I33, I34, I35, I36, I37, I38, I39, I42, I44, I45, I46, I47, I48, I49, I50, I51 together with equivalent Read2/CTV3 and ICD9 codes and their dates. These CVD diagnoses were recoded into binary prior cvd phenotype and included in sensitivity analyses. For the age-pooled version prior cvd was given 1 if the first measurement for the participant followed the diagnosis date, while in the age-split approach prior cvd was set to 1 if the first measurement within an age group followed the diagnosis date.

### Construction of analysis-ready measurement datasets

First, each measurement was assigned drugs associated with it using a 60-day window from prescription. This means that if a prescription for a drug in one of the six categories had been issued within 1 to 60 days prior to the measurement, the binary treatment indicator for this drug class was set to 1. Additional sensitivity datasets with a shorter window (1 to 30 days) were also prepared for comparison. Measurements were separated into pre- and post-treatment in the following procedure: We considered all measurements that occurred before any recorded treatment or at least two years after the last recorded treatment as pre-treatment. Measurements annotated with any of the investigated drugs within the choses prescription windod were considered post-treatment. Measurements for each participant were then aggregated. In the age-pooled approach, we averaged pre- and post-treatment SBP, DBP or LDL values and the maximum value from the remaining variables (drug classes, measurement source) was used in the aggregation. If a person had no treatment records, the post-treatment SBP/DBP/LDL value was set as missing. In the age-split approximation, measurements for each participant were first aggregated into age intervals averaging pre- and post-treatment and choosing the maximum value for other variables. The intervals were chosen as to minimize the standard deviation of group size and the main set consisted of five age intervals. We additionally prepared a set with only four age groups, based on the same procedure. To check if specific years influence the results, we also made a 5 age-group dataset with shifted group edges by moving the two lower internal cut-points downward one year and the two upper internal cut-points upward one year. If a participant did not have on- or off-drug measurements within an interval, those were set as missing. Each dataset was also augmented with age first and last seen in the GP records, year of birth and median age at measurement. All continuous variables were then adjusted for covariates (sex, 20 genetic principal components, assessment center, genotyping batch, coordinates) and standardized. All analyses were performed on these participant-level aggregated datasets, with repeated measurements summarized within individuals (and within age groups where applicable).

### Extraction of data to evaluate individual drug responses

Major classes of cardiological drugs were defined based on the British National Formulary (BNF) classification system. Drugs were identified by filtering lookup tables from the UK Biobank resource (see biobank.ndph.ox.ac.uk/ukb/ukb/auxdata/ primarycare codings.zip) according to the appropriate BNF paragraphs, sections and chapters.

- Diuretics: Defined as drugs listed under section 2.2 (“Diuretics”).
- Statins: Defined as drugs listed under section 2.12 (“Lipid-Regulating Drugs”).
- Beta-Blockers: Defined as drugs listed under section 2.4 (“Beta-Adrenoceptor Blocking Drugs”).
- Calcium-Channel Blockers (CCBs): Defined
- as drugs from paragraph 2.6.2 (“Calcium-Channel Blockers”).
- Angiotensin-Converting Enzyme Inhibitors (ACE Inhibitors): Defined as drugs from subparagraph 2.5.5.1 (“Angiotensin-Converting Enzyme Inhibitors”).
- Angiotensin II Receptor Antagonists: Defined as drugs from subparagraph 2.5.5.2 (“Angiotensin II Receptor Antagonists”).
- Renin Inhibitors: Defined as drugs from subparagraph 2.5.5.3. (“Renin Inhibitors”).

For each group, active substances were matched with corresponding brand names, using the classification data to associate these substances with entries in different drug coding systems (Read2, CTV3, DMD). This yielded a set of codes with which the electronic health records from UK Biobank was then filtered.

In addition, dosage and quantity information was extracted for each drug code. Where available, dosage and quantity values were directly retrieved from the structured data fields associated with the codes. In cases where such structured information was missing, quantity values were extracted from the textual descriptions within the coding system using regular expression techniques (e.g., identifying patterns such as “mg”, “ml”, “mg/ml”). This two-step approach ensured that dosage data was included wherever possible, providing a more complete picture of each drug’s use and characteristics (see Code Availability).

In the case of drug combinations, cases where drugs were given together with diuretics or calcium channel blockers were included. When the combination was not explicitly stated in the data (e.g. ‘with diuretics’ or ‘with a calcium channel blocker’), the information was noted and the drugs were treated as drugs of the relevant group (diuretic or CCB) without specifying the exact substance.

To ensure the integrity and quality of the dataset, records with missing values were removed. In particular, the following data were removed:

- Missing Dose Information: A total of 0.86% of records were found to be missing dose information.
- Missing Quantity Information: 13.85% of records were missing information on the quantity.
- Missing Dates: A small proportion (0.002%) of records had missing date information.

Combined, these filters removed 14.10% of all prescriptions due to missing data. In addition, individuals with no ARB prescriptions were excluded from the analysis. As a result, the final dataset included 19,114 individuals.

We then defined therapies and calculated the proportion of days covered (PDC) for ARBs. To ensure the correct classification of therapies, we identified the intervals between prescriptions for each patient and drug, which were added to the database. We defined a therapy for each patient as a set of prescriptions involving the same drug and a similar expected daily dose.

Prescriptions were grouped into separate therapies based on the interval between adjacent prescriptions. If the interval between two consecutive prescriptions exceeded 90 days, adjusted for the number of tablets prescribed, they were considered separate therapies. In addition, the algorithm considered the median mean daily dose over 11 consecutive prescriptions. It compared consecutive median values and considered that a significant dose change occurred when the dose was changed at least 2 times. When such a change occurred, treatment was divided into separate treatment periods.

For ARB therapy, the percentage of days on treatment (PDC), an indicator used to assess medication adherence, was also calculated. The calculation was based on the estimated daily dose of the drug based on the fact that the minimum dose is no less than 1 tablet.

Finally, we then applied specific inclusion criteria by removing therapies with a PDC below 50% or exceeding 200%. This filtering step eliminated approximately 1,200 therapies, ensuring that only reliable adherence data remained. After the PDC filtering, the final dataset contained approximately 17,000 individuals who were prescribed ARBs.

ARB therapy data was then aggregated for each participant to extract drug response proxy phenotypes. This included collection of counts of different ARB substances prescribed, total number of different drug classes prescribed to ARB-takers, duration of longest ARB therapy, average doses of most common ARBs and binary traits indicating if ARB was first or last therapy for a participant, if it was ever augmented with another drug, if the dose was ever increase or if an ARB was discontinued and a drug from another class was introduced. This process resulted in 17 ARB response traits.

### Detailed analysis of the BP measurements and prescription patterns

As the BP data collected comes from both the assessment centers and EHRs (primary care records) we analyzed the data with regards to:

- Time interval between each drug-annotated measurement and the date of prescription - here, as in the whole study, measurements made on the same date as prescription were not counted as on-drugs unless there was another prescription in 1-60 days prior.
- Average number of pills in a single prescription for all of the investigated drugs - for additional guidance on prescription window choice.
- Measurements stratified by number and source - to investigate differences in patterns of on- and off-drug measurements depending where the BP was measured as well as differences in BP values. Note that we use measurement source as a covariate throughout the study.
- Drug effects - in the aggregated data we ran linear mixed-effects models with blood pressure (or LDL) as the outcome, drug classes and covariates as fixed effects, and a participant-specific random intercept to account for repeated measurements within individuals.

## Supplementary Note 2: Supplementary results and discussion

### Comparison with standard approaches

To give a better overview to the types of discoveries made with our approach we also conducted standard GWAS. For baseline traits (SBP and DBP), we ran basic models using confounders (median age at measurement, number of measurements, age of first and last record, year of birth) along the usual covariates (sex, 20 genetic principal components, assessment center, genotyping batch, coordinates) and show a high concordance of p-values with Pearson *r*^2^ = 0.74 (Figure S14A). Two variants that are notable outliers and show very low p-values in marginal testing but not in our graphical model (rs77870048 and rs62053262) are within 5kb from each other and in high LD (*r*^2^ = 0.83) explaining their lower p-values in standard setting. At our chosen FDR threshold of 5%, many of the loci selected by out method are above the customary genome-wide significant threshold of 5 ·10^−8^. To elaborate on this, we evaluated which variants are selected by our method versus standard baseline GWAS. For this, we looked at marginal correlation values between variants and pre-treatment SBP in the age-pooled and first age group (*<* 50 years) in the age-split approach and show what these methods select largely non-overlapping variant sets with CI-GWAS selecting more rare variants without high LD to other variants (Figure S14B and Figure S9). This is in accordance with the goal of our graphical approach which is to identify trait-specific loci.

### Overlap between CI-GWAS result sets

Between the age-pooled and age-split analyses, the variant sets that we discover are largely non-overlapping (Figure S7B), see Figure S9 for explanation. In the age-split analysis, there is no overlap between variants in any of the age groups neither for DBP and nor for SBP. For each of the SBP and DBP-associated variant sets that exceeded 50 variants, we mapped SNPs to genes using Ensemble’s VEP joined with the genes with associated LoF variants (see Supplementary Data Tables). First, as expected, we observe that the gene-level overlap between age-split and age-pooled analyses for SBP and DBP is higher than the SNP-level overlap (Figure S13). This overlap is higher for the youngest age-group (54 genes for SBP and 33 genes for DBP) than for later age groups where ther is very little overlap (1 gene for SBP and 5 genes for DBP). Moreover, the single overlapping gene for SBP is *MSI2* is a very large gene which spans 424 kb, suggesting a spurious colocalisation [55]. Taken together, we again confirm that we identify age-specific genetic variation exemplified by the minimal shared signal between the age-pooled version and age group 50-55.

### Further description of loss-of-function variants

Conditional on the SNP associations and all other variants genome-wide, we also discover many genes with loss-of-function (LoF) variants associated with the pretreatment measures (Table 1). Examples of LoF variant associations (with 3 or more carriers within the UKB) include *SMAD2* detected in the pre-treatment SBP from all ages. This gene produces a signaling protein involved in artery remodeling [26, 27]. For DBP in all ages, we report associations with LoFs in *BAG6*. Common variants in this gene have been previously associated with BP [56]. In the youngest age group, for SBP, we detect association with LoF variants in *ANGPTL1* - a gene known for regulation of angiogenesis [57]. For DBP, in the same age group, we replicate association with *PDE3A*. This gene encodes a phosphodiesterase and its gain of function mutations cause brachydactyly with severe hypertension [28, 29]. In summary, in our WES analysis, conditioned on common variant background, we replicate known genetic syndromes associated with hypertension as well as provide novel findings. All LoF associations are listed in Supplementary Data Tables.

### Enrichment analysis

As rare and common variants act together to influence complex traits, we join genes with LoF variants discovered in our model together with the genes annotated to SNPs in a gene-level enrichment analysis [58, 59]. We performed gene level enrichment analyses with the (1) Reactome Pathways Database for pathway enrichment; (2) Gene Ontology (GO) Molecular Function to integrate the biological function of discovered genes; (3) ChEA and ENCODE transcription factor database to search for possible common regulators of the discovered genes (Figure 3C, Supplementary Data Tables).

From all of the 378 cardiovascular-health related traits within the GWAS catalog, the top overrepresented terms are systolic blood pressure, diastolic blood pressure, electrocardiogram features and medication use for hypertension (Figure 3C). There is no significant enrichment neither for SBP GWAS genes in the 27 genes we discover in the 56-60 age group nor for the DBP GWAS in the 39 geneswe discover for DBP, suggesting that these are novel findings (Figure 3C). Notably, the highest overlap with the genes associated with medication use for hypertension is observed in the ¡50 years-old age group (14 out of the 461 SBP age ¡50 genes, p-value = 8.45 ·10^−7^, OR = 5.65). Taken together, our results are highly replicable and suggest that the genetic variation for BP is shaped prior to mid-life, with limited evidence for SNPs whose effects contribute to BP variation later in life.

In a functional enrichment analysis, we find terms only in the youngest age groups (*<* 50) that are associated with organism development: axon guidance for SBP (p-value = 0.00036, OR = 2.21) and myogenesis for DBP (p-value = 0.0007, OR = 8.07) (Figure 3C). Based on ChIP-seq data, the discovered genes share common regulatory pathways across the age groups. However, the enrichment for the top transcription factor (TF) binding sites is most pronounced in the youngest age groups for both SBP and DBP-associated gene (Figure 3C). The TFs that we discover are REST (RE1-silencing transcription factor), SMAD4 (a central *TGF* −*β* pathway mediator), AR (androgen receptor), and SUZ12 (a Polycomb group protein); all with crucial functions in human development. REST affects potassium channel expression and vascular smooth muscle cell proliferation [60]. SMAD4 regulates vascular and cardiac development and remodeling. Gain of function mutations in SMAD4 cause hypertension in about 15% of patients [61]. In animal studies, AR signaling has been linked to hypertension in both males and females [62] [63]. SUZ12 belongs to a complex that epigenetically silences gene expression and mediates heart development and its aberrant signaling has been previously implicated in pulmonary hypertension [64] [65].

### Associations with possible confounders

Across the analyses we discover multiple SNP and LoF variants associated with likely confounders. We note that we find no variants associated with the year of birth, no SNPs associated with age at first record in the dataset (rs187729359) and two SNPs associated with age at last record in the dataset (rs147750638, rs78651908). All four are discovered in the age-pooled analysis and only rs147750638 is located within a protein-coding gene locus: *MARCHF4*, a membrane-bound E3 ubiquitin ligase, variants in which have been previously associated with height, smoking initiation and acute myeloid leukemia in the GWAS catalog. In the age-pooled model, we find 220 variants associated with number of measurements and none with median age at measurement. Among these 220 variants, we find SNPs in genes previously associated with risk of type 2 diabetes (*TCF7L2* and *HNF4A*). In *TCF7L2*, which is the most potent risk locus for type 2 diabetes risk [66], we find a previously unreported SNP associated positively with number of measurements (Pearson *r*^2^ = 0.008). We therefore hypothesize that the inclusion of the “number of measurements” phenotype may adjust for comorbidities as patients with accompanying diseases are more often measured.

In the age-split models, we find variants associated with number of measurements for each age group, but just as with SBP/DBP these numbers tend to decrease for older participants (age *<* 50 - 803 variants, age 50− 55 - 11 variants, age 56 −60 - 4 variants, age 61 −64 - 7 variants, age *>* 64 - 6 variants). We also find 597 variants associated with the median age at measurement in the youngest age group. This is expected as patients with genetically-determined syndromes should have more BP measurements early in life. Accordingly, 45 of these variants are LoF and include genes like *KCNQ4* associated with hearing loss in humans and known for its regulation of vascular tone [67] and Epidermal Growth Factor (*EGF*) which expression determines appropriate kidney function [68]. A pooled enrichment analysis of all of the SNPs and LoF variants associated with the possible confounders in the age-split approach points to multiple cardiovascular phenotypes. According to the PhenGenI Association 2021 database (accessed via Enrichr), 114 genes (13%) are associated with HLD cholesterol levels (adjusted p-value 1.09 ·10^−7^). Other examples of overrepresented terms include 65 genes associated with asthma (adjusted p-value 1.85e-4), 69 with myocardial infraction (1.85 ·10^−4^), 80 with stroke (3.89 ·10^−4^) and 162 with Pervasive Child Development Disorders (4.61 ·10^−4^). In summary, we show that the possible confounding factors included in our model are associated with variants linked to cardiovascular disease outcomes.

### Description of Supplementary Data Tables

First tab of the Supplementary Data Tables contains a guide for their contents.

**Supplementary Data Table A** Sample sizes for all traits across all model setups.

**Supplementary Data Table B** Complete results from the graphical model across all analysis setups including all sensitivity analyses.

**Supplementary Data Table C** All SNPs associated with pretreatment SBP and DBP in both the age-pooled and the age-split model.

**Supplementary Data Table D** All LoF variants associated with SBP/DBP in the age-pooled and age-split analyses.

**Supplementary Data Table E** Standard GWAS results for the SNPs discovered as associated with post-treatment SBP and DBP in the graphical model.

**Supplementary Data Table F** Standard GWAS results for the SNPs discovered as associated with post-treatment LDL in the graphical model.

**Supplementary Data Table G** Gene level enrichment analyses that includes both SNPs and the genes with LoF variants with the GWAS Catalog. This table includes any results with adjusted p-value *<* 0.05 as reported by Enrichr.

**Supplementary Data Table H** Gene level enrichment analyses that includes both SNPs and the genes with LoF variants with Reactome Pathways, ChEA TFs and Gene Ontology Molecular Function. This table includes any results with p-value *<* 0.05 as reported by Enrichr.

**Supplementary Data Table I** Gene lists compiled for each phenotype with *>* 50 variants from SNPs (mapped via VEP) and LoFs used for enrichment analysis.

**Supplementary Data Table J** Variants (both SNPs and LoFs) associated with potential confounding variables in the graphical model.

**Supplementary Data Table K** Associations of drug-response SNPs in Open Targets Database including Finngen. This table includes replication information for discovered SNPs and their LD partners in multiple datasets, but only up to r6 of FinnGen. Associations for rs12608822 and rs186696265 have been searched and confirmed in the r12 FinnGen release (see https://r12.finngen.fi/variant/19:19730943-C-T and https://r12.finngen.fi/variant/6:160690668-C-T)

**Supplementary Data Table L** All SNP and LoF variants discovered as associated with ARB treatment indicators in the graphical model.

**Supplementary Data Table M** Standard GWAS results for the ARB-taking associated variants from Table L.

**Supplementary Data Table N** Regression results testing whether ARB-taking associated variants are also associated with ARB drug response proxy phenotypes.

**Supplementary Data Table O** Multiple regression models predicting ARB drug response phenotypes using all SBP and/or DBP associated SNPs as predictors.

**Supplementary Data Table P** Lookup table mapping all main graphical model results to standard GWAS p-values (accompanies Figure S14A).

**Supplementary Data Table Q** Statistics for Figure 4B.

**Supplementary Data Table R** Statistics for number of pills per prescription for cardiometabolic drugs (accompanies Figure S6A).

## References

[1] Pirmohamed, M. Pharmacogenomics: current status and future perspectives. Nature Reviews Genetics 24, 350–362 (2023).

[2] McInnes, G., Yee, S. W., Pershad, Y. & Altman, R. B. Genomewide association studies in pharmacogenomics. Clin. Pharmacol. Ther. 110, 637–648 (2021).

[3] Kiiskinen, T. et al. Genetic predictors of lifelong medication-use patterns in cardiometabolic diseases. Nature Medicine 29, 209–218 (2023).

[4] Cordioli, M. et al. Socio-demographic and genetic risk factors for drug adherence and persistence: a retrospective nationwide and biobank study across 5 medication classes and 1 845 665 individuals. medRxiv (2024).

[5] Wu, Y. et al. Genome-wide association study of medication-use and associated disease in the uk biobank. Nature Communications 10, 1891 (2019).

[6] Postmus, I. et al. Pharmacogenetic meta-analysis of genome-wide association studies of ldl cholesterol response to statins. Nature Communications 5, 5068 (2014).

[7] Oni-Orisan, A. et al. The impact of adjusting for baseline in pharmacogenomic genome-wide association studies of quantitative change. NPJ Genom. Med. 5, 1 (2020).

[8] Salvi, E. et al. Genome-wide and gene-based meta-analyses identify novel loci influencing blood pressure response to hydrochlorothiazide. Hypertension 69, 51–59 (2017).

[9] McDonough, C. W. et al. Adverse cardiovascular outcomes and antihypertensive treatment: A genome-wide interaction meta-analysis in the international consortium for antihypertensive pharmacogenomics studies. Clinical Pharmacology & Therapeutics 110, 723–732 (2021).

[10] Zhang, H., Chhibber, A., Shaw, P. M., Mehrotra, D. V. & Shen, J. A statistical perspective on baseline adjustment in pharmacogenomic genome-wide association studies of quantitative change. npj Genomic Medicine 7, 33 (2022).

[11] Sadler, M. C. et al. Leveraging large-scale biobank ehrs to enhance pharmacogenetics of cardiometabolic disease medications. Nature Communications 16, 2913 (2025).

[12] Sadowski, M. et al. Characterizing the genetic architecture of drug response using gene-context interaction methods. Cell Genomics 4 (2024).

[13] Zhou, K. et al. Variation in the glucose transporter gene slc2a2 is associated with glycemic response to metformin. Nature Genetics 48, 1055–1059 (2016).

[14] Robinson, M. R. et al. Genotype–covariate interaction effects and the heritability of adult body mass index. Nat Genet 49, 1174–1181 (2017). Publisher: Nature Publishing Group.

[15] Cui, R., Groot, P. & Heskes, T. Learning causal structure from mixed data with missing values using Gaussian copula models. Stat Comput 29, 311–333 (2019).

[16] Machnik, N. et al. Causal inference for multiple risk factors and diseases from genomics data (2025). BioRxiv 2023.12.06.570392.

[17] Xie, Z. et al. Gene set knowledge discovery with enrichr. Current protocols 1, e90 (2021).

[18] Kurki, M. I. et al. Finngen provides genetic insights from a well-phenotyped isolated population. Nature 613, 508–518 (2023).

[19] Cerezo, M. et al. The nhgri-ebi gwas catalog: standards for reusability, sustainability and diversity. Nucleic acids research 53, D998–D1005 (2025).

[20] Keaton, J. M. et al. Genome-wide analysis in over 1 million individuals of european ancestry yields improved polygenic risk scores for blood pressure traits. Nature Genetics 56, 778–791 (2024).

[21] McDonough, C. W. et al. Adverse cardiovascular outcomes and antihypertensive treatment: A genome-wide interaction meta-analysis in the international consortium for antihypertensive pharmacogenomics studies. Clinical Pharmacology & Therapeutics 110, 723–732 (2021).

[22] Kilic, T., Okuno, K., Eguchi, S. & Kassiri, Z. Disintegrin and metalloproteinases (adams [a disintegrin and metalloproteinase] and adamtss [adams with a thrombospondin motif]) in aortic aneurysm. Hypertension 79, 1327–1338 (2022).

[23] Avery, C. L. et al. Drug–gene interactions and the search for missing heritability: a cross-sectional pharmacogenomics study of the qt interval. The pharmacogenomics journal 14, 6–13 (2014).

[24] Sadowski, M. et al. Characterizing the genetic architecture of drug response using gene-context interaction methods. Cell Genomics 4 (2024).

[25] Yengo, L. et al. Meta-analysis of genome-wide association studies for height and body mass index in 700000 individuals of european ancestry. Human Molecular Genetics 27, 3641–3649 (2018).

[26] Deng, H. et al. Activation of Smad2/3 signaling by low fluid shear stress mediates artery inward remodeling. Proceedings of the National Academy of Sciences 118, e2105339118 (2021). Publisher: Proceedings of the National Academy of Sciences.

[27] Bjørnstad, J. L. et al. Inhibition of SMAD2 phosphorylation preserves cardiac function during pressure overload. Cardiovascular Research 93, 100–110 (2012).

[28] Gallegos, F. et al. Decoding Monogenic Hypertension: A Review of Rare Hypertension Disorders. American Journal of Hypertension hpaf005 (2025).

[29] Chung, Y. W. et al. Targeted disruption of PDE3B, but not PDE3A, protects murine heart from ischemia/reperfusion injury. Proceedings of the National Academy of Sciences 112, E2253–E2262 (2015). Publisher: Proceedings of the National Academy of Sciences.

[30] Xiang, Q. et al. The association between the slco1b1, apolipoprotein e, and cyp2c9 genes and lipid response to fluvastatin: a meta-analysis. Pharmacogenetics and genomics 28, 261–267 (2018).

[31] Ference, B. A. et al. Effect of long-term exposure to lower low-density lipoprotein cholesterol beginning early in life on the risk of coronary heart disease: a mendelian randomization analysis. Journal of the American College of Cardiology 60, 2631–2639 (2012).

[32] Defesche, J. C. et al. Familial hypercholesterolaemia. Nature reviews Disease primers 3, 1–20 (2017).

[33] Altamirano, F. et al. Polycystin-1 assembles with kv channels to govern cardiomyocyte repolarization and contractility. Circulation 140, 921–936 (2019).

[34] Schaller, L. & Lauschke, V. M. The genetic landscape of the human solute carrier (slc) transporter superfamily. Human genetics 138, 1359–1377 (2019).

[35] He, J. et al. Solute carrier family 35 member f2 is indispensable for papillary thyroid carcinoma progression through activation of transforming growth factorβ type i receptor/apoptosis signal-regulating kinase 1/mitogen-activated protein kinase signaling axis. Cancer Science 109, 642–655 (2018).

[36] Che, B. et al. Slc35f2–syvn1–trim59 axis critically regulates ferroptosis of pancreatic cancer cells by inhibiting endogenous p53. Oncogene 42, 3260–3273 (2023).

[37] Rodriguez, S. et al. Aging and chronic high-fat feeding negatively affect kidney size, function, and gene expression in ctrp1-deficient mice. American Journal of Physiology-Regulatory, Integrative and Comparative Physiology 320, R19–R35 (2021).

[38] Zhang, K. et al. Genetic implication of a novel thiamine transporter in human hypertension. Journal of the American College of Cardiology 63, 1542–1555 (2014).

[39] Mosley, J. D. et al. A genome-wide association study identifies variants in kcnip4 associated with ace inhibitor-induced cough. The pharmacogenomics journal 16, 231–237 (2016).

[40] Lavertu, A., McInnes, G., Tanigawa, Y., Altman, R. B. & Rivas, M. A. LPA and APOE are associated with statin selection in the UK Biobank (2020). 2020.08.28.272765.

[41] Guan, Z.-W. et al. Pharmacogenetics of statins treatment: Efficacy and safety. Journal of Clinical Pharmacy and Therapeutics 44, 858–867 (2019).

[42] Benjamini, Y. & Yekutieli, D. The control of the false discovery rate in multiple testing under dependency. Ann. Stat. 29, 1165–1188 (2001).

[43] Strobl, E. V., Spirtes, P. L. & Visweswaran, S. Estimating and controlling the false discovery rate of the pc algorithm using edge-specific p-values. ACM Trans. Intell. Syst. Technol. 10 (2019). URL 10.1145/3351342.

[44] Li, J. & Wang, Z. J. Controlling the false discovery rate of the association/-causality structure learned with the pc algorithm. Journal of Machine Learning Research 10, 475–514 (2009). URL http://jmlr.org/papers/v10/li09a.html.

[45] Berisa, T. & Pickrell, J. K. Approximately independent linkage disequilibrium blocks in human populations. Bioinformatics 32, 283–285 (2016).

[46] Chang, C. C. et al. Second-generation plink: rising to the challenge of larger and richer datasets. GigaScience 4 (2015).

[47] Purcell, S. et al. Plink: a tool set for whole-genome association and population-based linkage analyses. Am J Hum Genet. 81, 559–575 (2007).

[48] Cui, R., Groot, P. & Heskes, T. L. Learning causal structure from mixed data with missing values using gaussian copula models. Stat Comput 29, 311–333 (2019).

[49] Pearson, K. On a New Method of Determining Correlation Between a Measured Character A, and a Character B, of which Only the Percentage of Cases Wherein B Exceeds (or Falls Short of) a Given Intensity is Recorded for Each Grade of A. Biometrika 7, 96–105 (1909).

[50] Pearson, K. Mathematical Contributions to the Theory of Evolution. VIII. On the Correlation of Characters Not Quantitatively Measurable. [Abstract]. Proceedings of the Royal Society of London 66, 241–244 (1899).

[51] Hunter, J. E. & Schmidt, F. L. Dichotomization of continuous variables: The implications for meta-analysis. Journal of Applied Psychology 75, 334–349 (1990). Place: US Publisher: American Psychological Association.

[52] Szustakowski, J. D. et al. Advancing human genetics research and drug discovery through exome sequencing of the uk biobank. Nature genetics 53, 942–948 (2021).

[53] Chang, C. C. et al. Second-generation PLINK: rising to the challenge of larger and richer datasets. GigaScience 4, s13742.–015–0047–8 (2015).

[54] Zhong, H. & Prentice, R. L. Bias-reduced estimators and confidence intervals for odds ratios in genome-wide association studies. Biostatistics 9, 621–634 (2008).

[55] Spence, J. P. et al. Specificity, length and luck drive gene rankings in association studies. Nature 649, 918–925 (2026).

[56] The International Consortium for Blood Pressure Genome-Wide Association Studies. Genetic variants in novel pathways influence blood pressure and cardiovascular disease risk. Nature 478, 103–109 (2011).

[57] Kubota, Y. et al. Cooperative interaction of Angiopoietin-like proteins 1 and 2 in zebrafish vascular development. Proc Natl Acad Sci U S A 102, 13502–13507 (2005).

[58] Kim, Y. J. et al. The contribution of common and rare genetic variants to variation in metabolic traits in 288,137 East Asians. Nat Commun 13, 6642 (2022).

[59] Williams, J. et al. Integrating Common and Rare Variants Improves Polygenic Risk Prediction Across Diverse Populations (2024). MedrXiv, 2024.11.05.24316779.

[60] Cheong, A. et al. Downregulated rest transcription factor is a switch enabling critical potassium channel expression and cell proliferation. Molecular cell 20, 45–52 (2005).

[61] Lin, A. E. et al. Gain-of-function mutations in smad4 cause a distinctive repertoire of cardiovascular phenotypes in patients with myhre syndrome. American journal of medical genetics Part A 170, 2617–2631 (2016).

[62] Davis, G. K. et al. Androgen receptor blockade differentially regulates blood pressure in growth-restricted versus ovarian deficient rats. Hypertension 74, 975– 982 (2019).

[63] Wu, C.-C. et al. Androgen-dependent hypertension is mediated by 20-hydroxy-5, 8, 11, 14-eicosatetraenoic acid–induced vascular dysfunction: role of inhibitor of κb kinase. Hypertension 57, 788–794 (2011).

[64] Bisserier, M. et al. Regulation of the methylation and expression levels of the bmpr2 gene by sin3a as a novel therapeutic mechanism in pulmonary arterial hypertension. Circulation 144, 52–73 (2021).

[65] Yu, J. et al. Long noncoding rna ahit protects against cardiac hypertrophy through suz12 (suppressor of zeste 12 protein homolog)-mediated downregulation of mef2a (myocyte enhancer factor 2a). Circulation: Heart Failure 13, e006525 (2020).

[66] del Bosque-Plata, L., Martínez-Martínez, E., Espinoza-Camacho, M.Á. & Gragnoli, C. The role of tcf7l2 in type 2 diabetes. Diabetes 70, 1220–1228 (2021).

[67] Chadha, P. S. et al. Reduced kcnq4-encoded voltage-dependent potassium channel activity underlies impaired β-adrenoceptor–mediated relaxation of renal arteries in hypertension. Hypertension 59, 877–884 (2012).

[68] Staruschenko, A., Palygin, O., Ilatovskaya, D. V. & Pavlov, T. S. Epidermal growth factors in the kidney and relationship to hypertension. American Journal of Physiology-Renal Physiology 305, F12–F20 (2013).

